# Immune Correlates Analysis of a Single Ad26.COV2.S Dose in the ENSEMBLE COVID-19 Vaccine Efficacy Clinical Trial

**DOI:** 10.1101/2022.04.06.22272763

**Authors:** Youyi Fong, Adrian B. McDermott, David Benkeser, Sanne Roels, Daniel J. Stieh, An Vandebosch, Mathieu Le Gars, Griet A. Van Roey, Christopher R. Houchens, Karen Martins, Lakshmi Jayashankar, Flora Castellino, Obrimpong Amoa-Awua, Manjula Basappa, Britta Flach, Bob C. Lin, Christopher Moore, Mursal Naisan, Muhammed Naqvi, Sandeep Narpala, Sarah O’Connell, Allen Mueller, Leo Serebryannyy, Mike Castro, Jennifer Wang, Christos J. Petropoulos, Alex Luedtke, Ollivier Hyrien, Yiwen Lu, Chenchen Yu, Bhavesh Borate, Lars W. P. van der Laan, Nima S. Hejazi, Avi Kenny, Marco Carone, Daniel N. Wolfe, Jerald Sadoff, Glenda E. Gray, Beatriz Grinsztejn, Paul A. Goepfert, Susan J. Little, Leonardo Paiva de Sousa, Rebone Maboa, April K. Randhawa, Michele P. Andrasik, Jenny Hendriks, Carla Truyers, Frank Struyf, Hanneke Schuitemaker, Macaya Douoguih, James G. Kublin, Lawrence Corey, Kathleen M. Neuzil, Lindsay N. Carpp, Dean Follmann, Peter B. Gilbert, Richard A. Koup, Ruben O. Donis, the Immune Assays Team, the Janssen Team, the Coronavirus Vaccine Prevention Network (CoVPN)/ENSEMBLE Team, the United States Government (USG)/CoVPN Biostatistics Team

## Abstract

Anti-spike IgG binding antibody, anti-receptor binding domain IgG antibody, and pseudovirus neutralizing antibody measurements four weeks post-vaccination were assessed as correlates of risk of moderate to severe-critical COVID-19 outcomes through 83 days post-vaccination and as correlates of protection following a single dose of Ad26.COV2.S COVID-19 vaccine in the placebo-controlled phase of ENSEMBLE, an international, randomized efficacy trial. Each marker had evidence as a correlate of risk and of protection, with strongest evidence for 50% inhibitory dilution (ID50) neutralizing antibody titer. The outcome hazard ratio was 0.49 (95% confidence interval 0.29, 0.81; p=0.006) per 10-fold increase in ID50; vaccine efficacy was 60% (43, 72%) at nonquantifiable ID50 (< 2.7 IU50/ml) and rose to 89% (78, 96%) at ID50 = 96.3 IU50/ml. Comparison of the vaccine efficacy by ID50 titer curves for ENSEMBLE-US, the COVE trial of the mRNA-1273 vaccine, and the COV002-UK trial of the AZD1222 vaccine supported consistency of the ID50 titer correlate of protection across trials and vaccine types.

## Introduction

The ENSEMBLE trial (NCT04505722) was conducted in Argentina, Brazil, Chile, Colombia, Mexico, Peru, South Africa, and the United States to test the ability of a single dose of the replication-incompetent human adenovirus type 26 (Ad26)-vectored Ad26.COV2.S vaccine vs. placebo to prevent moderate to severe-critical COVID-19.^1,2^ Estimated vaccine efficacy against COVID-19 with onset at least 28 days post-injection was 66.1% (95% confidence interval (CI): 55.0 to 74.8) in the primary analysis (median follow-up two months).^1^ The US Food and Drug Administration (FDA) granted an Emergency Use Authorization to the Ad26.COV2.S vaccine as a single primary vaccination dose for individuals aged ≥18 years and, more recently, as a single homologous or heterologous booster dose for individuals aged ≥18 years.^3^ The Ad26.COV2.S vaccine has also been issued an Emergency Use Listing by the World Health Organization,^4^ authorized by the European Commission,^5^ and approved or authorized in more than 100 countries.^6^

There is a need to develop and validate an immune biomarker that correlates with protection^7-9^ (a “correlate of protection,” or CoP) for several applications including aiding approval of demonstrated-effective vaccines for populations underrepresented in the phase 3 trials (e.g. young children^10,11^), aiding approval of refined versions of demonstrated-effective vaccines (e.g., strain or schedule changes), aiding approval of new candidate vaccines that face formidable challenges to directly establish efficacy in phase 3 trials, and providing a study endpoint in early-phase trials for comparison and down-selection of candidate next-generation vaccines.

For most licensed vaccines against viral diseases where a CoP has been established, the CoP is either binding antibodies (bAbs) or neutralizing antibodies (nAbs).^8^ A growing body of evidence supports such immune markers as CoPs for COVID-19 vaccines. First, both bAbs^12^ and nAbs^13^ acquired through infection have been shown to correlate with protection from reinfection, and adoptive transfer of purified convalescent immunoglobulin G (IgG) protected rhesus macaques from SARS-CoV-2 challenge.^14^ Second, nAb titers elicited by DNA,^15^ mRNA,^16^ and adenovirus vectored^17^ COVID-19 vaccines all correlated with protection of rhesus macaques from SARS-CoV-2 challenge. Third, passive immunization of nAbs has demonstrated protective efficacy in a phase 3 trial of high risk individuals.^18^ Fourth, bAbs and nAbs correlated with vaccine efficacy in meta-analyses of phase 3 randomized, placebo-controlled clinical trials.^19,20^ The evidence provided by correlates analyses of such randomized phase 3 trials carry extra weight in the evaluation of CoPs, as the gold standard for obtaining reliable and unbiased evidence.^21^

To this end, the US Government (USG) COVID-19 Response Team in public-private partnerships with the vaccine developers designed and implemented five harmonized phase 3 COVID-19 vaccine efficacy trials with a major objective being to develop a CoP based on an IgG bAb or nAb assay.^22^ The first correlates analysis in this program evaluated the mRNA-1273 COVID-19 vaccine in the COVE trial,^23^ which showed that both IgG bAb and nAb markers measured four weeks post second dose were strongly correlated with the level of mRNA-1273 vaccine efficacy against symptomatic COVID-19, with nAb titer mediating about two-thirds of the vaccine efficacy.^24^ These findings are consistent with those of the phase 3 COV002-UK trial of the AZD12222 (ChAdOx1 nCoV-19) vaccine, where vaccine efficacy against symptomatic COVID-19 increased with post-injection bAb and nAb markers.^25^

The ENSEMBLE trial is included in this USG-coordinated effort to identify CoPs. Using the same approach as used for COVE,^24^ for one dose of the Ad26.COV2.S vaccine in ENSEMBLE we assessed IgG bAb and nAb markers measured four weeks post one dose of the Ad26.COV2.S vaccine in ENSEMBLE as correlates of risk of COVID-19 and as correlates of protection against COVID-19. Three markers were studied: IgG bAbs against SARS-CoV-2 spike protein (spike IgG), IgG bAbs against the spike protein receptor binding domain (RBD IgG), and neutralizing antibodies measured by a pseudovirus neutralization assay (50% inhibitory dilution, ID50). We report spike IgG and RBD IgG readouts in WHO international units (IU) and calibrated ID50 titers to a WHO international standard, which enables comparing the results to those of the COVE and the COV002-UK trials.

## Results

### Immunogenicity subcohort and case-cohort set

The assessment of immune correlates was based on measurement of the antibody markers at D29 (hereafter, “D29” denotes the Day 29 study visit, with an allowable visit window of +/-three days around 28 days post-injection) in the case-cohort set, comprised of a stratified random sample of the study cohort (the “immunogenicity subcohort”) plus all vaccine recipients with the COVID-19 primary endpoint after D29 (“breakthrough cases”) (Extended Data Figure 1A). (Detailed information on the sampling design is in the Statistical Analysis Plan, provided as Supplementary Material.) Extended Data Figure 1B-1D describe the case-cohort set overall and by the three geographic regions Latin America (Argentina, Brazil, Chile, Colombia, Mexico, and Peru), South Africa, and United States, with antibody data available from 48, 15, and 29 breakthrough cases, respectively, and from 212, 200, and 409 non-cases, respectively. All analyses of D29 antibody markers restricted to per-protocol, baseline SARS-CoV-2 seronegative participants in the case-cohort set (Supplementary Table 1, Extended Data Figure 2).

### Participant demographics

The demographics and clinical characteristics of the immunogenicity subcohort (N=826 in the vaccine group, N=90 in the placebo group) are shown in Supplementary Table 2. Of all participants in the immunogenicity subcohort, 50.4% were ≥ 60 years old, 51.7% were considered at-risk for severe COVID-19 (defined as having one or more comorbidities associated with elevated risk of severe COVID-19^1^), and 44.8% had been assigned female sex at birth. At U.S. sites 49.3% had minority status (defined as other than White Non-Hispanic). The immunogenicity subcohort was 26.0% Latin America, 23.9% South Africa, and 50.0% United States. Supplementary Tables 3-5 provide demographics and clinical characteristics of the immunogenicity subcohort by geographic region.

### COVID-19 endpoint

Correlates analyses were performed based on adjudicated moderate to severe-critical COVID-19 with onset that was both ≥ 28 days post-vaccination <u>and</u> ≥ 1 day post-D29, through to January 22, 2021, the data cut date of the primary analysis.^1^ This COVID-19 endpoint was selected to be as close as possible to the COVID-19 endpoint used in the primary analysis^1^ (efficacies against the primary^1^ vs. correlates analysis “moderate to severe-critical COVID-19” endpoints were very similar), while also seeking inclusiveness of endpoints to aid statistical precision. See Online Methods for details on the analysis databases and exact differences between the two endpoints. The last COVID-19 endpoint included in the correlates analysis occurred 48 days post-D29 (Extended Data Figure 1E). Of the 92 breakthrough cases with antibody data, 7 were severe-critical (using the same definition as in ref.^1^), precluding correlates analyses restricted to severe-critical endpoints. Non-cases were defined as baseline seronegative per-protocol participants sampled into the immunogenicity subcohort with no evidence of SARS-CoV-2 infection up to the end of the correlates study period, which is up to 54 days post-D29, the last day such that at least 15 such vaccine recipients were still at risk in the immunogenicity subcohort, but no later than the data cut of January 22, 2021.

### SARS-CoV-2 lineages causing COVID-19 endpoints

Figure 1 in ref.^2^ (which reports the results of the final efficacy analysis) shows the distribution of SARS-CoV-2 lineages among COVID-19 endpoint cases for each country in the trial over time during the double-blind period of the trial (September 21, 2020 through July 9, 2021). Data in this figure through January 22, 2021 are relevant for the current work. With “reference” referring to the Wuhan-Hu-1 strain harboring the D614G point mutation and “other” referring to sequences with substitutions departing from reference not resulting in another SARS-CoV-2 lineage or variant, the results show two lineages in the US, at approximately equal prevalence (reference, other); almost all lineages beta in South Africa; and lineages reference, zeta, and other in Latin America in similar proportions. For the US most “other” lineages were close genetically to reference. These data are consistent with the preliminary sequencing data provided in ref.^1^

**Figure 1.**
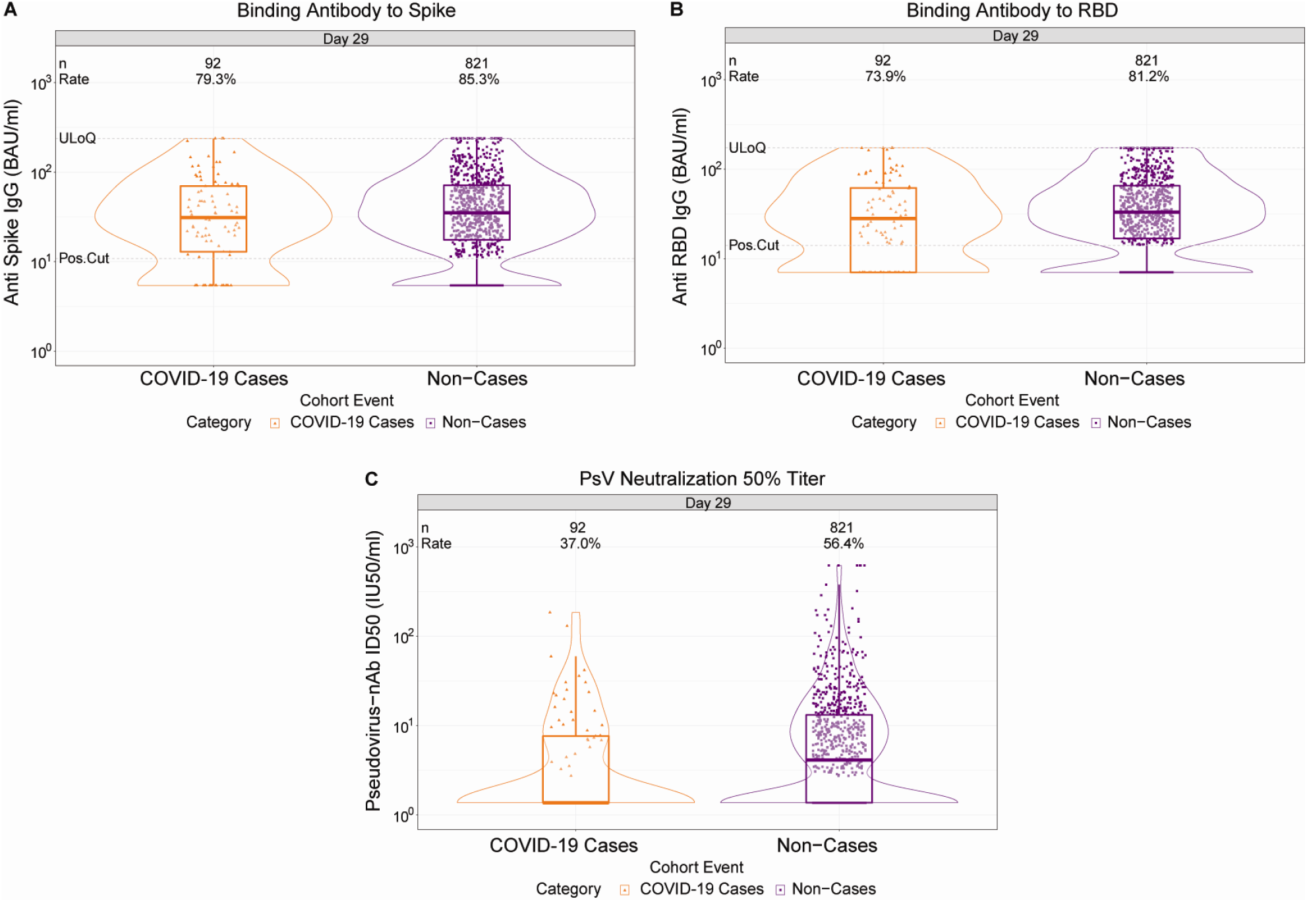
D29 antibody marker level by COVID-19 outcome status. (A) Anti-spike IgG concentration, (B) anti-receptor binding domain (RBD) IgG concentration, and (C) pseudovirus (PsV) neutralization ID50 titer. Data points are from baseline SARS-CoV-2 seronegative per-protocol vaccine recipients in the set. The violin plots contain interior box plots with upper and lower horizontal edges the 25^th^ and 75^th^ percentiles of antibody level and middle line the 50^th^ percentile, and vertical bars the distance from the 25^th^ (or 75^th^) percentile of antibody level and the minimum (or maximum) antibody level within the 25^th^ (or 75^th^) percentile of antibody level minus (or plus) 1.5 times the interquartile range. At both sides of the box, a rotated probability density curve estimated by a kernel density estimator with a default Gaussian kernel is plotted. Positive response rates were computed with inverse probability of sampling weighting. Pos.Cut, Positivity cut-off. Positive response for spike IgG was defined by IgG > 10.8424 BAU/ml and for RBD IgG was defined by IgG > 14.0858 BAU/ml. ULoQ, upper limit of quantitation. ULoQ = 238.1165 BAU/ml for spike IgG and 172.5755 BAU/ml for RBD IgG. LLoQ, lower limit of quantitation. Positive response for ID50 was defined by value > LLoQ (2.7426 IU50/ml). ULoQ = 619.3052 IU50/ml for ID50. Cases are baseline SARS-CoV-2 seronegative per-protocol vaccine recipients with the primary COVID-19 endpoint (moderate to severe-critical COVID-19 with onset both ≥ 1 day post D29 and ≥ 28 days post-vaccination) up to 54 days post D29 but no later than January 22, 2021.

### Vaccine recipient non-cases had higher D29 antibody marker levels than vaccine breakthrough cases

At D29, 85.3% (95% CI: 82.0%, 88.0%) and 81.2% (77.7%, 84.3%) of vaccine recipient non-cases had a positive spike IgG or RBD IgG response, respectively, whereas 56.4% (52.1%, 60.6%) had quantifiable ID50 nAb titer (Figure 1, Table 1). For each D29 marker, the response rate was lower in cases than in non-cases; this difference was largest for ID50 [response rate difference: -19.5% (95% CI: -29.7%, -8.2%)] (Table 1). For each D29 marker, the geometric mean value was also lower in cases than in non-cases, with ID50 again having the greatest difference [3.22 IU50/ml (95% CI: 2.50, 4.15) in cases vs. 4.95 (4.42, 5.55) in non-cases, ratio = 0.65 (0.52, 0.81)]. The bAb markers had slightly higher case/non-case geometric ratios, with 95% CI upper bounds close to 1. Similar results were seen in each ENSEMBLE geographic region (Supplementary Table 6, Extended Data Figures 3-5), with D29 ID50 nAb titer in United States participants having the greatest response rate difference [cases minus non-cases; -26.8% (−41.6%, -6.3%)] and the lowest geometric mean ratio [cases/non-cases; 0.55 (0.41, 0.72)] across all markers and geographic regions.

**Table 1.** D29 antibody marker response rates and geometric means by COVID-19 outcome status. Analysis based on baseline SARS-CoV-2 seronegative per-protocol vaccine recipients in the case-cohort set. Median (interquartile range) days from vaccination to D29 was 29 (2).

| D29 Marker | COVID-19 Cases<sup>1</sup> |  |  | Non-Cases in Immunogenicity Subcohort<sup>2</sup> |  |  | Comparison |  |
| --- | --- | --- | --- | --- | --- | --- | --- | --- |
|  | N | Positive Response Rate (95% CI) | Geometric Mean (GM) (95% CI) | N | Positive Response Rate (95% CI) | Geometric Mean (GM) (95 % CI) | Response Rate Difference (Cases – Non-Cases) | Ratio of GM (Cases/Non-Cases) |
| Anti Spike IgG (BAU/ml) | 92 | 79.3% (69.7%, 86.5%) | 28.98 (23.09, 36.39) | 821 | 85.3% (82.0%, 88.0%) | 33.96 (31.04, 37.16) | -5.9% (-16%, 1.9%) | 0.85 (0.71, 1.02) |
| Anti RBD IgG (BAU/ml) | 92 | 73.9% (63.9%, 82.0%) | 27.54 (22.32, 33.97) | 821 | 81.2% (77.7%, 84.3%) | 32.49 (29.95, 35.26) | -7.3% (-17.8%, 1.5%) | 0.85 (0.71, 1.01) |
| Pseudovirus-nAb ID50 (IU50/ml) | 92 | 37.0% (27.6%, 47.4%) | 3.22 (2.50, 4.15) | 821 | 56.4% (52.1%, 60.6%) | 4.95 (4.42, 5.55) | -19.5% (-29.7%, -8.2%) | 0.65 (0.52, 0.81) |

<sup>1</sup>Cases are baseline SARS-CoV-2 seronegative per-protocol vaccine recipients with the primary COVID-19 endpoint (moderate to severe-critical COVID-19 with onset that was both $\geq 28$ days post-vaccination and $\geq 1$ day post-D29) up to 54 days post D29, but no later than the data cut (January 22, 2021).
<sup>2</sup>Non-cases/Controls are baseline seronegative per-protocol vaccine recipients sampled into the immunogenicity subcohort with no evidence of SARS-CoV-2 infection up to the end of the correlates study period, which is up to 54 days post D29 but no later than the data cut (January 22, 2021). See Extended Data Figure 2. CI, confidence interval; GM, geometric mean.
RBD, receptor-binding domain.

The D29 bAb markers were highly correlated with each other (Spearman rank r = 0.91), whereas they were only moderately correlated with ID50 (r = 0.55 for spike IgG and ID50; r = 0.54 for RBD IgG and ID50) (Extended Data Figure 6). For each D29 marker, the reverse cumulative distribution function curve in the context of the overall vaccine efficacy estimate is shown in Supplementary Figure 1.

As expected because the population is baseline seronegative, D29 positive response rates were near zero in placebo recipients (e.g., for ID50, 0.6% and 0% for cases and non-cases, respectively) (Extended Data Figure 7).

### Each D29 antibody marker is an inverse correlate of risk in vaccine recipients

The cumulative incidence of COVID-19 for vaccine recipient subgroups defined by D29 antibody marker tertile (Figure 2A-C) show that COVID-19 risk decreased with increasing tertile. The hazard ratio (High vs. Low tertile) was 0.75 (95% CI: 0.42, 1.32) for spike IgG, 0.61 (0.34, 1.09) for RBD IgG, and 0.41 (0.22, 0.75) for ID50, with the point estimates and confidence intervals indicating strongest evidence for ID50 as a correlate of risk. Only ID50 passed the pre-specified family-wise error rate (FWER) multiplicity-adjusted p-value threshold for testing whether the hazard rate of COVID-19 differed across the Low, Medium, and High tertiles (Figure 2D; p=0.003, FWER-adjusted p=0.011) (multiplicity adjustment was performed over the six categorical and quantitative markers). Evidence for the spike and RBD bAb markers as inverse correlates of risk across tertiles was weaker, with unadjusted p-values 0.50 and 0.16, respectively (Figure 2D).

**Figure 2.**
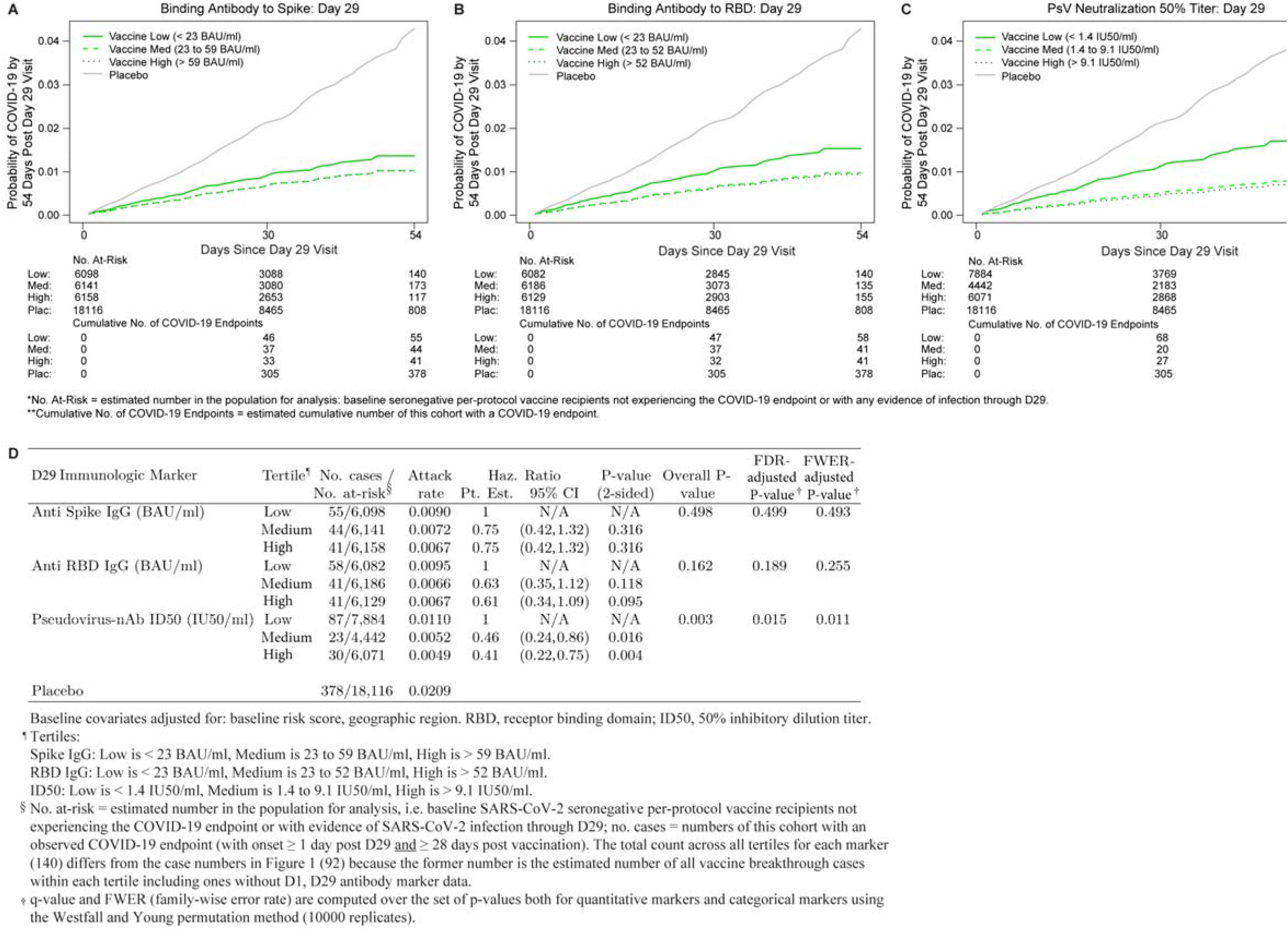
COVID-19 risk by D29 antibody marker level. The plots and table show covariate-adjusted cumulative incidence of COVID-19 by Low, Medium, High tertile of D29 antibody marker level in baseline SARS-CoV-2 seronegative per-protocol participants. (A) Anti-spike IgG concentration; (B) anti-receptor binding domain (RBD) IgG concentration; (C) pseudovirus (PsV) neutralization ID50 titer; (D) each of the markers in (A)-(C). The overall p-value is from a generalized Wald test of whether the hazard rate of COVID-19 differed across the Low, Medium, and High subgroups. Baseline covariates adjusted for were baseline risk score and geographic region.

Similar results were observed for the D29 quantitative markers, with estimated hazard ratio per 10-fold increase in antibody marker level of 0.69 (95% CI: 0.41, 1.16) for spike IgG, 0.59 (0.33, 1.06) for RBD IgG, and 0.49 (0.29, 0.81) for ID50 (Figure 3A). Only ID50 passed the multiple testing correction (FWER-adjusted p=0.016). (Supplementary Table 7 shows the hazard ratios per standard deviation-increase in each D29 marker.) Figure 3B and 3C show analogous results across pre-specified subgroups of vaccine recipients for RBD IgG and ID50, respectively. Similar results were obtained, with nearly all hazard ratios below one.

**Figure 3.**
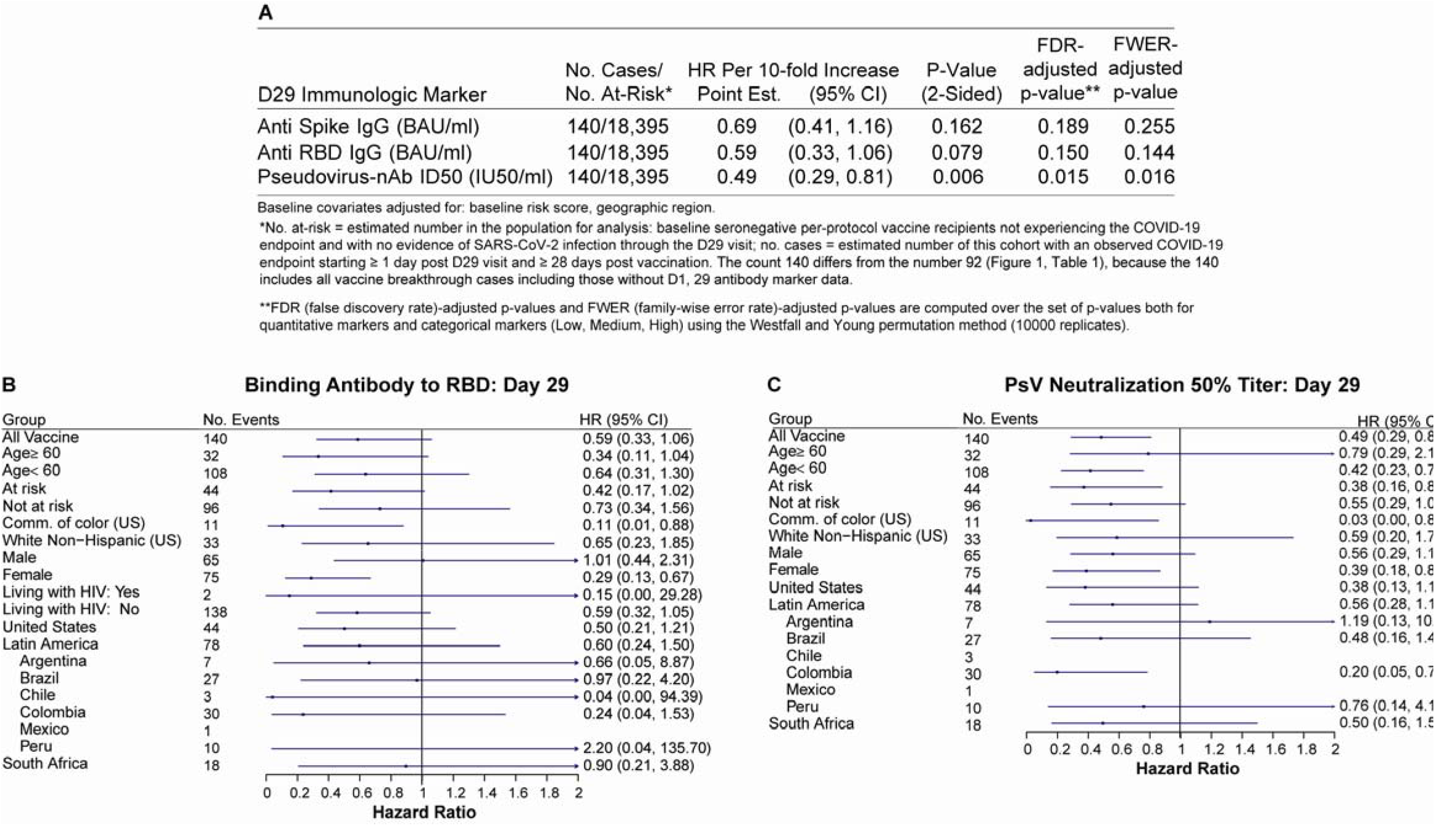
Hazard ratio of COVID-19 as D29 antibody marker level increases. The table and plots show covariate-adjusted hazard ratios of COVID-19 per 10-fold increase in each Day 29 antibody marker in baseline SARS-CoV-2 seronegative per-protocol vaccine recipients (A) overall and (B, C) in subgroups. (A) Inferences for anti-spike IgG concentration, anti-receptor binding domain (RBD) IgG concentration, pseudovirus neutralization ID50 titer; (B, C) Forest plots for (B) anti-RBD IgG concentration; (C) pseudovirus neutralization ID50 titer. Baseline covariates adjusted for were baseline risk score and geographic region.

When vaccine recipients were divided into subgroups defined by having an antibody marker level above a specific threshold and varying the threshold over the range of values, nonparametric regression showed that cumulative incidence of COVID-19 (from 1 to 54 days post-D29) decreased as the ID50 threshold increased (Figure 4A). This decrease in risk was steepest across increasing thresholds closer to the assay lower limit of quantitation (LLOQ = 2.74 IU50/ml) and was more gradual across higher increasing thresholds. For any quantifiable ID50 titer, the risk estimate for COVID-19 was 0.009 (95% CI: 0.007, 0.012), whereas at the highest threshold examined (>185 IU50/ml) the risk estimate for COVID-19 was 0.004 (95% CI: 0.000, 0.009). The bAb markers also showed decreases in risk (although less pronounced) with increasing threshold value (Extended Data Figure 8A, 8B).

**Figure 4.**
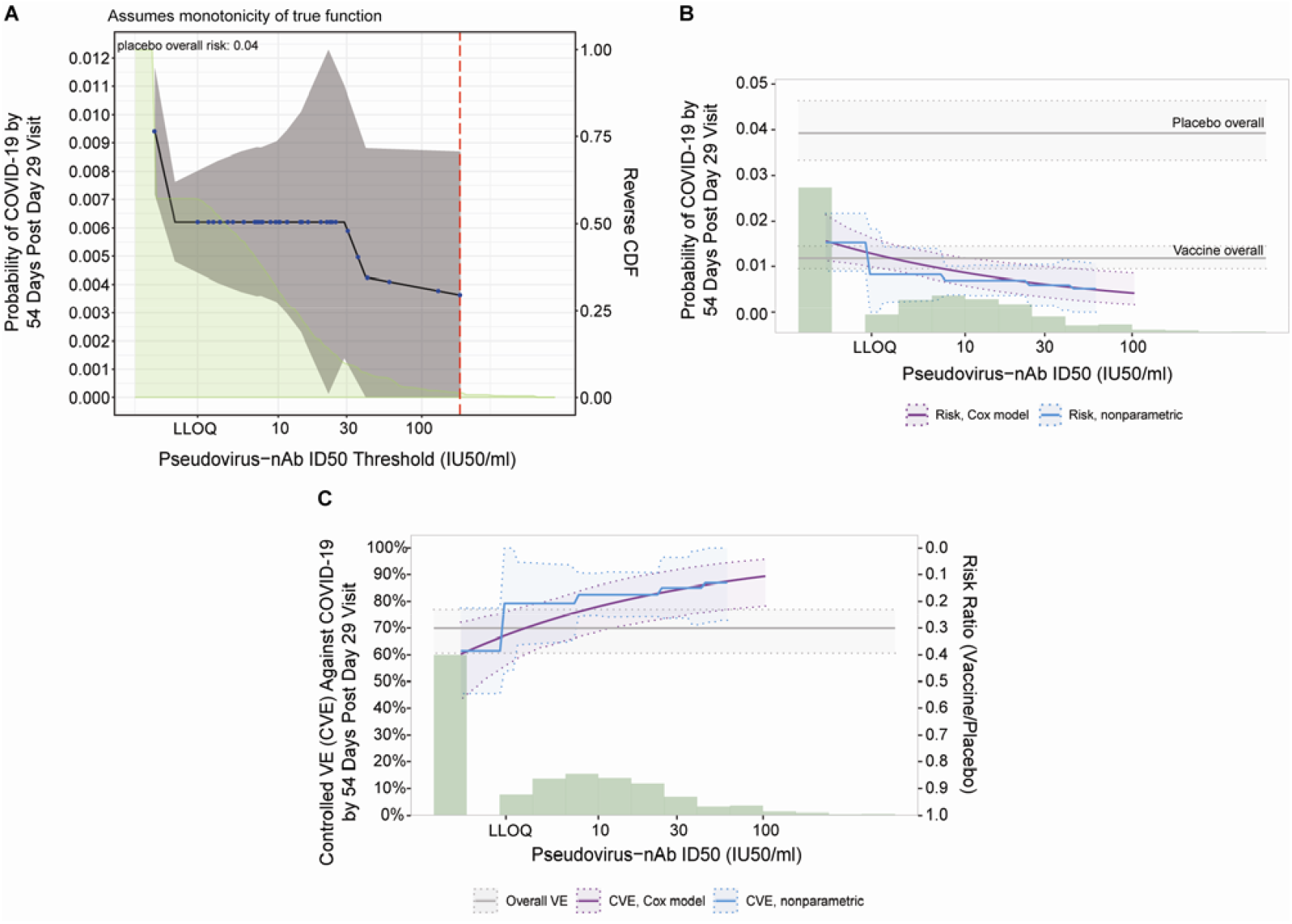
Analyses of D29 ID50 titer as a correlate of risk and as a correlate of protection. Analyses were performed in baseline SARS-CoV-2 seronegative per-protocol vaccine recipients. (A) Covariate-adjusted cumulative incidence of COVID-19 by 54 days post D29 by D29 ID50 titer above a threshold. The blue dots are point estimates at each COVID-19 primary endpoint linearly interpolated by solid black lines; the gray shaded area is pointwise 95% confidence intervals (CIs). The estimates and CIs were adjusted using the assumption that the true threshold-response is nonincreasing. The upper boundary of the green shaded area is the estimate of the reverse cumulative distribution function (CDF) of D29 ID50 titer. The vertical red dashed line is the D29 ID50 threshold above which no COVID-19 endpoints occurred (in the time frame of 1 to 54 days post D29). (B) Covariate-adjusted cumulative incidence of COVID-19 by 54 days post D29 by D29 ID50 titer, estimated using (solid purple line) a Cox model or (solid blue line) a nonparametric method. The dotted black lines indicate bootstrap point-wise 95% CIs. The upper and lower horizontal gray lines are the overall cumulative incidence of COVID-19 from 1 to 54 days post D29 in placebo and vaccine recipients, respectively. (C) Vaccine efficacy (solid purple line) by D29 ID50 titer, estimated using a Cox proportional hazards implementation of ^39^. The dashed black lines indicate bootstrap point-wise 95% CIs. Vaccine efficacy (solid blue line) by Day 29 ID50 titer, estimated using a nonparametric implementation of Gilbert et al.^39^ (described in the SAP). The blue shaded area represents the 95% CIs. In (B) and (C), the green histogram is an estimate of the density of Day 29 ID50 titer and the horizontal gray line is the overall vaccine efficacy from 1 to 54 days post D29, with the dotted gray lines indicating the 95% CIs. Baseline covariates adjusted for were baseline risk score and geographic region. LLOQ, limit of quantitation. In (B, C), curves are plotted over the range from LLOQ/2 to the 97.5^th^ percentile = 96.3 IU50/ml.

Figure 4B and Extended Data Figure 8C, 8D show the Cox modeling results in terms of estimated cumulative incidence of COVID-19 (from 1 to 54 days post-D29) across D29 marker levels. For each antibody marker, COVID-19 risk decreased as antibody marker level increased. Across the full range of D29 ID50 values examined (nonquantifiable ID50 < 2.74 IU50/ml to 96.3 IU50/ml, the 97.5^th^ percentile value), estimated risk decreased from 0.016 (0.011, 0.021) to 0.004 (0.002, 0.008), a 4-fold reduction in risk (Figure 4B). For D29 RBD IgG, estimated risk also decreased across the range of values examined, from 0.016 (0.010, 0.025) at negative response (7 BAU/ml) to 0.008 (0.004, 0.013) at 173 BAU/ml (the 97.5^th^ percentile), a 2-fold reduction in risk (Extended Data Figure 8D). Results for D29 spike IgG were similar (Extended Data Figure 8C).

### Vaccine efficacy increases with D29 antibody marker level

Figure 4C and Extended Data Figure 8E, 8F show estimated vaccine efficacy against COVID-19 (from 1 to 54 days post-D29) across a range of levels of a given D29 antibody marker. For each marker, estimated vaccine efficacy rose with increasing marker level. This increase was greatest for ID50 titer: At nonquantifiable D29 ID50, estimated vaccine efficacy was 60% (95% CI 43, 72%); this increased to 78% (69, 86%) at 9.9 IU50/ml and to 89% (78, 96%) at 96.3 IU50/ml (purple curve, Figure 4C). Nonparametric estimation of the vaccine efficacy-by-D29 ID50 curve suggests that vaccine recipients with unquantifiable ID50 titer had low vaccine efficacy (estimated at 60%) with a jump in vaccine efficacy just above the LLOQ to 79% (blue curve, Figure 4C).

Two sensitivity analyses were performed to evaluate how strong unmeasured confounding would have to be to overturn an inference that D29 antibody marker impacted vaccine efficacy. The first sensitivity analysis, based on E-values,^26^ assessed the robustness of the inference that vaccine efficacy is greater at High vs. Low ID50 tertile (see the SAP for details). The results indicated some robustness to confounding of this inference for ID50 but not for the bAb markers (Supplementary Table 8). The second sensitivity analysis “flattened” the estimated vaccine efficacy-by-D29 antibody marker level curve by assuming a certain amount of unmeasured confounding (see the SAP for details). Estimated vaccine efficacy still increased with D29 ID50 titer in this sensitivity analysis (Extended Data Figure 9).

### Vaccine efficacy increases with D29 ID50 titer within each ENSEMBLE geographic region

Vaccine efficacy increased with D29 ID50 titer in each geographic region (Figure 5A). The US curve was shifted upwards compared to the South Africa curve, which was in turn shifted upwards compared to the Latin America curve. The curves also indicated higher vaccine efficacy at nonquantifiable ID50 in the US (69%; 95% CI: 43, 83%) compared to in South Africa (60%; 16, 82%) and in Latin America (43%; 5, 64%); however, the confidence intervals overlapped. Extended Data Figure 10 shows similar results for spike IgG and for RBD IgG, where vaccine efficacy also increased with D29 bAb marker level (with the exception that vaccine efficacy appeared to remain constant in South Africa with increasing D29 RBD IgG concentration) and the lowest bAb levels were needed in the US out of the three regions to mark a given level of vaccine efficacy. (Participant demographic characteristics of geographic region subgroups of the immunogenicity subcohort are shown in Supplementary Tables 3-5; response rates and magnitudes are shown by case/non-case status, for each geographic region, in Supplementary Table 6 and Extended Data Figures 3-5).

**Figure 5.**
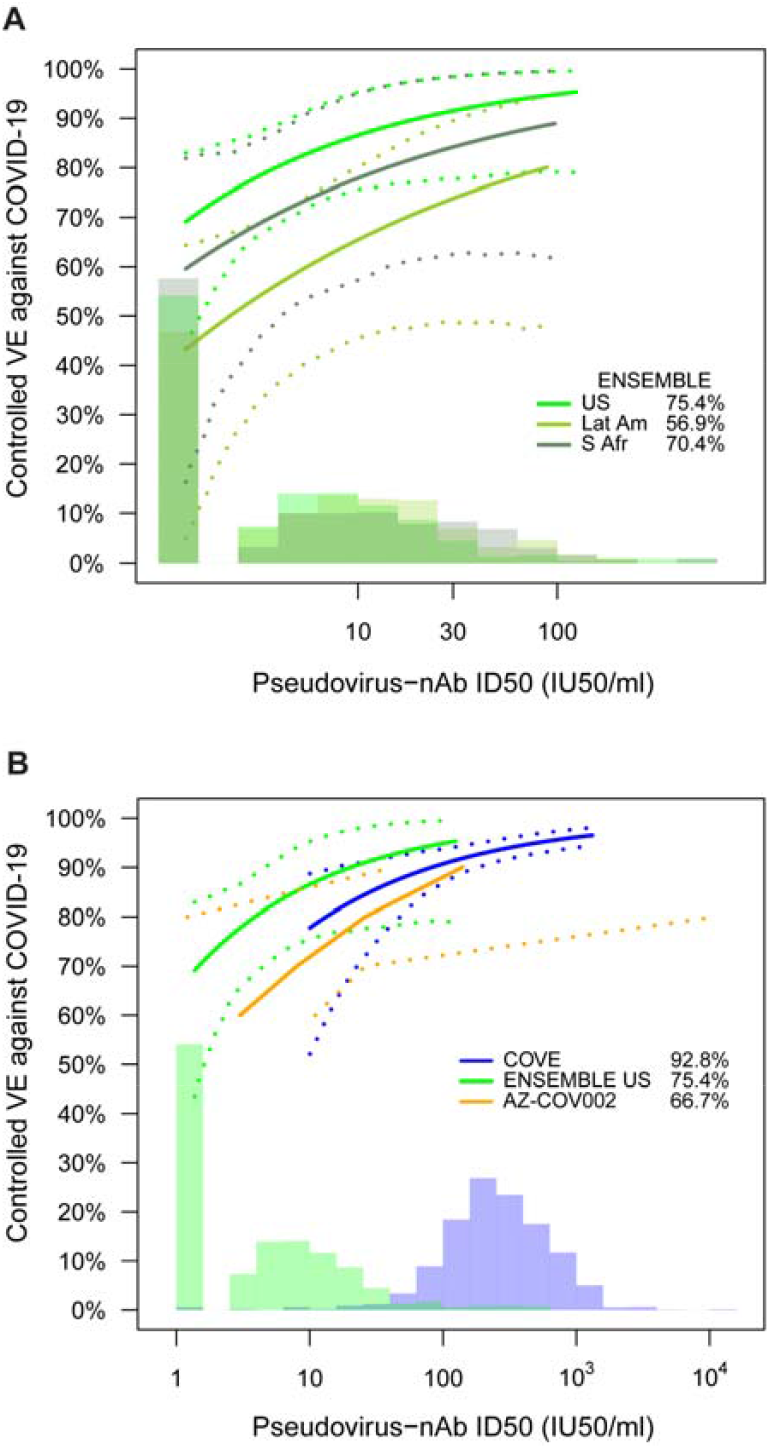
Vaccine efficacy (solid lines) in baseline SARS-CoV-2 seronegative per-protocol vaccine recipients by A) D29 ID50 titer in ENSEMBLE by geographic region (US, United States; Lat Am, Latin America; S Afr, South Africa); B), D57 ID50 titer in COVE, D29 ID50 titer in ENSEMBLE (US), D56 ID50 titer in COV002, all estimated using the Cox proportional hazards implementation of Gilbert et al.^**39**^. The dashed lines indicate bootstrap point-wise 95% CIs. The follow-up periods for the VE assessment were: A) ENSEMBLE-US, 1 to 53 days post D29; ENSEMBLE-Lat Am, 1 to 48 days post D29; ENSEMBLE-S Afr, 1 to 40 days post D29; B) COVE (doses D1, D29), 7 to 100 days post D57; ENSEMBLE-US, 1 to 53 days post D29; COV002 (doses D0, D28; VE defined as 1-relative risk of whether or not an event occurred = 28 days post-D28 till the end of the study period). The green histograms are an estimate of the density of D29 ID50 titer in ENSEMBLE (including by geographic region in A). The blue histograms are an estimate of the density of ID50 titer in baseline SARS-CoV-2 negative per-protocol vaccine recipients in COVE. Curves are plotted over the range from 10 IU50/ml to 97.5^th^ percentile of marker for COVE and from 2.5^th^ percentile to 97.5^th^ percentile for ENSEMBLE. Baseline covariates adjusted for were: ENSEMBLE, baseline risk score and geographic region; COVE: baseline risk score, comorbidity status, and Community of color status; COV002: baseline risk score.

### Vaccine efficacy increases with D29 ID50 titer in a cross-trial, cross-platform comparison

We next compared the vaccine efficacy-by-ID50 titer curves for three double-blind, placebo-controlled COVID-19 vaccine efficacy trials: ENSEMBLE (one dose: D1; VE curve by D29 ID50 titer), COVE (two doses: D1, D29; VE curve by D57 ID50 titer), and the COV002 (United Kingdom) trial^27^ of the AZD1222 (ChAdOx1 nCoV-19) chimpanzee adenoviral-vectored COVID-19 vaccine (two doses: D0, D28; VE curve by D56 ID50 titer). In this comparison for ENSEMBLE we restricted to the US (ENSEMBLE-US) in order to match COVE in its restriction to the US. Direct comparison of vaccine efficacy at a given ID50 titer in ENSEMBLE to vaccine efficacy at the same ID50 titer in COVE is possible because the Duke and Monogram assay readouts underwent concordance testing^24,28^ and were both calibrated to the WHO IS 20/136,^24,28^ expressed in IU50/ml. A similar comparison can be performed vs. the COV002 results, since the same pseudovirus neutralization assay (Monogram) was used in COV002 and ENSEMBLE.^25^

In each trial, vaccine efficacy rose with increasing ID50 titer (Figure 5B). Comparison at high and at low ID50 titers is hindered by the limited overlap of adenovirus-vectored and mRNA vaccine-elicited ID50 titers, with span of values (IU50/ml) from 2.5^th^ to 97.5^th^ percentile 1.4 to 96.3 in ENSEMBLE (the span in ENSEMBLE-US is 1.4 to 98) vs. 32 to 1308 in COVE. In the intersection of these ID50 titer spans (32 to 96.3 IU50/ml), the point estimates of vaccine efficacy are similar and the confidence bands show large overlap. While the confidence intervals of the curves in ENSEMBLE-US are wide, the lower overall vaccine efficacy in ENSEMBLE-US compared to COVE could be explained by the lower ID50 titers, consistent with results of meta-analyses.^20,29^

## Discussion

Each evaluated D29 antibody marker was an inverse correlate of risk of moderate to severe-critical COVID-19 over 83 days post Ad26.COV2.S vaccination, with strongest evidence for ID50 titer, passing the pre-specified multiple testing correction bar. In addition, vaccine efficacy increased with higher D29 antibody marker levels, with results supporting the importance of achieving quantifiable antibodies, as negative binding antibody response and nonquantifiable neutralization corresponded to marginal vaccine efficacy of about 50%. Overall, these findings constitute a step towards establishing an immune marker surrogate endpoint for adenovirus-vectored COVID-19 vaccines, and possibly even a surrogate endpoint that is transportable across vaccine platforms. As discussed further below, it remains to be seen how well each evaluated D29 antibody marker predicts vaccine efficacy against SARS-CoV-2 strains other than those circulating during the trial period, as well as over longer follow-up periods; such data would be important for informing the use of any of these biomarkers as a surrogate endpoint in practice.

Strengths of the study include the fact that all analyses were pre-specified, which increases confidence in the conclusions; the restriction to participants who were SARS-CoV-2 seronegative at enrollment, thus ensuring that the only immune responses studied as correlates are vaccine-elicited ones (i.e. no pre-existing infection-induced immune responses); and the fact that the data come from the double-blind follow-up period of a randomized, placebo-controlled phase 3 vaccine efficacy trial.

The estimated relationship of ID50 titer with vaccine efficacy appeared to differ between the US, Latin America and South Africa, which could be explained by the greater match of the vaccine strain to the reference strain (which predominated in the US) compared to the different strains that circulated in Latin America and South Africa. In support of this hypothesis, Ad26.COV2.S efficacy against moderate to severe-critical COVID-19 with onset ≥ 28 days post-vaccination was reported to be higher against the reference strain [58.2% (95% CI: 35.0%, 73.7%)] than against non-reference lineages [44.4% (34.6%, 52.8%)], particularly against gamma [36.5% (14.1%, 53.3%)], over a median follow-up of 121 days post-vaccination.^2^ Other potential explanations include longer follow-up in the US (perhaps allowing expansion of neutralizing antibody breadth, which is associated with improved coverage of SARS-CoV-2 variants over time^30^) and/or a lower placebo arm attack rate in the US (as greater antibody levels may be needed to protect against greater exposure^8^).

A pillar of the USG-led effort to identify COVID-19 vaccine correlates of protection has been planning and execution of harmonized design and analysis,^22^ to enable cross-trial and meta-analysis correlates assessment. The COVE and ENSEMBLE correlates studies share harmonized trial protocols and study populations, restriction of the analysis to the randomized, placebo-controlled double-blind follow-up period, the same two-phase case-cohort marker sampling design and reproducibly-implemented statistical methods, and common assay readouts measured 4 weeks post-vaccination that were assessed as correlates.^22^ Although the definitions of the primary endpoints differed (COVE: symptomatic COVID-19 of any severity; ENSEMBLE: moderate to severe-critical COVID-19), very few mild cases occurred in ENSEMBLE (1 of 117 cases of symptomatic COVID-19 in the vaccine group, and 3 of 351 cases of symptomatic COVID-19 in the placebo group in the primary efficacy analysis^1^), supporting similarity of the endpoints.

Whereas in ENSEMBLE evidence was strongest for the nAb ID50 marker as a correlate, in COVE the three antibody markers had similar evidence levels as correlates, all passing the pre-specified multiplicity correction bar. Furthermore, similar vaccine efficacy by nAb ID50 curves were observed in ENSEMBLE-US and COVE, and in both trials the vast majority of circulating strains were similar to the reference strain^1,2,31^ (which was used in the nAb assay). This generates the hypothesis that the most transportable correlate across vaccine platforms may involve assessing nAbs against circulating strains, which can be evaluated in the future based on additional data from ENSEMBLE and from other COVID-19 vaccine efficacy trials. Possible explanations for the observed difference in correlate strength for the bAb markers include the different ranges of marker levels, degrees of variation in vaccine efficacy across marker levels (“effect size”), and/or the greater correlations of bAb markers with ID50 in COVE vs.

## ENSEMBLE

We also compare ENSEMBLE results to those of COV002.^25^ In COV002, the bAb markers were strong inverse correlates of risk of symptomatic COVID-19, of similar strength as ID50 nAb titer, similar to COVE. While both the Ad26.COV2.S and AZD1222 vaccines are Ad26-vectored, they also differ (one vs. two doses; pre-fusion stabilized vs. native-like spike; human vs. chimpanzee adenovirus, with differing human receptor usage and reactogenicity profiles between adenovectors). Moreover, different variants [(B.1.177 and B.1.1.7 (alpha)] were circulating at the sites at which COV002 was conducted.^25^

We next discuss some limitations of this study and their implications on future work. First, other Ad26.COV2.S-induced immune responses of interest (e.g. spike-specific T-cell responses,^32^ Fc effector antibody functions^33^) were not assessed. Analyses of D29 spike-specific antibody-dependent cellular phagocytosis (ADCP) are underway; future work will address how ADCP and other immune markers may work together with bAb and/or nAb markers as correlates of protection. A second limitation is the relatively short follow-up (slightly over two months post-D29), which prevented assessment of D29 antibody marker correlates over longer term risk. Measurement of the D29 markers in vaccine breakthrough COVID-19 events occurring after January 22, 2021 (the cut-off of the primary analysis) will enable a future analysis of correlates for COVID-19 through 6-7 months. A third limitation is that the study took place before emergence of the delta and omicron variants (with analysis pooled over all SARS-CoV-2 strains which were mainly reference, beta, zeta, and other^1,2^) and before any boosters were given. Future work is being planned to assess, in the USG trials, levels of post-vaccination nAbs against omicron spike-pseudotyped virus as correlates of risk of omicron COVID-19 (and likewise nAbs against delta spike-pseudotyped virus as delta COVID-19 correlates). The region-specific differences in circulating strains comprise a fourth limitation, in that it is not possible to assess whether strain and/or geographic region has an isolated impact on the correlates of risk and protection. We discuss below our plans for future work to evaluate the dependency of vaccine efficacy on SARS-CoV-2 features and to conduct similar correlates analyses of the COV3009 study of two doses of Ad26.COV2.S. A fifth limitation is that the comparison of vaccine efficacy by antibody marker curves across efficacy trials did not use a common reference covariate distribution in the adjustment for prognostic factors, and the estimates of vaccine efficacy by antibody marker can be biased if a confounder of the effect of the marker on COVID-19 risk was not accounted for.

Given the great interest in assessing correlates against severe COVID-19 and the fact that many Ad26.COV2.S-induced antibody responses show increased magnitude and affinity maturation over time post-D29,^30,34^ the study’s scope of a single clinical endpoint (moderate to severe-critical COVID-19) and a single antibody measurement timepoint (D29) are further limitations. Currently, antibody responses are being assayed in D29 and D71 samples from the remaining ∼300 vaccine breakthrough COVID-19 events during the entire double-blinded period. Planning is underway to assess correlates for COVID-19 over longer-term follow-up, for severe COVID-19, for asymptomatic SARS-CoV-2 infection, and for viral load.

Another important question is how vaccine efficacy depends on SARS-CoV-2 spike features (e.g., amino acid motifs, distances to the vaccine insert, neutralization sensitivity scores), and whether/how the immune correlates depend on these spike features. Future work is planned to address these questions, with the overarching objective being to build a general model for predicting vaccine efficacy across SARS-CoV-2 strains/spike features and time since vaccination, based on D29 and possibly also D71 antibody markers. The data from the additional vaccine breakthrough cases discussed above will provide an opportunity to construct and evaluate such a model, and future work can also evaluate validity of the model specifically for delta and for omicron.

## Online Methods

### Trial design, study cohort, and COVID endpoints

Enrollment for the ENSEMBLE trial began on September 21, 2020. A total of 44,325 participants were randomized (1:1 ratio) to receive a single injection of Ad26.COV2.S or placebo on Day 1. Serum samples were taken on D1 and on D29 for potential antibody measurements. Antibody measurements were evaluated as correlates against the moderate to severe-critical COVID-19 endpoint defined in the main text.

While the correlates analysis only included cases up to Jan 22^nd^, 2021 (the cut-off date of the primary analysis^1^), the correlates analysis was performed using the analysis database of the final analysis.^2^ Compared to the analysis database of the primary analysis, the analysis database of the final analysis includes changes to the SAP and protocol, as well as information that became available only after the database lock date on cases up to Jan 22^nd^, 2021. Specifically, for the primary analysis, the case definition of the moderate to severe-critical COVID-19 endpoint was algorithmically programmed according to the protocol definition (with only severe-critical being assessed by the Case Severity Adjudication Committee). After the primary analysis, the severity was assessed by the (blinded) adjudication committee for all case definitions. This also includes central confirmation results which were obtained after the primary analysis on cases with an onset prior to Jan 22^nd^. Further differences between the moderate to severe-critical COVID-19 endpoint for the correlates analysis vs. that for the primary analysis are: the correlates analysis counted endpoints starting both ≥ 1 day post-D29 and ≥ 28 days post-vaccination and RT-PCR positivity of a nasal swab for SARS-CoV-2 was determined at a local laboratory (with or without central confirmation), whereas the primary analysis counted endpoints starting ≥ 28 days post-vaccination and all participants whose nasal swabs tested RT-PCR+ for SARS-CoV-2 at a local laboratory must have also had a respiratory tract sample confirmed as SARS-CoV-2 positive at a central laboratory using the m-2000 SARS-CoV-2 real-time RT-PCR assay (Abbott).^1^

Correlates analyses were performed in baseline SARS-CoV-2 seronegative participants in the per-protocol cohort, with the same definition of “per-protocol” as in Sadoff et al.^1^ Correlates analyses included COVID-19 endpoints starting both ≥ 1 day post-D29 and ≥ 28 days post-vaccination through January 22, 2021 (excluding cases with any evidence of SARS-CoV-2 infection, such as a positive nucleic acid amplification test or rapid antigen test result, up to D29). Correlates analyses were also done counting endpoints starting seven days after D29 or later through the same data cut, under the rationale that the D29 antibody marker measurements in participants who are diagnosed with the COVID-19 endpoint between 1-6 days post-D29 may possibly be influenced by SARS-CoV-2 infection. The point estimates of both analyses were similar; we report only the results that start counting COVID-19 endpoints at both ≥ 1 day post-D29 <u>and</u> ≥ 28 days post-vaccination, given the greater precision (approximately 35% more vaccine breakthrough cases).

### Solid-phase electrochemiluminescence S-binding IgG immunoassay (ECLIA)

Serum IgG binding antibodies against spike and serum IgG binding antibodies against RBD were quantitated using a validated solid-phase electrochemiluminescence S-binding IgG immunoassay as previously described.^24^ Conversion of arbitrary units/ml (AU/ml) readouts to bAb units/ml (BAU/ml) based on the World Health Organization 20/136 anti SARS-CoV-2 immunoglobulin International Standard^35^ was also as previously described.^24^

### Pseudovirus neutralization assay

Neutralizing antibody activity was measured at Monogram in a formally validated assay (detailed in Huang et al.^28^) that utilized lentiviral particles pseudotyped with full-length SARS-CoV-2 Spike protein. The lentiviral particles also contained a firefly luciferase (Luc) reporter gene, enabling quantitative measurement of infection via relative luminescence units (RLU). Supplementary Table 9 provides the assay limits. Readouts from the Monogram assay have been calibrated to those from the Duke pseudovirus neutralization assay (used in the immune correlates analysis of the COVE trial of the mRNA-1273 vaccine^24^) based on the World Health Organization 20/136 anti SARS-CoV-2 immunoglobulin International Standard^35^ and conversion to International Units/ml (IU50/ml).

### Ethics

All experiments were performed in accordance with the relevant guidelines and regulations. All participants whose serum samples were assayed in this work provided informed consent.

### Statistical methods

All data analyses were pre-specified in the Statistical Analysis Plan (SAP) (available as a supplementary file), except that the total duration of follow-up for studying cumulative incidence of COVID-19 post-D29 was changed from 66 days to 54 days after learning that the marginalized Cox modeling method yielded confidence intervals about the vaccine-efficacy-by-D29-marker-level curve that were wider than they should be based on statistical theory (precipitated by only a few vaccine recipients in the immunogenicity subcohort being at-risk for COVID-19 at 66 days). This problem was solved by switching to a latest time point of 54 days (Section 3.2 of the SAP), and the point estimates of the vaccine-efficacy-by-D29-marker-level curve were very similar for the two choices.

#### Case-cohort set included in the correlates analyses

A case-cohort^36^ sampling design was used to randomly sample participants for D1, D29 antibody marker measurements. This random sample was stratified by the following baseline covariates: randomization arm, baseline SARS-CoV-2 serostatus, and 16 baseline demographic covariate strata defined by all combinations of: underrepresented minority (URM) within the US vs. non-URM within the US vs. Latin America vs. South Africa participant, age 18-59 vs. age ≥ 60, and presence vs. absence of comorbidities (see the SAP for details, as well as Extended Data Figure 2 and Supplementary Table 1).

#### Covariate adjustment

All correlates analyses adjusted for the logit of predicted COVID-19 risk score built from machine learning of data from placebo arm participants (see Supplementary Text 1 and Supplementary Table 10) and geographic region (US, South Africa, Latin America).

#### Correlates of risk in vaccine recipients

All correlates of risk and protection analyses were performed in per-protocol baseline seronegative participants with no evidence of SARS-CoV-2 infection or right-censoring up to D29. For each of the three D29 markers, the covariate-adjusted hazard ratio of COVID-19 (either across marker tertiles or per 10-fold increase in the quantitative marker) was estimated using inverse probability sampling weighted Cox regression models with 95% CIs and Wald-based p-values. These Cox model fits were also used to estimate marker-conditional cumulative incidence of COVID-19 through 54 days post-D29 in per-protocol baseline seronegative vaccine recipients, with 95% CIs computed using the percentile bootstrap. The Cox models were fit using the survey package^37^ for the R language and environment for statistical computing.^38^ The same marker-conditional cumulative incidence of COVID-19 parameter was also estimated using nonparametric dose-response regression with influence-function-based Wald-based 95% CIs.^39^ Point and 95% CI estimates about marker-threshold-conditional cumulative incidence were computed by nonparametric targeted minimum loss-based regression.^40^

#### Correlates of protection

##### Controlled vaccine efficacy

For each marker, vaccine efficacy by marker level was estimated by a causal inference approach using both Cox proportional hazards estimation and nonparametric monotone dose-response estimation.^39^ Two sensitivity analyses of the robustness of results to potential unmeasured confounders of the impact of antibody markers on COVID-19 risk were also conducted, which specified a certain amount of confounding that made it harder to infer a correlate of protection (see the SAP for details). One of the sensitivity analyses was based on E-values^26^ and assessed the robustness of the inference that vaccine efficacy is greater for the upper marker tertile compared to the lower marker tertile. The other sensitivity analysis estimated how much vaccine efficacy increases with quantitative D29 antibody marker despite the specified unmeasured confounder.

#### Hypothesis testing

For hypothesis tests for D29 marker correlates of risk, Westfall-Young multiplicity adjustment^41^ was applied to obtain false-discovery rate adjusted p-values and family-wise error rate (FWER) adjusted p-values. Permutation-based multi-testing adjustment was performed over both the quantitative marker and tertilized marker CoR analyses. All p-values were two-sided.

#### Cross-trial comparisons

Calibration of ID50 nAb titers between the Duke neutralization assay (COVE trial samples) and the Monogram PhenoSense neutralization assay (COV002 and ENSEMBLE trial samples), performed using the WHO Anti-SARS CoV-2 Immunoglobulin International Standard (20/136) and Approach 1 of Huang et al.^28^ (with arithmetic mean as the calibration factor) is described in the supplementary material of Gilbert, Montefiori, McDermott et al.^24^

#### Software and data quality assurance

The analysis was implemented in R version 4.0.3^38^; code was verified using mock data.

## Supporting information

Statistical Analysis Plan

## Data Availability

The complete de-identified patient data set will be made available to others. The data sharing policy of Janssen Pharmaceutical Companies of Johnson & Johnson is available at
https://www.janssen.com/clinical-trials/ transparency. As noted on this site, requests for access to the study data can be submitted through Yale Open Data Access (YODA) Project site at http://yoda.yale.edu.

## Data Availability Statement

The complete de-identified patient data set will be made available to others. The data sharing policy of Janssen Pharmaceutical Companies of Johnson & Johnson is available at https://www.janssen.com/clinical-trials/transparency. As noted on this site, requests for access to the study data can be submitted through Yale Open Data Access (YODA) Project site at http://yoda.yale.edu.

## Code Availability Statement

All analyses were done reproducibly based on publicly available R scripts hosted on the GitHub collaborative programming platform (https://github.com/CoVPN/correlates_reporting2).

## Acknowledgments

This work was partially funded by the Office of the Assistant Secretary for Preparedness and Response, Biomedical Advanced Research and Development Authority, under Government Contract Nos. HHSO100201700018C with Janssen and 75A50122C00008 with Labcorp – Monogram Biosciences; by the National Institutes of Health, National Institute of Allergy and Infectious Diseases (NIAID) under Public Health Service Grants UM1 AI068635 (HVTN SDMC), UM1 AI068614 (HVTN LOC), and R37AI054165; and by the Intramural Research Program of the NIAID Scientific Computing Infrastructure at Fred Hutch, under ORIP grant S10OD028685. This work was also supported by Janssen Research and Development, an affiliate of Janssen Vaccines and Prevention and part of the Janssen pharmaceutical companies of Johnson & Johnson.

The content is solely the responsibility of the authors and does not necessarily represent the official views of the National Institutes of Health. The findings and conclusions in this report are those of the author(s) and do not necessarily represent the views of the Department of Health and Human Services or its components.

