## Supplementary material for "Immune Correlates Analysis of a Single Ad26.COV2.S Dose in the ENSEMBLE COVID-19 Vaccine Efficacy Clinical Trial": Statistical Analysis Plan

April 6, 2022

Version 1.0

### Contents

List of Tables 4

List of Figures 5

1 Introduction 6

2 Antibody Assays and Day 29 Markers 6

3 Study Cohorts and Endpoints 9

10 Baseline Risk Score (Proxy for SARS-CoV-2 Exposure and/or Moderate/Severe COVID-19) 31

**11 Correlates Analysis Descriptive Tables by Case/Non-Case Status** **34**

**12 Correlates of Risk Analysis Plan** **35**

**13 Correlates of Protection: Generalities** **47**

**14 Correlates of Protection: Correlates of Vaccine Efficacy Analysis Plan** **47**

**15 Correlates of Protection: Interventional Effects** **50**

**16 Summary of the Set of CoR and CoP Analyses and Their Requirements and Contingencies, and Synthesis of the Results, Including Reconciling Any Possible Contradictions in Results** **60**
16.1 Synthesis Interpretation of Results . . . 63
16.2 Multiple Hypothesis Testing Adjustment for CoP Analysis . . . 66

**17 CoP: Meta-Analysis Analysis Plan** **67**

**18 Estimating a Threshold of Protection Based on an Established or Putative CoP (Population-Based CoP)** **67**

**19 Considerations for Baseline SARS-CoV-2 Seropositive Study Participants** **68**

**20 Avoiding Bias with Pseudovirus Neutralization Analysis due to Use of Anti-HIV Antiretroviral Drugs** **68**

**21 Accommodating Crossover of Placebo Recipients to the Vaccine Arm** **68**

### 1 Introduction

The statistical analyses specified in this SAP are implemented in R. The R scripts are hosted on the Github code repository
(https://github.com/CoVPN/correlates_reporting2)

### 2 Antibody Assays and Day 29 Markers

The antibody markers of interest are measured using two different humoral immunogenicity assays [more detail on assay type (2) can be found in Sholukh et al. (2020)]:

(1) **bAbs**: **Binding antibodies** to the vaccine insert SARS-CoV-2 proteins;

(2) **Pseudovirus-nAbs**: **Neutralizing antibodies** against viruses **pseudotyped** with the vaccine insert SARS-CoV-2 proteins.

We describe the statistical details needed for data analysis below.

(1) **bAb assay**: The MSD-ECL Multiplex Assay (MSD-ECL = meso scale discovery-electrochemiluminescence assay).

The MSD assay measures binding antibody to antigens corresponding to: Spike (an engineered version of the Spike protein harboring a double proline substitution (S-2P) that stabilizes it in the closed, prefusion conformation [McCallum et al. (2020)]); the Receptor Binding Domain (RBD) of the Spike protein; and Nucleocapsid protein (N), which is not contained in any of the COVID-19 vaccines.

The bAb assay readouts are in units AU/ml, where AU stands for arbitrary units from a standard curve. The process of validating the assay defined a lower limit of detection (LOD), an upper limit of detection (ULOD), a lower limit of quantitation (LLOQ), an upper limit of quantitation (ULOQ), and a positivity cut-off for each antigen that defines positive vs. negative response. These values are as follows:

- bAb Spike:
  - Pos. Cutoff = 1204.71 AU/ml
  - LOD = 34.18 AU/ml
  - ULOD = 19,136,250 AU/ml
  - LLOQ = 199.64 AU/ml
  - ULOQ = 1,128,438.87 AU/ml
- bAb RBD:
  - Pos. Cutoff = 517.86 AU/ml
  - LOD = 58.59 AU/ml
  - ULOD = 8,201,250 AU/ml

- LLOQ = 184.7172 AU/ml
- ULOQ = 598,133.3615 AU/ml

- N:
  - Pos. cutoff = 9779.62 AU/ml
  - LOD = 39.06 AU/ml
  - ULOD = 21,870,000
  - LLOQ = 1870.70 AU/ml
  - ULOQ = 239,449.31

The Vaccine Research Center established factors for converting the MSD assay readouts from AU/ml to WHO International Units/ml. For the three binding antibody variables CoV-2 Spike IgG, CoV-2 RBD IgG, and CoV-2 N IgG, these conversion factors are 0.0090, 0.0272, and 0.0024, respectively. These conversion factors are applied, such that all binding Ab readouts are reported in WHO International Units/ml (BAU/ml), for all analyses. These conversion factors are also applied to yield the LOD, ULOD, LLOQ, and ULOQ on the WHO BAU/ml scale. The following shows the assay limits on the BAU/ml scale:

- bAb Spike:
  - Pos. Cutoff = 10.8424 BAU/ml
  - LOD = 0.3076 BAU/ml
  - ULOD = 172,226.2 BAU/ml
  - LLOQ = 1.8429 BAU/ml
  - ULOQ = 238.1165 BAU/ml
- bAb RBD:
  - Pos. Cutoff = 14.0858 BAU/ml
  - LOD = 1.593648 BAU/ml
  - ULOD = 223,074 BAU/ml
  - LLOQ = 5.0243 BAU/ml
  - ULOQ = 172.5755 BAU/ml
- bAb N:
  - Pos. Cutoff = 23.4711 BAU/ml
  - LOD = 0.093744 BAU/ml
  - ULOD = 52,488 BAU/ml
  - LLOQ = 4.4897 BAU/ml

- ULOQ = 574.6783 BAU/ml

For the three binding antibody markers, all values below the positivity cut-off are assigned the value positivity cut-off divided by 2. For immunogenicity reporting, values greater than the ULOQ are not given a ceiling value of the ULOQ, the actual readouts are used. For the immune correlates analyses, values greater than the ULOQ are assigned the value of the ULOQ.

(2) **Pseudovirus-nAb assay**: A firefly luciferase (ffLuc) reporter neutralization assay for measuring neutralizing antibodies against SARS-CoV-2 Spike-pseudotyped viruses.

Based on the assay in the Monogram lab, serum inhibitory dilution 50% titer (ID50) values are estimated based on a starting serum dilution of 1:40, with a total of ten 3-fold dilutions. Each sample is diluted initially at 1:20, then diluted serially 3-fold for a total of 10 concentrations. The starting dilution of 1:20 is reported as 1:40 after addition of the virus. So, the dilution series is 1:40 to 1:787,320 (= 40 * 39). Thus 1:40 is the LOD on the scale of the assay. The process of validating the assay defined the LOD, LLOQ, and ULOQ for ID50 as follows:

- ID50:
  - LOD = 40
  - LLOQ = 42
  - ULOQ = 9484

ID50 values below the LLOQ are assigned the value LLOQ/2 = 42/2 = 21. For immunogenicity reporting, values greater than the ULOQ are not given a ceiling value of the ULOQ, the actual readouts are used. For the immune correlates analyses, values greater than the ULOQ are assigned the value of the ULOQ.

ID50 values are reported in international units with the following calibration factor, defined using the D614G strain in the assay:

- Calibration factor ID50: 0.0653

The original readouts are calibrated to the IU scale by multiplying each original ID50 value by 0.0653 (See Feng et al. (2021) Table 2 and Gilbert et al. (2022) Supplementary Material), and units are reported in international units as IU50/ml for ID50. Consequently, the LOD, LLOQ and ULOQ for IU50/ml are as follows in International Units:

- IU50/ml:
  - LOD = 2.612
  - LLOQ = 2.7426
  - ULOQ = 619.3052

Based on each immunoassay applied to pairs of serum samples collected from participants on Day 1 (baseline, first dose of vaccination visit) and Day 29 (second dose of vaccination visit), the following set of antibody markers was defined for immunogenicity and immune correlates analyses.

- For bAb: $\log_{10}$ IgG concentration (BAU/ml) at each time point, the difference in $\log_{10}$ concentration (Day 29 minus Day 1) representing $\log_{10}$ fold-rise in IgG concentration from baseline to dose two, and the difference in $\log_{10}$ concentration (Day 29 minus Day 1) representing $\log_{10}$ fold-rise in IgG concentration from baseline to 28 days post dose two. These markers are defined for each antigen Spike, RBD, and N.
- For PsV nAb: $\log_{10}$ serum inhibitory dilution 50% titer (IU50/ml) at each time point, as well as the $\log_{10}$ fold-rise of this marker over Day 1 to Day 29.

### 3 Study Cohorts and Endpoints

#### 3.1 Study Cohort for Correlates Analyses

The primary analysis cohort for correlates analyses is baseline SARS-CoV-2 seronegative participants in the per-protocol cohort, with the per-protocol cohort defined the same as for the primary analysis of vaccine efficacy in the protocol (Sadoff et al., 2021). We refer to this cohort representing the primary population for correlates analysis as the Per-Protocol baseline seronegative (PPBN) Cohort.

Because the primary analysis of vaccine efficacy is in baseline seronegative individuals, CoR and CoP analyses are only done in baseline seronegative individuals, and the analysis of data from baseline seropositive individuals is for purposes of immunogenicity characterization.

In baseline seronegative individuals, antibody marker data in placebo recipients are relevant for verifying the expectation that almost all Day 29 marker responses will be negative, given the lack of SARS-CoV-2 antigen exposure.

#### 3.2 Study Endpoints

Endpoints for per-protocol correlates analyses are included if they occur at least 7 days after the Day 29 visit, to help ensure that the endpoint did not occur prior to Day 29 antibody measurement. Thus participants with a per-protocol endpoint diagnosed up to 6 days post Day 29 visit, or with any evidence of infection up to 6 days post Day 29 visit (e.g., based on a NAAT positive test result or an RT-PCR test result), are excluded from the per-protocol correlates analyses. In addition, the analyses are also done including endpoints starting 1 day after the Day 29 visit, and excluding cases with any evidence of infection by the Day 29 visit. These analyses are justified by their advantage of including more vaccine breakthrough endpoints (about 35% more), which improves precision of correlates analyses, as well as by the expectation that most cases diagnosed 1–6 days post Day 29 visit did not have their Day 29 antibody markers perturbed by a natural infection that occurred before the Day 29 visit. More specifically, it is expected that within the first 5 days of a SARS-CoV-2 infection, the natural infection is unlikely to make antibodies contributing to the antibody marker level, and beyond that time point, the natural infection would increasingly contribute to the antibody level. The analyses that start counting endpoints 7 days or 1 day post Day 29 visit use separate sets of inverse probability of sampling (IPS) weights. Also note that only endpoints occurring at least 28 days post first vaccination are included in correlates analyses, consistent with one of the primary analysis approaches of Sadoff et al. (2021).

Figure 1 defines five study endpoints.

In ENSEMBLE, the primary endpoint is *moderate-to-severe* infection. This is closely related to the COVID infection endpoint in Figure 1, except for the exclusion of the *mild* infections (see study protocol and study SAP for specific details). For consistency between the primary analysis and the correlates analysis, the primary study endpoint will be used here. While the severe COVID endpoint is of paramount clinical importance, the number of events at the time of the first correlates analysis is too small to assess correlates against this endpoint, such that correlates analyses for severe COVID will be done once more endpoints have accrued through longer-term follow-up.

When a correlates analysis is done, all available follow-up for participants is included through to the minimum of (1) the time of the database lock for the correlates analysis and (2) the last day of follow-up post Day 29 visit for which stable inference on marginalized cumulative incidence can be made; call this time the 'administrative censoring time'. This means that the time of right censoring for a given failure time endpoint will be the first event of loss to follow-up or the date of administrative censoring defined as the last date of available follow-up. For CoP analyses, which use both vaccine and placebo recipient data and leverage the randomization, follow-up is censored at the time of unblinding if this date occurs earlier. In general for the first correlates analyses all blinded follow-up is included and no post-unblinding follow-up is included.

For the first correlates analyses, the administrative censoring time is taken to be a participant's earliest event of January 22, 2021 (date of data base lock for selecting samples for the correlates study) and 54 days after the Day 29 study visit. 54 days is chosen to address the fact that the end of follow-up time $t_0$ for defining the marginalized cumulative incidence parameter of interest in the vaccine arm needs to be chosen so that there are a reasonable number of participants at risk in the subcohort at time $t_0$. We choose $t_0$ as the last time such that 15 participants in the subcohort are still at risk, pooling over the three geographic regions, which yields $t_0 = 54$ days.

##### 3.3 Follow-up Included in the Initial Correlates Analyses

The data lock date for inclusion of (Day 1, Day 29) samples for antibody measurement of vaccine breakthrough cases was January 22, 2021. All correlates analyses administratively censor the COVID-19 failure time variable by the calendar date January 22, 2021. Therefore only vaccine breakthrough cases occurring by January 22, 2021 are included in the analysis.

#### 4 Objectives of Immune Correlates Analyses of a Phase 3 Trial Data Set

##### 4.1 Characterize Vaccine Immunogenicity

There are two objectives to characterize the binding and neutralizing antibody immunogenicity of the vaccine:

Stage 1 To characterize vaccine immunogenicity (bAb, PsV nAb) at Days 1 and 29

Stage 2 To characterize vaccine immunogenicity/durability (bAb, PsV nAb) over time (Days 1, 29, 71, time of cross-over and Day 546)

##### 4.2 Correlates of Risk and Correlates of Protection

We broadly classify the proposed analyses into two related categories: correlates of risk (CoR) and correlates of protection (CoP) analyses. CoR analyses seek to characterize correlations/associations of markers with future risk of the outcome amongst vaccinated individuals in the study cohort. CoP analyses seek to formally characterize causal relationships among vaccination, antibody markers and the study endpoint, and use data from both vaccine and placebo recipients. Table 1 summarizes these objectives and statistical frameworks that are commonly used to these ends.

The advantage of CoR analyses is that it is possible to obtain definitive answers from the phase 3 data sets, that is one can credibly characterize associations between markers and outcome. The advantage of CoP analyses is that the effects being estimated have interpretation directly in terms of how an antibody marker can be used to reliably predict vaccine efficacy (the criterion for use of a non-validated surrogate endpoint for accelerated approval, Fleming and Powers (2012)). The disadvantage of CoR analyses are that a CoR may fail to be a CoP, for example due to unmeasured confounding, lack of transitivity where a vaccine effect on an antibody marker occurs in different individuals than clinical vaccine efficacy, or off-target effects (VanderWeele, 2013). The disadvantage of CoP analyses is that statistical inferences rely on causal assumptions that cannot be completely verified from the phase 3 data, such that compelling evidence may require multiple phase 3 trials and external evidence on mechanism of protection (e.g., from adoptive transfer or vaccine challenge trials). Our approach presents results for both CoR and CoP analyses, seeking clear exposition of how to interpret results, the assumptions undergirding the validity of the results, and diagnostics of these assumptions and assessment of robustness of findings to violation of assumptions.

We conjecture that an antibody marker could qualify as a non-validated surrogate endpoint (meeting accelerated approval criteria) based on meeting all three conditions: (1) demonstration of a strong and robust CoR with confounding control; (2) external data supporting functionality and connection to a mechanism of protection; and (3) CoP analyses supporting that the biomarker is likely to be a CoP and not only a CoR. Mechanisms of protection as in (2) may be learned through passive antibody transfer studies and vaccine challenge studies in animals and/or humans.

Table 1: Correlates of Risk (CoRs) and Correlates of Protection (CoPs) Objectives for Day 29 Markers

| Objective Type | Objective |
|---|---|
| **CoRs (Risk Prediction Modeling)** | **To assess Day 29 markers as CoRs in vaccine recipients**<br>a. Relative risks of outcome across marker levels<br>b. Absolute risk of outcome across marker levels<br>c. Machine learning risk prediction for multivariable markers |
| **CoP: Correlates of VE** | **To assess Day 29 markers as correlates of VE in vaccine recipients**<br>a. Principal stratification effect modification analysis<br>b. Assesses VE across subgroups of vaccine recipients defined by Day 29 marker level in vaccine recipients |
| **CoP: Controlled Effects on Risk and VE** | **To assess Day 29 markers for how assignment to vaccine and a fixed marker value would alter risk compared to assignment to placebo** |
| **CoP: Stochastic Interventional Effects on Risk and VE** | **To assess Day 29 markers for how stochastic shifts in their distribution would alter mean risk and VE** (Hejazi et al. (2021)) |
| **CoP: Mediators of VE** | **To assess Day 29 markers as mediators of VE**<br>a. Mechanisms of protection via natural direct and indirect effects<br>a. Estimate the proportion of VE mediated by a marker or markers |

##### 4.3 Synthesis of the Phase 3 Correlates Analyses for Decisions

Establishment of an immunologic biomarker for approval/bridging applications is generally not based on pre-fabricated criteria nor a single type of correlates analysis. Therefore, the goal of the correlates analysis is to generate evidence about correlates from many perspectives, and to synthesize the evidence to support certain decisions. Consequently, we believe there is value in assessing all of the types of correlates presented in Table 1 in this trial, given that the analyses address distinct questions. Obtaining a set of results from multiple distinct approaches that provide complementary and coherent support may increase the rigor and robustness of an evidence package supporting potential use of an antibody marker as a validated surrogate (for traditional approval) or as a non-validated surrogate (for accelerated approval) (Fleming and Powers, 2012); these uses of a biomarker are summarized below. However, the assumptions needed for valid inferences are somewhat different across the methods, and some of these assumptions have testable implications; therefore examination of the assumptions may lead to favoring some methods over others, and affect the synthesis and interpretation of results, and moreover if diagnostics support that some necessary assumptions are infeasible then certain analyses will be canceled, as described below.

Section 16 summarizes the approach that is used and the interpretation of the set of multiple correlates of protection methods. Furthermore, depending on the number of study endpoints in the vaccine and placebo arms at the time a trial delivers primary results, some of the Day 29 marker correlates types defined in Table 1 will be evaluable at the first correlates analyses, whereas others will not be evaluable until additional evaluable vaccine breakthrough endpoints have been observed.

As detailed in Table 4, some CoR analyses are done after there are at least 25 evaluable vaccine breakthrough cases, which is considered to be a minimal number to achieve worthwhile precision. On the other hand, the most nonparametric/flexible CoR analyses require more cases, as do the CoP analyses in general, given the need to adjust for all potential confounders in order to fully identify the causal effects parameters of interest and the greater challenge in estimation (compared to CoR analysis) posed by the need to deal with missing potential outcomes.

Finally, we note that meta-analysis of multiple VE trials will provide important empirical support for potentially establishing an immunologic surrogate endpoint, which underscores the necessity of standardizing the VE trials (common study endpoints, common labs and immunoassays, common statistical methods and data analysis).

##### 4.4 Accommodation of Multiple Geographic Regions in the ENSEMBLE Trial

Primary analysis results on vaccine efficacy have been reported overall and for each of three major geographic regions: U.S., Central/South America, South Africa. The immunogenicity analyses will report results separately across the three major geographic regions. The first immune correlates analyses will be conducted pooling over all geographic regions, to maximize statistical power, with the Cox-model based controlled vaccine efficacy analyses repeated for each geographic region separately.

#### 5 Applications of Immune Correlates Analyses: Vaccine Approval Pathways and Standards of Evidence

Suppose that one or more phase 3 trials demonstrates beneficial vaccine efficacy against the primary clinical endpoint (e.g., symptomatic infection, i.e. COVID) meeting pre-specified success criteria, and correlates analyses of Day 29 antibody marker data are conducted based on the clinical data and antibody data from the phase 3 trial(s). These correlates analyses, combined with additional data supporting the role of antibody markers as mechanisms of protection or as surrogates of mechanisms of protection, can buttress two potential applications of an antibody marker (Table 2).

Table 2: Two Potential Vaccine Approval Pathways Based on a Day 29 Antibody Marker Endpoint

| | |
|---|---|
| Traditional Approval | If the marker is scientifically well-established to reliably predict vaccine efficacy, then subsequent efficacy trials may use the marker as the primary endpoint<br>a. Same vaccine for different populations<br>b. Possibly new vaccines in the same class for the same or different populations |
| Accelerated Approval | If the marker is judged reasonably likely to predict vaccine efficacy but not yet scientifically well established, then accelerated approval based on the marker endpoint may be possible (requires verification of beneficial clinical VE in post marketing studies)<br>a. Same vaccine for different populations<br>b. Possibly new vaccines in the same class for the same or different populations |

Fleming and Powers (2012) defined a *validated surrogate* as a marker that is appropriate for use as an outcome measure for traditional approval of a specific class of interventions against a specific disease, when such interventions are deemed safe and have demonstrated strong evidence that risks from off-target effects are acceptable. They also defined a *non-validated surrogate* as a marker appropriate for use as an outcome measure for accelerated approval as one established to be "reasonably likely to predict clinical benefit" for a specific disease setting and class of interventions. These definitions provide two goalposts for immune correlates analyses of COVID-19 VE trials.

Table 3 summarizes one possible set of requirements for a Day 29 antibody marker to be accepted as a *validated surrogate* for a COVID-19 disease endpoint for use in approving COVID-19 vaccines for specific populations (e.g., SARS-CoV-2 seronegative adults) using Fleming and Power's definition. These potential requirements are conjectures provided for conceptualization purposes, and are not based on COVID-19 regulatory guidance documents.

Table 3: Potential Traditional Approval Requirements for a Day 29 Antibody Marker

| Requirements (1.-6. Required) | Endpoints and Evidence Bar |
|---|---|
| 1. Strong evidence for CoR and CoP in vaccine recipients in animal and/or human challenge models | COVID and VL endpoints: Highly significant and predictive and Severe COVID : Point estimates in the right direction and COV-INF, ASYMP-COV-INF: No countervailing evidence[1] |
| 2. Strong evidence that the marker is a mechanistic CoP or tightly correlated with a mechanistic CoP (likely deriving from animal challenge studies of vaccines or passively transferred antibodies) | Study endpoints used in challenge models such as subgenomic SARS-CoV-2 RNA |
| 3. Supportive evidence from natural history studies of CoRs of re-infection in SARS-CoV-2 infected individuals | Same endpoints as in Phase 3 trial (COVID , severe COVID , ASYMP-COV-INF, COV-INF, VL Dx) |
| 4. Phase 3 trial strong evidence as a CoR in vaccine recipients | COVID and $\geq 1$ other endpoint: Highly significant and predictive and Point estimates in the right direction for the other endpoints Require consistent results from multiple trials |
| 5. Phase 3 trial strong supportive evidence as CoP, for at least one CoP type, plus point estimates in the right direction for the other CoP types (consistency of evidence) | COVID Point estimates of association/causal parameters in the right direction for the other endpoints[2] Require consistent results from multiple trials |
| 6. Temporal ordering support for several of the above results, e.g., CoRs and CoPs are stronger for COVID occurrence proximal to vaccination than distal, synchronized with pattern of biomarker waning | COVID , severe COVID , ASYMP-COV-INF, COV-INF, VL Dx |
| 7. Additional support from non-vaccine interventions, e.g., demonstration of a neutralization CoP for a monoclonal Ab | COVID , severe COVID , ASYMP-COV-INF, COV-INF, VL Dx |

[1]Countervailing evidence could be any observations that provide evidence against a CoP, e.g., relative to Bradford-Hill criteria (see Section 16).

[2]Because CoPs can differ by study endpoint Plotkin (2010) and vaccine efficacy can differ by study endpoint, this criterion will not necessarily be important.

A potential goalpost for a *non-validated surrogate* for accelerated approval can be conceptualized as the same as that for traditional approval, with modifications:

- The package of evidence for the seven sources listed in Table 3 may be less stringent quantitatively, and not requiring success on all of the first six categories.
- Source 4 (Phase 3 CoR in vaccine recipients) would need to have strong evidence (highly statistically significant and highly predictive).
- The support for an immune correlate may be more restricted to a given study endpoint.
- It may no longer be required to have replication of results across two or more Phase 3 trials.

It is hypothesized that a single validated assay will yield a validated or non-validated surrogate endpoint, e.g., based on binding antibody IgG concentration or serum IU50/ml titer to viruses pseudotyped with the Spike vaccine insert protein (or live SARS-CoV-2). However, the goalposts could potentially also be met by a synthesis biomarker aggregated from measurements from multiple validated assays if this aggregation substantially improves the correlate (e.g., a co-correlate Plotkin (2010); Plotkin and Gilbert (2018)). However, the preferred approach, for parsimony and practical utility, would be to define a correlate of protection as a single biomarker derived from a single assay.

### 6 Timeline/Sequencing of Correlates Analyses

The correlates analyses are initiated by the availability of (a) a data set defined at or after the primary analysis data set triggered by the accrual of a certain number of primary endpoints (approximately 150); and (b) Day 1, 29 antibody marker data from correlates-eligible COVID primary endpoint cases from at least 25 baseline seronegative vaccine recipients. The latter requirement ensures that there are enough endpoint cases to achieve worthwhile precision for CoR analyses. The HVTN 505 trial serves as a precedent where 25 evaluable vaccine recipient cases provided enough data to reasonably characterize correlates of risk for a preventive candidate HIV vaccine (Janes et al., 2017; Fong et al., 2018; Neidich et al., 2019; Gilbert et al., 2020b). In addition, simulation studies show that correlates analyses at 20 endpoints have notably lower precision.

Table 4 shows the minimum number of baseline seronegative vaccine recipient endpoints evaluable for correlates analyses that are required before conducting the various planned correlates analyses.

Table 4: Minimum Numbers of Evaluable Endpoints in baseline seronegative Vaccine Recipients to Initiate Correlates Analyses

| Correlates Analysis Type | Number |
|---|---|
| **CoRs (Risk Prediction Modeling)** | |
| a. (Semi)parametric models with strongly parametrized associations: | |
| Cox, hinge/threshold logistic regression | 25 |
| b. Flexible parametric models: Generalized additive model | 35 |
| c. Nonparametric thresholds: Donovan et al. (2019)/ | |
| van der Laan et al. (2021) | 35 |
| d. Superlearner estimated optimal surrogate Price et al. (2018) | 35 |
| **CoP: Correlates of VE** | 50 |
| **CoP: Controlled VE** | 50 |
| **CoP: Stochastic Interventional VE** | 50 |
| **CoP: Mediators of VE** | 50 |

The values above are minimal numbers, where it is preferable to base correlates analyses on several hundred vaccine breakthrough cases, for much improved precision of confidence intervals and statistical power of hypothesis testing procedures.

##### 6.1 Timeline of Statistical Analysis Reports

We summarize the plans for analysis reports over the whole period of the study. When the Day 1, 29 antibody data from the immunogenicity subcohort are available, the first immunogenicity report will be produced. When Day 1, 29 antibody data on COVID cases are also available, the first correlates of risk report will be produced, focusing on Stage 1 data only. When there is enough follow-up to measure antibody markers at the later time points, additional immunogenicity and correlates reports will be made, including those that assess outcome-proximal correlates of risk and protection based on Stage 2 data. The initial correlates reports will only include the symptomatic infection/COVID study endpoint; as data sets become available for the other endpoints the reports will add correlates analyses against the secondary endpoints.

#### 7 General Statistical Issues in Immune Correlates Assessment

Throughout this section, we define the asymptomatic infection endpoint as seroconversion without prior occurrence of the COVID endpoint.

**Issue 1: Timing of endpoint definition, accounting for diagnosis at presentation (i.e., date of virological confirmation of symptomatic COVID – COVID diagnosis) or during post-COVID-19 diagnosis follow-up.**

- **COV-INF:** Defined at presentation (if COVID endpoint) or at first positive serotest visit, whichever occurs first
- **COVID:** Defined at presentation/virologic confirmation
- **Asymptomatic infection:** Defined at first positive serotest (without prior COVID endpoint)
- **Non-severe COVID:** Ascertained by post-COVID diagnosis follow-up, where the failure time could be defined by the time of resolution of symptoms
- **Severe COVID:** Occurs at presentation or at any time during post-COVID diagnosis follow-up

At COVID endpoint diagnosis, participants roll over onto a post-diagnosis follow-up track (Figure 2). This is irrelevant for analysis of the first three endpoints listed above, but for the non-severe COVID endpoint and the severe COVID endpoint special considerations are needed for proper correlates analyses. Survival analysis theory typically requires *predictable processes*, such that non-severe COVID and severe COVID would have failure times defined when the classification of the endpoint is known. However, alternatively, the analysis could be simplified by defining the failure time for all three endpoints COVID, severe COVID, and non-severe COVID to be the date of presentation, even though at that time one needs to look into the future to determine whether the COVID endpoint is severe or non-severe. Such an approach could be justified by thinking of the data as a competing risks data structure, where one observes the time to COVID, and each COVID endpoint has an associated binary endpoint "type", severe or non-severe. The analyses will use this simplified approach. A justification of this simplified approach is that severe COVID is a very rare event among vaccine recipients, and it is the fact of having the event that is important, not whether it happened at or 9 days post COVID diagnosis, such that using a more refined failure time would be unlikely to carry additional meaningful information. If greater than 10% of COVID endpoint cases are missing the endpoint type, then methods accounting for missing endpoint types will be used (e.g., Heng et al., 2020).

**Issue 2: Is the endpoint appropriately analyzed using ordinary survival analysis or competing risks survival analysis?**

For this issue, we consider use of a time-to-event method to assess vaccine efficacy. In general, a competing risk of a given endpoint of interest is an endpoint that, once it occurs, precludes the possibility of future occurrence of the other endpoint.

1. COVID is a competing risk for asymptomatic infection
2. Severe COVID is a competing risk for non-severe COVID

Therefore, the asymptomatic infection and non-severe COVID endpoints may be best analyzed by competing risks methods. For example, instead of estimating cumulative incidence $P(T \leq t|A = a)$ for a given randomization arm $A = a$, where $T$ is the time from enrollment until the endpoint, we analyze cumulative incidence $P(T \leq t, J = 1|A = a)$, where $T$ is the time to the first event of $J = 1$ (event of interest) or $J = 2$ (competing event), and cumulative VE($t$) may be assessed using the parameter

$$\text{VE}(t) = 1 - \frac{P(T \leq t, J = 1|A = 1)}{P(T \leq t, J = 1|A = 0)}.$$

In addition, hazard-ratio-based VE may be defined as one minus the cause ($J = 1$)-specific hazard ratio (Prentice et al., 1978; Gilbert, 2000).

It is also worth noting that:

1. Asymptomatic infection is not a competing risk for COVID, because participants experiencing the asymptomatic infection endpoint continue follow-up for the COVID endpoint (such that at asymptomatic infection diagnosis it is not known whether the infection is truly asymptomatic or pre-symptomatic), and it is not certain that seroconversion prevents future COVID (if future knowledge supports this conclusion, then asymptomatic infection could be treated as a competing risk).

2. Non-severe COVID is not a competing risk for severe COVID. At presentation, if the COVID event does not qualify as severe, then post-COVID diagnosis follow-up is required to determine whether the endpoint registers as non-severe or severe. One will only know the endpoint is not severe after post-COVID diagnosis follow-up is completed (symptoms resolve), such that the failure time is not known until the end of post COVID diagnosis follow-up. Therefore, non-severe COVID is not a competing risk for severe COVID, and the severe COVID endpoint can be analyzed using ordinary survival analysis ignoring the non-severe COVID endpoint.

In sum, the COV-INF, COVID, and severe COVID endpoints will be analyzed by ordinary survival analysis methods, whereas the asymptomatic infection and non-severe COVID endpoints will be analyzed using competing risks methods. Moreover, adding nomenclature precision, for the parent infection endpoint, the daughter endpoints COVID and asymptomatic infection are semi-competing risks data (nomenclature in the survival analysis literature), and for the COVID parent endpoint, the daughter endpoints severe COVID and non-severe COVID are semi-competing risks data.

In addition, one non-clinical endpoint may be important for correlates assessment: SARS-CoV-2 viral load at COVID diagnosis (VL Dx) (e.g., measured by nasal swab), or alternatively area under the viral load curve (AUC-VL) from the COVID diagnosis date through to undetectable viral load, or to an alternative threshold indicating low viral load. Viral load endpoints are putative surrogates of disease progression and severity for the individual, and are also putative surrogates for secondary transmission; moreover the quantitative nature of viral load endpoints may afford an opportunity to increase statistical power.

**Issue 3: Coarseness level of the failure time variable**

1. **COVID:** Event time defined in 'continuous time' on the day of virological confirmation.

2. **Asymptomatic infection:** Event time defined only at fixed infrequent visits (e.g., Month 6, 12, 18, 24).

3. **COV-INF:** Event time defined as 'mixed continuous and discrete', equal to the day of virological confirmation (if COVID) and by the first seropositive visit (if asymptomatic infection).

4. **Non-severe COVID:** Event time may be defined in continuous time, as the number of days from enrollment to COVID diagnosis plus the number of additional days until the COVID event is known to be non-severe. However, following the decision made for Issue 1, we simplify and define the event time at COVID diagnosis.

5. **Severe COVID:** Event time may be defined in continuous time, as the number of days from enrollment to COVID diagnosis plus the number of additional days until the COVID event is known to be severe (which may be zero days). However, following the decision made for Issue 1, we simplify and define the event time at COVID diagnosis.

**Issue 4: Binary endpoint vs. failure time endpoint**

In general, in phase 3 trials with prospective follow-up for event occurrence where right-censoring occurs (either due to administrative censoring or loss to follow-up), it can be advantageous to conduct data analysis in a survival analysis paradigm. Many of the correlates analyses are specified as such. However, because the endpoints are rare, and the rate of loss to follow-up is anticipated to be very low, reliable and interpretable answers may be obtained based on simpler methods that use binary endpoints, and deal with loss to follow-up in a cruder way. If retention is very high, such that bias and precision may be minimally impacted by use of a binary endpoint, some of the correlates analyses may use a binary endpoint. In settings with competing risks, such analyses would treat the endpoint as multinomial and utilize methodology accordingly.

In sum, correlates methods are needed that consider time-to-event or binary endpoints, with or without accounting for a competing risk. In addition, the methods need to be able to handle continuous, discrete, and mixed continuous/discrete failure times.

### 8 Case-cohort Sampling Design for Measuring Antibody Markers

Figure 3 illustrates the case-cohort (Prentice, 1986) sampling design that is used for measuring Day 1, 29 antibody markers (and the later time points at a later point in time) in a random sample of trial participants. The random sample is stratified by the key baseline covariates: randomization arm, baseline SARS-CoV-2 serostatus, and 16 baseline demographic covariate strata defined by all combinations of: underrepresented minority (URM) within the U.S. vs. non-URM within the U.S. vs. Latin America vs. South Africa participant, age 18-59 vs. age $\geq$ 60, and presence vs. absence of comorbidities. This results in a total of 64 sampling strata. A U.S. participant is classified as a URM if they are at least one of the following races and ethnicities: Black or African American, Hispanic or Latinos, American Indian or Alaska Native, Native Hawaiian, and/or other Pacific Islander. A participant is classified as a Latin America participant if they enrolled at any site other than a U.S. or South Africa site. For the sake of defining these baseline strata for sampling into the immunogenicity subcohort, if a U.S. participant's race/ethnicity is missing, they are classified as a non-URM (more precisely, they have not reported being a URM).

Because the design uses a stratified random sample instead of the simple random sample proposed by Prentice (1986), the design may also be referred to as a "two-phase sampling design" (Breslow et al., 2009b,a), where "phase one" refers to variables measured in all participants and "phase two" refers to variables only measured in a subset (thus the "case-cohort sample" constitutes the phase-two data).

The case-cohort design enables obtaining marker data (Day 1, 29) for the immunogenicity subcohort during early trial follow-up in real-time batches, thereby accelerating the time until final data set creation and hence data analysis and results on Day 29 marker correlates. The design allows using the same immunogenicity subcohort to assess correlates for multiple endpoints. This makes the design operationally simpler than a case-control sampling design.

#### 8.1 Randomly Sampled Subcohort for the Case-Cohort Design

The immunogenicity subcohort was sampled from the subset of participants in the Full Analysis Set (FAS) cohort used in the primary analysis of vaccine efficacy against the primary endpoint (with the FAS defined as all randomized participants who received the investigational product) for whom all of the following information was available: baseline SARS-CoV-2 status; age, race/ethnicity, and heightened COVID at-risk status; and Day 1 and Day 29 samples collected.

Tables 5 and 6 summarize the planned sizes of the immunogenicity subcohorts for the U.S. as well as for Latin America and South Africa, by baseline factors used to stratify the random sampling. In the U.S. subcohort 8 baseline demographic strata are used; each of the Latin America and South Africa subcohorts includes 4 baseline demographic strata. For U.S. strata, as in all USG COVID-19 Team trials, a 50:50 balance by underrepresented minority status Yes:No is specified. The subcohort sampling is implemented to create representative sampling across the entire period of enrollment. For the sampling into the U.S. region, Minority includes Blacks or African Americans, Hispanics or Latinos, American Indians or Alaska Natives, Native Hawaiians, and other Pacific Islanders. Non-Minority includes all other races with observed race (Asian, Multiracial, White, Other) and observed ethnicity Not Hispanic or Latino. Therefore Unknown and Not reported have missing values for this sampling stratum variable, such that these participants are not eligible for sampling into the immunogenicity subcohort for the U.S. stratum.

Table 5: Planned U.S. immunogenicity subcohort Sample Sizes by Baseline Strata for Antibody Marker Measurement

| | Baseline SARS-CoV-2 Negative[2] | | | | | | | | Baseline SARS-CoV-2 Positive[3] | | | | | | | |
|---|---|---|---|---|---|---|---|---|---|---|---|---|---|---|---|---|
| Bas. Cov. Strata[1] | 1 | 2 | 3 | 4 | 5 | 6 | 7 | 8 | 1 | 2 | 3 | 4 | 5 | 6 | 7 | 8 |
| Vaccine | 58 | 58 | 58 | 58 | 58 | 58 | 58 | 58 | 18 | 18 | 18 | 18 | 18 | 18 | 18 | 18 |
| Placebo | 7 | 7 | 7 | 7 | 7 | 7 | 7 | 7 | 18 | 18 | 18 | 18 | 18 | 18 | 18 | 18 |

[1]This schema specifies 8 baseline covariate strata for stratified sampling in the U.S. immunogenicity subcohort:
1 = underrepresented minority (URM) in U.S., Age 18-59, absence of comorbidities; 2 = URM in U.S., Age 18-59, presence of comorbidities; 3 = URM in U.S., Age $\geq$ 60, absence of comorbidities; 4 = URM in U.S., Age $\geq$ 60, presence of comorbidities; 5 = non-URM in U.S., Age 18-59, absence of comorbidities; 6 = non-URM in U.S., Age 18-59, presence of comorbidities; 7 = non-URM in U.S., Age $\geq$ 60, absence of comorbidities; 8 = non-URM in U.S., Age $\geq$ 60, presence of comorbidities.

[2]The vaccine group baseline seronegative strata are assigned large sample sizes because the correlates of risk analysis focuses on baseline seronegative vaccine recipients. The placebo group baseline seronegative strata
are assigned small sample sizes given the expectation that almost all Day 29 bAb and nAb readouts will be negative/zero given the absence of prior exposure to SARS-CoV-2 antigens.

[3]Equal stratum sizes are assigned for the vaccine and placebo groups in order to compare bAb and nAb responses in previously infected persons, studying potential differences in natural+vaccine-elicited responses vs. natural-elicited responses.

Table 6: Planned Latin America and South Africa immunogenicity subcohort Sample Sizes by Baseline Strata for Antibody Marker Measurement

| | Baseline SARS-CoV-2 Negative[2] | | | | | | | | Baseline SARS-CoV-2 Positive[3] | | | | | | | |
|---|---|---|---|---|---|---|---|---|---|---|---|---|---|---|---|---|
| Bas. Cov. Strata[1] | 1 | 2 | 3 | 4 | 5 | 6 | 7 | 8 | 1 | 2 | 3 | 4 | 5 | 6 | 7 | 8 |
| Vaccine | 58 | 58 | 58 | 58 | 58 | 58 | 58 | 58 | 18 | 18 | 18 | 18 | 18 | 18 | 18 | 18 |
| Placebo | 7 | 7 | 7 | 7 | 7 | 7 | 7 | 7 | 18 | 18 | 18 | 18 | 18 | 18 | 18 | 18 |

[1]This schema specifies 8 baseline covariate strata for stratified sampling in the Latin America and South Africa immunogenicity subcohorts (4 strata per geographic region): 1 = Latin America, Age 18-59, absence of comorbidities; 2 = Latin America, Age 18-59, presence of comorbidities; 3 = Latin America, Age $\geq$ 60, absence of comorbidities; 4 = Latin America, Age $\geq$ 60, presence of comorbidities; 5 = South Africa, Age 18-59, absence of comorbidities; 6 = South Africa, Age 18-59, presence of comorbidities; 7 = South Africa, Age $\geq$ 60, absence of comorbidities; 8 = South Africa, Age $\geq$ 60, presence of comorbidities.

[2]The vaccine group baseline seronegative strata are assigned large sample sizes because the correlates of risk analysis focuses on baseline seronegative vaccine recipients. The placebo group baseline seronegative strata are assigned small sample sizes given the expectation that almost all Day 29 bAb and nAb readouts will be negative/zero given the absence of prior exposure to SARS-CoV-2 antigens.

[3]Equal stratum sizes are assigned for the vaccine and placebo groups in order to compare bAb and nAb responses in previously infected persons, studying potential differences in natural+vaccine-elicited responses vs. natural-elicited responses.

If certain strata do not have enough eligible participants available for sampling, then additional sampling is done from other strata to keep the total immunogenicity subcohort sample size close to 1616. A separate USG COVID-19 Team Antibody Marker sampling plan describes the sampling algorithm employed (available upon request).

#### 8.2 Correlates Objectives Addressed in Two Stages

There are two stages of the correlates analyses. Stage 1 includes antibody marker data from all COVID and COV-INF cases diagnosed through to the last date of: (1) the time that at least 25 evaluable vaccine breakthrough COVID endpoint cases are available for analysis; and (2) the time of a data-cut at or after the primary analysis used to define the data base for the first correlates analysis. Only Day 1, 29 antibody markers, and COVID and COV-INF diagnosis time point antibody markers, are measured in Stage 1. The objectives of Stage 1 correlates analyses focus on Day 29 markers, which are the objectives listed in Table 1. Stage 1 focuses on Day 29 markers because in general validated or non-validated surrogate endpoints for approved vaccines are based on the peak antibody time point, and this approach fits the priority to develop a validated or non-validated surrogate endpoint as rapidly as possible.

Stage 2 includes antibody marker data from all COVID and COV-INF cases diagnosed after the Stage 1 cases through to the end of the trial, including all available sampling time points (6–7 time points). For immunogenicity subcohort participants, the antibody markers at all available time points other than Day 1, 29 are measured for Stage 2 correlates analyses (4–5 additional time points). The Stage 2 clinical endpoint data and antibody marker data enable assessment of longitudinal antibody markers as outcome-proximal correlates of instantaneous endpoint risk and as various types of outcome-proximal correlates of protection.

The Stage 1 immunogenicity subcohort sampling plan is finalized prior to or shortly after study start. The Stage 2 sampling plan is not made until after the results on vaccine efficacy at the primary analysis are known. The Study Oversight Group may modify the scope of the set of samples for immunoassay measurements in Stage 2 based on analysis results. The essential distinguishing mark of Stage 1 vs. Stage 2 is assessment of Day 29 marker correlates that can be done using antibody data only from Day 1, 29 markers vs. assessment of outcome-proximal correlates that requires antibody data longitudinally including at endpoint diagnosis dates.

###### 8.2.1 Prioritize antibody marker measurement at COVID and COV-INF diagnosis sampling time points

Conduct of the immunologic assays on diagnosis date samples for all COVID and all COV-INF endpoint samples is of the highest priority, equal to the priority of conducting the assays on the Day 1, 29 samples.

### 9 Unsupervised Feature Engineering of Antibody Markers (Stage 1: Day 1, 29)

#### 9.1 Descriptive Tables and Graphics

##### 9.1.1 Antibody marker data

Binding antibody titers to full length SARS-CoV-2 Spike protein, to the RBD domain of the Spike protein, and to the Nucleocapsid (N) protein will be measured in all participants in the immunogenicity subcohort (augmented with infected cases). N-specific binding antibody titers are not used for correlates analyses or for graphical reporting; these data are only used for tabular reporting. Binding antibody IgG Spike, IgG RBD, IgG N, as well as fold-rise in these three markers from baseline, are measured at each pre-defined time point. Indicators of 2-fold rise and 4-fold rise in IgG concentration (fold rise [post/pre] $\geq$ 2 and $\geq$ 4, 2FR and 4FR) are measured at each pre-defined post-vaccination timepoint. Binding antibody responders to a given antigen at each pre-defined timepoint are defined as participants with value above the antigen-specific positivity cut-off. Binding antibody IgG 2FR (4FR) at each pre-defined timepoint to a given antigen are defined as participants who had baseline values below the LLOQ with IgG concentration at least 2 times (4 times) above the assay LLOQ, or as participants with baseline values above the LLOQ with at least a 2-fold (4-fold) increase in IgG concentration.

Pseudovirus neutralizing antibody IU50/ml titer, as well as fold-rise in IU50/ml titer from baseline, are measured at each pre-defined time point. Indicators of 2-fold rise and 4-fold rise in IU50/ml titer (fold rise [post/pre] $\geq$ 2 and $\geq$ 4, 2FR and 4FR) are measured at each pre-defined post-vaccination timepoint. Neutralization responders at each pre-defined timepoint are defined as participants who had baseline values below the LLOQ with detectable IU50/ml neutralization titer above the assay LLOQ, or as participants with baseline values above the LLOQ with a 4-fold increase in neutralizing antibody titer. Neutralization 2FR (4FR) at each pre-defined timepoint are defined as participants who had baseline values below the LLOQ with IU50/ml at least 2 times (4 times) above the assay LLOQ, or as participants with baseline values above the LLOQ with at least a 2-fold (4-fold) increase in neutralizing antibody titer.

Note that for defining positive response, 2FR, and 4FR, a reason why values below the LOD are set to half the LOD before calculating the indicator of response, is to ensure that a vaccine recipient that has an unusually low antibody readout at baseline and a post-vaccination value below or near the LOD is not erronesously counted as a responder.

The following list describes the antibody variables that are measured from immunogenicity subcohort and infection case participants. (The pre-defined time points are Day 1 and Day 29.)

1. Individual anti-Spike antibody concentration at each pre-defined time point
2. Individual anti-Spike antibody fold-rise concentration post-vaccination relative to baseline at each pre-defined post-vaccination time point
3. Individual anti-RBD antibody concentration at each pre-defined time point
4. Individual anti-RBD antibody fold-rise post-vaccination relative to baseline at each pre-defined post-vaccination time point
5. Individual anti-N antibody concentration at each pre-defined time point
6. Individual anti-N antibody fold-rise post-vaccination relative to baseline at each pre-defined post-vaccination time point
7. 2-fold-rise and 4-fold rise (fold rise in anti-Spike antibody concentration [post/pre] $\geq 2$ and $\geq 4$, 2FR and 4FR) at each pre-defined post-vaccination time point
8. 2-fold-rise and 4-fold rise (fold rise in anti-RBD antibody concentration [post/pre] $\geq 2$ and $\geq 4$, 2FR and 4FR) at each pre-defined post-vaccination time point
9. 2-fold-rise and 4-fold rise (fold rise in anti-N antibody concentration [post/pre] $\geq 2$ and $\geq 4$, 2FR and 4FR) at each pre-defined post-vaccination time point
10. Individual pseudovirus-nAb IU50/ml value at each pre-defined time point
11. Pseudovirus-nAb responders, at each pre-defined timepoint defined as participants who had baseline values below the LOD with detectable pseudovirus-nAb IU50/ml titers above the assay LLOQ or as participants with baseline values above the LLOQ with a 4-fold increase in pseudovirus-nAb IU50/ml titers

Summaries of the immunogenicity data will be reported in tables. In particular, the tables will include, for each pre-defined post-baseline time point:

1. For each binding antibody marker, the estimated percentage of participants defined as responders, and with concentrations $\geq$ 2x LLOQ or $\geq$ 4 x LLOQ, will be provided with the corresponding 95% CIs using the Clopper-Pearson method

2. For the IU50/ml pseudo-virus neutralization antibody marker, the estimated percentage of participants defined as responders, participants with 2-fold rise (2FR), and participants with 4-fold rise (4FR) will be provided with the corresponding 95% CIs using the Clopper-Pearson method

3. Geometric mean titers (GMTs) and geometric mean concentrations (GMCs) will be summarized along with their 95% CIs using the t-distribution approximation of log-transformed concentrations/titers (for each of the 3 Spike-targeted marker types including bAb and pseudovirus-nAb IU50/ml, as well as for binding Ab to N).

4. Geometric mean titer ratios (GMTRs) or geometric mean concentration ratios (GMCRs) are defined as geometric mean of individual titers/concentration ratios (post-vaccination/pre-vaccination for each injection)

5. GMTRs/GMCRs will be summarized with 95% CI (t-distribution approximation) for any post-baseline values compared to baseline, and post-Day 29 values compared to Day 29

6. The ratios of GMTs/GMCs will be estimated between groups with the two-sided 95% CIs calculated using t-distribution approximation of log-transformed titers/concentrations [the groups compared are vaccine recipient Non-Cases vs. vaccine recipient breakthrough cases starting 7 days post Day 29 (7+day Cases), and vaccine recipient Non-Cases vs. vaccine recipient breakthrough cases starting 1 day post Day 29 (1-6day Cases)].

7. The differences in the responder rates, 2FRs, 4FRs between groups will be computed along with the two-sided 95% CIs by the Wilson-Score method without continuity correction (Newcombe (1998) (the groups for comparison are as described in the previous bullet).

All of the above point and confidence interval estimates will use inverse probability of antibody marker sampling weighting in order that estimates and inferences are for the population from which the whole study cohort was drawn. In two-phase sampling data analysis nomenclature, the "phase 1 ptids" are the per-protocol individuals excluding individuals with a COVID failure event or any other evidence of SARS-CoV-2 infection < 7 days post Day 29 visit. The "phase 2 ptids" are then the subset of these phase 1 ptids in the immunogenicity subcohort with Day 1 and 29 Ab marker data available. Thus, marker data for the COVID endpoint cases outside the subcohort will not be used in immunogenicity analyses; these cases are excluded from immunogenicity analyses. Similarly, for Day 29 marker correlates analyses the "phase 1 ptids" are the per-protocol individuals excluding individuals with a COVID failure event or any other evidence of SARS-CoV-2 infection < 7 days post Day 29. The "phase 2 ptids" are then the subset of these phase 1 ptids in the immunogenicity subcohort with Day 1 and Day 29 Ab marker data available. Thus again, marker data for the COVID endpoint cases outside the subcohort will not be used in immunogenicity analyses; these cases are excluded from immunogenicity analyses.

The estimated weight $\hat{w}_{subcohort.x}$ is the inverse sampling probability weight, calculated as the empirical fraction (No. phase 1 ptids / No. phase 2 ptids) within each of the baseline strata [(vaccine, placebo) × (baseline seronegative, baseline seropositive) × (demographic strata)]. For individuals outside the phase 1 ptids, $\hat{w}_{subcohort.x}$ is assigned the missing value code NA. All other individuals have a positive value for $\hat{w}_{subcohort.x}$, including cases not in the subcohort. This weight is only used for case-blinded immunogenicity inferential analyses. Note that $\hat{w}_{subcohort.x}$ is used for all immunogenicity subcohort immunogenicity analyses, which are based solely on the immunogenicity subcohort, for Day 1 and Day 29 markers. (Not used for correlates analyses.)

Tables will be provided separately for (1) baseline seronegative individuals, (2) baseline seropositive individuals, (3) baseline seronegative individuals by subgroup defined as in Table 7, and (4) baseline seropositive individuals by the same subgroups as in (3). Each table will show data for all available time points and for each of the vaccine and placebo arms.

Table 7 shows subgroups that are analyzed, within each of the four immunogenicity reports (overall, U.S. region, South Africa region, Latin America region).

Table 7: Baseline Subgroups that are Analyzed.[1]

| |
|---|
| **Age:** $18-59$, $\geq 60$ |
| **Country:** U.S., Argentina, Brazil, Chile, Colombia, Mexico, Peru, South Africa |
| **Heightened risk for Severe COVID:** At risk, Not at risk |
| **Age x Risk for Severe COVID:** |
| All 4 combinations of (Age 18-59, Age $\geq$ 60) $\times$ (At risk, Not at risk) |
| **Sex Assigned at Birth:** Male, Female |
| **Age x Sex Assigned at Birth:** |
| All 4 combinations of (Age 18-59, Age $\geq$ 60) $\times$ (Male, Female) |
| **Hispanic or Latino Ethnicity:** Hispanic or Latino, Not Hispanic or Latino |
| **Race or Ethnic Group:** |
| White Non-Hispanic[2], Black, Asian, American Indian or Alaska Native (NatAmer)[3] |
| Native Hawaiian or Other Pacific Islander (PacIsl), Multiracial, |
| Other, Not reported, Unknown |
| **Underrepresented Minority Status in the U.S.:** |
| URM,[4] Non-URM |
| **Age x Underrepresented Minority Status in the U.S.:** |
| All 4 combinations of (Age 18-59, Age $\geq$ 60) $\times$ (URM, Non-URM) |

[1]All analyses are done within strata defined by randomization arm and baseline seropositive/negative status, such that these variables are not listed here as subgroups for analysis.
[2]White Non-Hispanic is defined as Race=White and Ethnicity=Not Hispanic or Latino. All of the other Race subgroups are defined solely by the Race variable, with levels Black, Asian, American Indian or Alaska Native, Native Hawaiian or Other Pacific Islander, Multiracial, Other, Not reported, Unknown. Only the U.S. region reports results by White Non-Hispanic vs. Community of Color. Communities of color is defined by the complement of being known White Non-Hispanic.
[3] For the Latin America region report only, the American Indian or Alaska Native category is labeled as "Indigenous South American."
[4] See beginning of Section 8 for a definition.

For comparing antibody levels between groups, the following groups are compared:

- baseline seronegative vaccine vs. baseline seronegative placebo
- baseline seropositive vaccine vs. baseline seropositive placebo
- baseline seronegative vaccine vs. baseline seropositive vaccine
- Within baseline seronegative vaccine recipients, compare each of the following pairs of subgroups listed in Table 7: Age $\geq$ 60 vs. age 18-59; risk for severe COVID: at risk vs. not at risk; age $\geq$ 60 at risk vs. age $\geq$ 60 not at risk; age 18-59 at risk vs. age 18-59 not at risk; male vs. female; Hispanic or Latino ethnicity: Hispanic or Latino vs. Not Hispanic or Latino; Underrepresented minority status: Communities of color vs. White Non-Hispanic (within the U.S.).

The entire immunogenicity analysis is done in the per-protocol cohort with both Day 1 and Day 29 marker data available (the two-phase sample).

##### 9.1.2 Graphical description of antibody marker data

The Day 1, 29 antibody marker data collected from the immunogenicity subcohort participants will be described graphically. These data are representative of the entire study cohort. Importantly, only antibody data from the immunogenicity subcohort are included (i.e., no data from cases outside the subcohort are included). This makes the analyses unsupervised (independent of case-control status), enabling interrogation and optimization of the antibody biomarkers prior to the inferential correlates analyses.

Plots are developed for the following purposes. All of the analyses are done separately within each of the four subgroups defined by randomization arm cross-classified with baseline seronegative/seropositive status. In addition, many of the descriptive analyses will also be done separately for each demographic subgroup of interest listed above. For descriptive plots of individual marker data points that pool over one or more of the baseline strata subgroups, plots show all observed data points.

For each antibody marker readout, both Day 29 and baseline-subtracted Day 29 readouts are of interest. We will refer to the latter as 'delta.' All readouts, including delta, will be plotted on the $\log_{10}$ scale, with plotting labels on the natural scale. As such, delta is $\log_{10}$ fold-rise in the marker readout from baseline.

The following descriptive graphical analyses are done.

1. The distribution of each antibody marker readout at Day 1 and Day 29 will be described with plots of empirical reverse cumulative distribution functions (rcdfs) and boxplots (including individual data points) within each of the four groups defined by randomization arm (vaccine, placebo) and baseline serostratum (seronegative, seropositive). Inverse probability of sampling into the subcohort weights ($\hat{w}_{subcohort.x}$) are used in the estimation of the rcdf curves; henceforth we refer to these weights as "inverse probability of sampling" (IPS) weights. Analyses of Day 1 markers always pool across vaccine and placebo recipients given that the two subgroups are the same at baseline.

2. Plots are arranged to compare each Day 29 marker readout between randomization arms within each of the baseline seropositive and baseline seronegative subgroups.

3. Plots are also arranged to compare each Day 29 marker readout between baseline serostatus groups within each randomization arm.

4. The correlation of each antibody marker readout between Day 1 and Day 29, and between Day 1 and delta, is examined within each of the baseline strata subgroups, and within each randomization arm and baseline serostratum. Pairs plots/scatterplots will be used, annotated with baseline strata-adjusted Spearman rank correlations, implemented in the PResiduals R package Li and Shepherd (2012) available on CRAN. For calculating the correlation within each randomization arm and baseline serostratum, because PResiduals does not currently handle sampling weights, the correlation esti-

mates are computed as follows: For each re-sampled data set in the second approach to graphical plotting, the covariate-adjusted Spearman correlation is calculated. The average of the estimated correlations across re-sampled data sets is reported.

5. The correlation of each pair of Day 1 antibody marker readouts are compared within each of the baseline demographic subgroups and baseline serostratum, pooling over the two randomization arms. Pairs plots/scatterplots and baseline-strata adjusted Spearman rank correlations are used, with covariate-adjusted Spearman rank correlations computed as described above.

6. The correlation of each pair of Day 29 and delta antibody marker readouts are compared within each of the baseline demographic subgroups, randomization arm, and baseline serostratum. Pairs plots/scatterplots and baseline-strata adjusted Spearman rank correlations are used, with covariate-adjusted Spearman rank correlations computed as described above.

#### 9.2 Methods for Positive Response Calls for bAb and nAb Assays

As noted above, binding antibody responders at each pre-defined timepoint are defined as participants with concentration above the specified positivity cut-off, with a separate cut-off for each antigen Spike, RBD, N (10.8424, 14.0858, and 23.4711, respectively, in BAU/ml). This approach is used for each of the Spike and RBD and N protein antigen targets.

Pseudovirus neutralization responders at each pre-defined timepoint are defined as participants who had baseline IU50/ml values below the LLOQ with detectable IU50/ml neutralization titer above the assay LLOQ, or as participants with baseline values above the LLOQ with a 4-fold increase in neutralizing antibody titer. Otherwise a value is negative for pseudovirus neutralization.

#### 9.3 SARS-CoV-2 Antigen Targets Used for bAb and nAb Markers

The homologous vaccine strain antigens (Wuhan) are used for the immune correlates analyses for the bAb markers, whereas the homologous vaccine strain with D614G mutation (away from the Wuhan WT strain) is used for the pseudovirus nAb markers.

#### 9.4 Score Antibody Markers Combining Information Across Individual bAb and nAb Readouts

Day 29 score antibody markers that combine information across the individual markers are defined and included in the multivariable CoR machine learning analyses, which are not done based on bAb and nAb markers alone but will await additional markers. In particular, five score variables are studied:

1. Maximum signal-diversity score calculated as described in He and Fong (2019)

2. First two linear principal components PCA1 and PCA2

3. Nonlinear extensions of principal components FSDAM1 and FSDAM2 calculated as in Fong and Xu (2020)

The purpose of these score markers is to seek to maximally capture the main immune response signal and to study whether there are more than one distinct signals that are associated with the COVID outcome, and to study whether score markers can provide strengthened association with COVID compared to the individual assay markers. The score markers are included as input features in the machine learning (superlearning) prediction modeling (multivariable CoR objective).

### 10 Baseline Risk Score (Proxy for SARS-CoV-2 Exposure and/or Moderate/Severe COVID-19)

A baseline risk score will be developed based off a pre-specified list of baseline covariates potentially relevant for SARS-CoV-2 exposure and moderate-to-severe infection risk. This baseline risk score will be controlled for in correlates analyses to adjust for potential confounding.

The risk score is developed using placebo arm data only, restricting to baseline seronegative per-protocol placebo recipients. The risk score is defined as the logit of the predicted outcome probability from a regression model estimated using the ensemble algorithm superlearner (i.e. stacking), where this logit predicted outcome is scaled to have empirical mean zero and empirical standard deviation one. The settings of superlearner (i.e., loss function, cross-validation technique, library of learners) that are used for implementation of superlearner for building a baseline risk score are described in Section 12.6.1.

For predictive modeling of the COVID endpoint, cases are COVID endpoints starting 1 day post Day 29 visit and non-cases are participants with follow-up beyond 7 days post Day 29 visit that never registered a COVID endpoint.

Independent of the superlearner risk score, important individual risk factors will also be specified for inclusion as adjustment factors in correlates analyses. In particular, in addition to the risk score, the at-risk indicator and the two geographic region indicator variables are adjusted for in all correlates analyses. This choice is justified by the epidemiological data showing that these variables are strong risk factors for COVID-19 infection, and making use of the flexibility of super learner to develop a model for how age relates to risk.

Henceforth we refer to the baseline variables that are adjusted for in correlates analyses as "baseline factors" which, depending on the risk score results and performance, will consist of either only the individual key risk factors, or key individual risk factors plus the baseline risk score. For the overall analyses including all geographic regions, some of the selected individual risk factors may depend on region, for example for the U.S. likely communities of color status would be included. The selected baseline factors to adjust for are: baseline risk score and geographic region (U.S., Central/South America, South Africa).

For all overall (all region) correlates analyses conducted using methods that specify baseline endpoint hazards, a separate baseline hazard will be used for each of the three geographic regions U.S., Central/South America, and South Africa. All correlates analyses that adjust for baseline factors in other ways, will include geographic region as a covariate to adjust for.

Table 8: List of baseline covariates considered for risk score analysis.

| Variable Name | Definition |
|---|---|
| EthnicityHispanic | Indicator ethnicity = Hispanic (1 = Hispanic, 0 = complement) |
| EthnicityNotreported | Indicator ethnicity = Not reported (1 = Not reported, 0 = complement) |
| Black | Indicator race = Black (1=Black, 0=complement) |
| Asian | Indicator race = Asian (1=Asian, 0=complement) |
| NatAmer | Indicator race = American Indian or Alaska Native<br>(1=NatAmer, 0=complement) |
| Multiracial | Indicator race = Multiracial (1=Multiracial, 0=complement) |
| Notreported | Indicator race = Not reported (1=Notreported, 0=complement) |
| Unknown | Indicator race = unknown (1=Unknown, 0=complement) |
| URMforsubcohortsampling | Indicator of under-represented minority (1=Yes, 0=No) |
| HighRiskInd | Baseline covariate indicating $\geq$ 1 co-existing conditions<br>(1=yes, 0=no, NA=missing) |
| Sex | Sex assigned at birth (1=female, 0=male/undifferentiated/unknown) |
| Age | Age at enrollment in years (integer ¿= 18, NA=missing).<br>Note that the randomization strata included Age 18-59 vs. Age $\geq$ 60. |
| BMI | BMI at enrollment (Ordered categorical 1, 2, 3, 4, NA=missing);<br>1 = Underweight BMI $<$ 18.5; 2 = Normal BMI 18.5 to $<$ 25;<br>3 = Overweight BMI 25 to $<$ 30; 4 = Obese BMI $\geq$ 30 |
| Country.X1 | Indicator country = Argentina (1 = Argentina, 0 = complement) |
| Country.X2 | Indicator country = Brazil (1 = Brazil, 0 = complement) |
| Country.X3 | Indicator country = Chile (1 = Chile, 0 = complement) |
| Country.X4 | Indicator country = Columbia (1 = Columbia, 0 = complement) |
| Country.X5 | Indicator country = Mexico (1 = Mexico, 0 = complement) |
| Country.X6 | Indicator country = Peru (1 = Peru, 0 = complement) |
| Country.X7 | Indicator country = South Africa (1 = South Africa, 0 = complement) |
| Region.X1 | Indicator region = Latin America (1 = Latin America, 0 = complement) |
| Region.X2 | Indicator country = Southern Africa (1 = Southern Africa, 0 = complement) |
| CalDtEnrollIND.X1 | Indicator variable representing enrollment occurring between 4-8 weeks<br>periods of first subject enrolled<br>(1 = Enrollment between 4-8 weeks, 0 = complement). |
| CalDtEnrollIND.X2 | Indicator variable representing enrollment occurring between 8-12 weeks<br>periods of first subject enrolled<br>(1 = Enrollment between 8-12 weeks, 0 = complement). |
| CalDtEnrollIND.X3 | Indicator variable representing enrollment occurring between 12-16 weeks<br>periods of first subject enrolled<br>(1 = Enrollment between 12-16 weeks, 0 = complement). |

[1]Binary input variable/s having $\leq$ 3 cases in the variable = 1 or 0 subgroup will be dropped from analysis.

[2]Input variable having $>$ 5% missing values will be dropped from analysis.

[3]Input variables with $<$ than 5% missing values will be included in analysis upon imputing the missing values using the mice package in R.

### 11 Correlates Analysis Descriptive Tables by Case/Non-Case Status

The key tables summarizing the distribution of each of the Day 29 antibody markers are listed below. For each table, for each time point Day 1, Day 29 separately, the positive response rate with 95% CI, and the GMT or GMC with 95% CI, is reported for each of the case and non-case groups. In addition, the point and 95% CI estimate of the difference in positive response rate (non-cases vs. cases) and the GMT or GMC ratio (non-cases/cases), is reported. Two cases vs. non-cases comparisons are done: 7+day Cases vs. Non-cases, and 1+day Cases vs. Non-cases, with 7+day Cases and 1+day Cases defined below. The same set of Non-cases is used in each comparison.

1. Immunogenicity table: Antibody levels in the baseline SARS-CoV-2 seronegative per-protocol cohort (vaccine recipients). 7+day Cases are baseline seronegative per-protocol vaccine recipients with the symptomatic infection COVID-19 primary endpoint diagnosed starting 7 days after the Day 29 study visit. 1-6day Cases are baseline seronegative per-protocol vaccine recipients with the symptomatic infection COVID-19 primary endpoint diagnosed starting 1 day after the Day 29 study visit and before 7 days post Day 29 study visit. Thus 1+day Cases are the union of both the 7+day Cases and the 1-6day Cases. Non-cases/Controls are baseline seronegative per-protocol vaccine recipients sampled into the immunogenicity subcohort with no evidence of SARS-CoV-2 infection up to the end of the correlates study period, which is up to 54 days post D29 but no later than the data cut (1/22/21).

2. Antibody levels in the baseline SARS-CoV-2 seropositive per-protocol cohort (vaccine recipients). Cases are baseline seropositive per-protocol vaccine recipients with the symptomatic infection COVID-19 primary endpoint diagnosed starting 7 days after the Day 29 study visit. Non-cases/Controls are baseline seropositive per-protocol vaccine recipients sampled into the immunogenicity subcohort with no evidence of SARS-CoV-2 infection up to the end of the correlates study period, which is up to 54 days post D29 but no later than the data cut (1/22/21).

3. Antibody levels in baseline SARS-CoV-2 seropositive placebo recipients. Cases are baseline seropositive per-protocol placebo recipients with the symptomatic infection COVID-19 primary endpoint diagnosed any time after D1 or after the Day 29 visit (by time of antibody measurement). Non-cases/Controls are baseline seropositive per-protocol placebo recipients sampled into the immunogenicity subcohort with no evidence of SARS-CoV-2 infection up to the end of the correlates study period, which is up to 54 days post D29 but no later than the data cut (1/22/21).

4. Repeat Table 2 above for fold-rise from baseline (of interest given the analysis cohort is baseline seropositive).

5. Repeat Table 3 above for fold-rise from baseline (of interest given the analysis cohort is

baseline seropositive).

The point and confidence interval estimates are computed using inverse probability sampling weights $\hat{w}_{subcohort.x}$ for 7+day Cases and for Non-cases, and using $\hat{w}_{start1.x}$ for 1-6+day Cases and 7+day Cases combined, as defined in Section 12.3.1.

### 12 Correlates of Risk Analysis Plan

This analysis plan for CoRs and CoPs focuses on the COVID primary endpoint, with its continuous failure times (failure time defined by the day of the event) and no competing risks.

#### 12.1 CoR Objectives

The following CoR objectives are assessed in baseline seronegative per-protocol vaccine recipients:

1. **Univariable CoR** To assess each individual Day 29 antibody marker as a CoR of outcome in vaccine recipients, adjusting for baseline factors
2. **Multivariable CoR** To build models predictive of outcome based on a set of Day 29 antibody marker readouts, adjusting for baseline factors

#### 12.2 Outline of the Set of CoR Analyses

The univariable CoR objective is addressed by Cox proportional hazards regression and nonparametric threshold regression. The multivariable CoR objective is addressed by superlearning. All of these analyses are implemented in automated and reproducible press-button fashion.

In addition, supportive exploratory analyses of the univariable CoR objective are conducted using flexible parametric regression modeling: generalized additive model regression.

#### 12.3 Day 29 Markers Assessed as CoRs and CoPs

The following Day 29 markers are assessed as CoRs and CoPs, usually as quantitative variables and in some analyses as ordered trinary variables or binary variables, all of which do not subtract Day 1 (baseline) values:

1. binding Ab to Spike (IgG BAU/ml)
2. binding Ab to RBD (IgG BAU/ml)
3. pseudovirus neutralization IU50/ml

For all univariable CoR analyses (first objective), the non-baseline subtracted versions of the Day 29 antibody markers are studied; the baseline-subtracted versions are not studied given that the analyses are done in the baseline seronegative cohort for which Day 1 readouts will generally be negative. The multivariable machine learning CoR analyses include synthesis markers that combine information across the individual markers listed above, as well as including 2FR and 4FR versions of variables.

##### 12.3.1 Inverse probability sampling weights used in CoR analyses

In section 9.1, estimated inverse probability sampling (IPS) weights $\hat{w}_{subcohort.x}$ were defined for per-protocol immunogenicity subcohort members, for the purpose of immunogenicity analyses. This section describes two IPS weights that are used for CoR and CoP analyses, the first for the main analyses that count cases starting 7 days post Day 29 ($\hat{w}_{start7.x}$) and the second for the sensitivity analyses that count cases starting 1 day post Day 29 ($\hat{w}_{start1.x}$).

Consider the correlates analyses that count cases starting 7 days post Day 29. For baseline sampling stratum $x$ [(vaccine, placebo) $\times$ (demographic strata)], the IPS weight $w_{start7.x}$ assigned to a non-case participant in stratum $x$ is defined by $\hat{w}_{start7.x} = 1/\hat{\pi}_{start7}(x) = N_x/n_x$, where $N_x$ is the number of stratum $x$ vaccine recipient non-cases in the Per-Protocol Baseline Seronegative (PPBN) cohort and $n_x$ is the number of these participants that also have Day 1 and Day 29 marker data available, where participants with any evidence of SARS-CoV-2 infection before 7 days post Day 29 visit are excluded from the counts $N_x$ and $n_x$. For non-case participant $i$ in the immunogenicity subcohort, $\hat{w}_{start7.i} = 1/\hat{\pi}_{start7}(X_i)$ denotes the weight $\hat{w}_{start7.x}$ for this individual's sampling stratum. All cases are assigned sampling weight $N_1/n_1$ where $N_1$ is the total number of vaccine recipient cases in the PPBN cohort restricting to cases with event time starting 7 days post Day 29, and $n_1$ is the number of these participants that also had the Day 1 and Day 29 markers measured, and again participants with any evidence of SARS-CoV-2 infection $<$ 7 days post Day 29 visit are excluded from the counts $N_x$ and $n_x$.

In terms of two-phase sampling data analysis nomenclature, "phase 1 ptids" are defined as the entire PPBN cohort except excluding participants with any evidence of SARS-CoV-2 infection $<$ 7 days post Day 29 visit. The "phase 2 ptids" are then the subset of these phase 1 ptids with Day 1 and Day 29 Ab marker data available. Thus the weight $\hat{w}_{start7.x}$ is the inverse sampling probability weight, calculated as the empirical fraction (No. phase 1 ptids / No. phase 2 ptids) within each of the baseline negative strata defining the immunogenicity subcohort sampling. For baseline seronegative individuals outside the phase 1 ptids, $\hat{w}_{start7.x}$ is assigned the missing value code NA. All other individuals have a positive value for $\hat{w}_{start7.x}$.

Next consider the correlates analyses of Day 29 markers that include cases starting 1 day post Day 29. For baseline sampling stratum $x$ [(vaccine, placebo) $\times$ (demographic strata)], the IPS weight $w_{start1.x}$ assigned to a non-case participant in stratum $x$ is defined by $\hat{w}_{start1.x} = 1/\hat{\pi}_{start1}(x) = N_x/n_x$, where $N_x$ is the number of stratum $x$ vaccine recipient non-cases in the PPBN cohort and $n_x$ is the number of these participants that also have Day 1 and Day 29 marker data available, where participants with any evidence of SARS-CoV-2 infection before 1 day post Day 29 visit are excluded from the counts $N_x$ and $n_x$. For non-case participant $i$ in the immunogenicity subcohort, $\hat{w}_{start1.i} = 1/\hat{\pi}_{start1}(X_i)$ denotes the weight $\hat{w}_{start1.x}$ for this individual's sampling stratum. All 1-6+day Cases and 7+day Cases are assigned sampling weight $N_1/n_1$ where $N_1$ is the total number of vaccine recipient cases in the PPBN cohort restricting to cases with event time starting 1 days post Day 29, and $n_1$ is the number of these participants that also had the Day 1 and Day 29 markers measured, and again participants with any evidence of SARS-CoV-2 infection < 1 days post Day 29 visit are excluded from the counts $N_x$ and $n_x$.

In terms of two-phase sampling data analysis nomenclature, for the Day 29 marker analyses "phase 1 ptids" are defined as the entire PPBN cohort except excluding participants with any evidence of SARS-CoV-2 infection < 1 days post Day 29 visit. The "phase 2 ptids" are then the subset of these phase 1 ptids with Day 1 and Day 29 Ab marker data available. Thus the weight $\hat{w}_{start1.x}$ is the inverse sampling probability weight, calculated as the empirical fraction (No. phase 1 ptids / No. phase 2 ptids) within each of the baseline negative strata used in the sampling design to define the immunogenicity subcohort. For baseline negative individuals outside the phase 1 ptids, $\hat{w}_{start1.x}$ is assigned the missing value code NA. All other individuals have a positive value for $\hat{w}_{start1.x}$. In sum, the weights $\hat{w}_{start1.x}$ are calculated in the same way as the weights $\hat{w}_{start7.x}$, except the relevant time window for evidence of infection or COVID is at least 1 days post Day 29 visit instead of at least 7 days post Day 29 visit.

##### 12.3.2 Choice of regression methods

Time-to-event methods use the Day 29 visit date as the time origin.

The IPW complete-case Cox regression model designed for case-cohort sampling designs will be used for estimation and inference on hazard ratios of outcomes by Day 29 marker levels, and for estimation and inference on marginalized marker-conditional cumulative incidence over time. The models will be fit using the *survey* R package available on CRAN, and will adjust for the baseline factors. We use a method from the survey package that assumes without replacement two-phase sampling and not Bernoulli sampling, which matches the sampling design and approach to weight estimation (**?**).

The final time point $t_F$ of follow-up for correlates analyses is taken to be 54 days after the Day 29 visit. Let $T$ be the failure time, $S$ a Day 29 marker of interest, and $X$ the vector of baseline factors that are adjusted for. With $S_1(t|s,x) = P(T > t|S = s, X = x, A = 1)$, the Cox model fit yields an estimate of $S_1(t|s, X_i)$ for each individual $i$ in the phase-two sample. The marginalized conditional risk $risk_1(t|s) = E_X[P(T \leq t|s, X, A = 1)]$ through time $t$ (for all times $t$ through $t_F$ simultaneously) is estimated based on the equation

$$risk_1(t|s) \quad = \quad \int (1 - S_1(t|s,x))dH(x) \tag{1}$$

where $H(\cdot)$ is the distribution of $X$ in $A = 1$ individuals.

Given the estimates $\hat{S}_1(t|s, X_i)$ for each of the $n_1$ participants in the PPBN cohort assigned to the vaccine arm, we then estimate $risk_1(t|s)$ by G-computation:

$$\widehat{risk}_1(t|s) = \sum_{i=1}^{n_1}(1 - \hat{S}_1(t|s, X_i)). \tag{2}$$

The bootstrap is used to obtain 95% pointwise confidence intervals for $risk_1(t_F|s)$.

The bootstrap process will be performed by resampling with replacement the subjects within the subcohort and the subjects outside the subcohort separately within each stratum and by resampling with replacement subjects with undetermined stratification variables. Across all bootstrap samples, the number of participants in each stratum in the immunogenicity subcohort remains fixed, but the number of cases does not stay the same.

The results of the above Cox modeling will be output in a variety of ways:

1. Plot $\widehat{risk}_1(t_F|s)$ vs. $s$ with 95% CIs for continuous $S = s$ varying over its whole range. Include on the plot the estimate of $\widehat{risk}_0(t_F)$ with a 95% CI for the placebo arm (horizontal bands), computed by a Cox model marginalizing over the same baseline factors as for the analysis of the vaccine arm.

2. Based on a fit of the Cox model to a nominal categorical antibody marker defined as the tertiles of $S$, plot $\widehat{risk}_1(t|s)$ for each category of $S$ values with 95% CIs, for all time points $t$ from Day 29 through $t_F$. If more than 20% of vaccine recipients have $S$ below the positive/negative cutoff of the assay, then the categories instead will be (1) values $\leq$ cutoff; (2) values below the median of values $>$ cutoff; (3) values above the median of values $>$ cutoff. Include on the plot the estimated curve $\widehat{risk}_0(t)$ with 95% CIs for the placebo arm, computed by a Cox model marginalizing over the same baseline factors as for the analysis of the vaccine arm.

3. Tabular reporting of the hazard ratio per 10-fold change in the quantitative Day 29 antibody marker with 95% confidence interval and 2-sided p-value

4. Tabular reporting of the hazard ratio for the Middle and Upper categories of the categorical Day 29 antibody marker vs. the Lower category, with 95% confidence interval and 2-sided p-value, as well as a global generalized Wald two-sided p-value for whether the hazard rate of the endpoint varies across the three categories. The table includes the attack rate (with no. of cases / no. at risk) through $t_F$ for each of the three vaccine marker subgroups and for the placebo arm.

5. Report point and 95% CI estimates for the hazard ratio per 10-fold change in the Day 29 antibody marker, for the entire per-protocol baseline seronegative vaccine cohort and for each of the baseline demographic strata subgroups defined in Table 7 (reported via forest plotting).

6. Westfall-Young (1997) q-values and FWER-adjusted p-values for the generalized Wald tests are included in the table.

The bootstrap is used to calculate 95% pointwise CIs for $risk_1(t_F|s)$ in $s$. The 2-sided Wald p-value for testing the regression coefficient of the marker in the Cox model provides a valid test of the null hypothesis $H_0 : risk_1(t_F|s) = risk_1(t_F)$ for all $s$, and is reported.

In addition, point estimates and 95% confidence intervals about $risk_1(t_F)$ will be computed by nonparametric monotone-dose response regression, as described in the beginning of Section 15.1.

Moreover, the same Cox model analysis described above will be used to estimate the alternative marginalized conditional risk parameter defined by $risk_1(t|S \geq s)$ where $risk_1(t|S \geq s) = E_X[P(T \leq t|S \geq s, X, A = 1)]$, which can be estimated by

$$\widehat{risk}_1(t|S \geq s) = \sum_{i=1}^{n_1}(1 - \hat{S}_1(t|S \geq s, X_i)).$$

This parameter is useful because typically subgroups of interest are defined by having marker response above a threshold. We will plot $\widehat{risk}_1(t_F|S \geq s)$ vs. $s$ with 95% CIs for continuous $S$ with $s$ varying over the range of $S$. This type of analysis is also included because it analyzes the same parameter as the nonparametric threshold estimation method described below, providing a way to address the threshold question both by Cox modeling and by nonparametric analysis.

If the outcome under study is subject to competing risks, then the Cox model is fit in the same way, except counting the competing risk as right-censoring. Now the parameter being estimated is the marginal conditional cumulative incidence function $risk_1(t, 1|s) = E_X[P(T \leq t, J = 1|s, X, A = 1)]$ where $J = 1$ is the outcome of interest.

The Fine-Gray proportional subdistribution hazards model is used to estimate $risk_1(t, 1|s, x)$ (implemented in the *cmprsk* R package available on CRAN). While we prefer the interpretation of cause-specific hazards (Prentice et al., 1978) to subdistribution hazards as in Fine and Gray (1999) in a rare event situation such as in the COVID-19 VE trials (vaccine arm), the Fine-Gray method is expected to give similar answers, except it is more easily implemented. Therefore, we estimate $risk_1(t, 1|s)$ by G-computation

$$\widehat{risk}_1(t, 1|s) = \sum_{i=1}^{n_1} \widehat{risk}_1(t, 1|s, X_i). \tag{3}$$

As for the analyses without competing risks, the bootstrap is used for calculating 95% confidence intervals and for testing $H_0 : risk_1(t_F, 1|s) = risk_1(t_F, 1)$ for all $s$.

###### 12.3.3 Univariate CoR: Nonparametric threshold regression modeling

The van der Laan et al. (2021) extension of the nonparametric CoR threshold estimation method of Donovan et al. (2019) is applied to each of the non-baseline subtracted Day 29 antibody markers, using the version accounting for right-censoring of some follow-up times, assessing failure through the fixed time point $t_F$. The analyses adjust for the same baseline factors $X$ as used in the Cox model CoR analyses.

The extension adjusts for baseline covariates by estimating the conditional mean function $E[I(T \leq t_F)|S \geq s, X, A = 1]$ using discrete-SuperLearner and then empirically averaging over the baseline covariates $X$ to estimate the marginal risk $risk_1(t_F|S \geq s) = E_X[I(T \leq t_F)|S \geq s, X, A = 1]$ for each threshold $s$ of the the antibody marker in a specified discrete set. We do not perform pooled regression across the thresholds $s$, which ensures we are totally nonparametric in estimating the threshold dependence of $E_X[I(T \leq t_F)|S \geq s, X, A = 1]$ on $s$. The SuperLearner library includes a range of increasingly flexible parametric learners including logistic regression (glm), bayesian logistic regression (bayesglm), and L1-penalized logistic regression (glmnet). (Two of each learner is included in the library, one with only main-term variables and another with main-term and interaction variables.)

An advantage of the nonparametric CoR threshold method compared to Cox modeling that specifies a log linear hazard ratio with the marker is that it can potentially detect a threshold of very low risk. The method is implemented with and without the monotonicity constraint that $risk_1(t_F|S \geq s)$ is monotone non-increasing in $s$, where the results assuming monotonicity are reported unless there is evidence for violation of this assumption.

The results are reported in the same way that Donovan et al. (2019) reports results in its Figure 2, where point estimates and simultaneous 95% confidence bands for $risk_1(t_F|S \geq s)$ are plotted for a range of threshold values (the simultaneous confidence bands cover the entire curve in $s$ with at least 95% probability). The method uses the same empirical two-phase sampling estimated weights (IPS weights) as used for the other univariable IPW complete-case CoR analyses. In addition, for each pre-specified risk threshold $c$ set to take values over a grid between 0 and the estimated outcome rate in placebo recipients, the method is applied to estimate the inverse function $s_c = inf\{s : E_X[I(T \leq t_F)|S \geq s, A = 1, X] \leq c\}$, where $s_c$ is estimated by substitution of the marginal risk function estimate. Note that the substitution estimator of $s_c$ requires that the marginal risk function is estimated for all thresholds, which is computationally infeasible. Instead, we estimate the marginal risk function on a sufficiently large discrete set and linearly interpolate to obtain marginal risk estimates for all thresholds outside the discrete set. In order for this estimand to be well defined, we operate (for this estimand only) under the assumption that $s \mapsto E_X[I(T \leq t_F)|S \geq s, A = 1, X]$ is monotone. For the substitution-based estimator of the inverse function $s_c$ to be well-defined, we require the estimate of $s \mapsto E_X[I(T \leq t_F)|S \geq s, A = 1, X]$ to be monotone as well. If there is evidence that the function estimate is not monotone then we replace the estimate with its monotone projection, which preserves its theoretical properties (**?**).

A plot of point and simultaneous 95% confidence interval estimates of $s_c$ (over the grid of $c$ values) is provided to help indicate marker thresholds defining subgroups with very low risk of outcome. The confidence interval estimates for $s_c$ are derived directly from the simultaneous 95% confidence band estimates for the marginal risk function $s \mapsto E_X[I(T \leq t_F)|S \geq s, A = 1, X]$, and therefore its estimates and inference are compatible with those of the marginal risk function. In particular, no multiple testing adjustments are needed.

The analysis is done using targeted maximum likelihood estimation (TMLE) as described in van der Laan, Zhang, and Gilbert (2021), and the simultaneous confidence bands are of the Wald-type, obtained from the asymptotic distribution of the TMLE. As for other correlates analyses, the baseline risk score and geographic region are adusted for.

##### 12.3.4 P-values and Multiple hypothesis testing adjustment for CoR analysis

In general, p-values are only reported from pre-specified and automated (press-button) analyses. For the CoR analyses, p-values are reported for the univariable Cox regression analyses of the specified Day 29 antibody marker variables. Two-sided p-values for hypothesis testing of a Day 29 marker CoR are calculated both for the Cox regression of quantitative markers (two-sided Wald tests), and for the Cox regression of markers binned into tertiles (two-sided Generalized Wald tests). Therefore a total of twelve 2-sided p-values for CoRs are calculated.

It is not completely clear whether to perform multiple hypothesis testing adjustment, given the expectation that the correlations among the markers are high, and possibly very high, meaning that multiplicity correction could incur a relatively high cost on the false negative error rate.

However, given that robust evidence supporting an antibody marker as a CoR will be required for qualifying a marker, we will conduct multiplicity adjustment for CoR analysis, as the ability to make an inference that a marker passed pre-specified multiplicity adjusted criteria should aid an overall evidence package for establishing a validated or non-validated surrogate endpoint. Therefore, multiplicity adjustment is performed across the twelve 2-sided p-values.

A permutation-based method (Westfall et al., 1993) will be used for both family-wise error rate (Holm-Bonferroni) and false-discovery rate (q-values; Benjamini-Hochberg) correction. $10^4$ replicates of the data under the null hypotheses will be created by randomly resampling the immunologic biomarkers with replacement. For each Cox regression CoR analysis the unadjusted p-value, the FWER-adjusted p-value, and the q-value is reported for whether there is a covariate-adjusted association, where all p-values and q-values are 2-sided. The FWER-adjusted p-values and q-values are computed pooling over both the quantitative marker and tertilized marker CoR analyses. As a guideline for interpreting CoR findings, markers with FWER-adjusted p-value $\leq 0.05$ are flagged as having statistical evidence for being a CoR. Additionally, markers with unadjusted p-value $\leq 0.05$ and q-value $\leq 0.10$ are flagged as having a hypothesis generated for being a CoR.

The FWER adjustment is done for all Day 29 markers among bAb Spike, bAb RBD, and PsV nAb ID50. It is required that antibody data are available for some of the neutralization markers before the multiplicity adjustment is performed. The first data analysis has available the bAb Spike, bAb RBD, and PsV nAb ID50 markers; therefore multiplicity adjustment is performed for this initial analysis.

##### 12.4 Multivariable CoR: Cox Proportional Hazards Models

Once the complete ADCP data are available, a multivariable Cox model is fit (using the same fitting approach as for individual markers) that includes the three Day 29 markers bAb RBD, pseudovirus nAb ID50, and ADCP, with the same baseline prognostic factors adjusted for in the univariable marker analyses also adjusted for in the multivariable marker analyses. Point estimates and 95% confidence intervals are reported for the 3 marker hazard ratio parameters. An unadjusted p-value from a generalized Wald test for whether the set of 3 markers has any correlation with HIV-1 acquisition is (rejecting the complete null hypothesis that the 3 hazard ratios are all unity) is reported. The p-values for the individual hazard ratio parameters are also reported but it is the generalized Wald test p-value that is the pre-specified test for whether the set of markers correlate with outcome.

In addition, the Cox models will be repeated in exploratory analyses with three separate Cox models fit for pairs of antibody markers: (1) D29 bAb RBD, D29 PsV nAb ID50; (2) D29 bAb

RBD, D29 ADCP; (3) D29 PsV nAb ID50, ADCP. Point estimates, 95% confidence intervals, and unadjusted p-values for the individual hazard ratios are reported for each hazard ratio parameter.

#### 12.5 Multivariable CoR: Superlearning of Optimal Risk Prediction Models

##### 12.5.1 Objectives

The multivariable CoR objective is addressed through two sub-objectives: first to build an 'estimated optimal surrogate' (Price et al., 2018), a model that best predicts the outcome from Day 29 antibody markers and baseline factors. The second sub-objective is estimation and inference on variable importance measures for each Day 29 antibody marker, for ranking of antibody markers by their importance/influence on predicting risk. The analysis plan is patterned off of the analysis of the HVTN 505 HIV-1 vaccine efficacy trial (Neidich et al., 2019). For these analyses both baseline-subtracted and non-baseline subtracted versions of the Day 29 markers are used, in a broader unbiased analysis to build models most predictive of outcome.

##### 12.5.2 Input variable sets

Day 29 antibody markers are classified into the following antibody marker variable sets, with individual variables listed within categories:

1. Binding antibody anti-Spike (S-bAb)
    a Day 29 anti-Spike IgG concentration
    b delta (Day 29 - Day 1) anti-Spike IgG concentration
    c indicator 2FR anti-Spike IgG concentration
    d indicator 4FR anti-Spike IgG concentration
2. Binding antibody anti-RBD (RBD-bAb)
    (a) Day 29 anti-RBD concentration
    (b) delta anti-RBD concentration
    (c) indicator 2FR anti-RBD concentration
    (d) indicator 4FR anti-RBD concentration
3. Pseudovirus neutralizing antibody anti-Spike (pseudovirus-nAb)
    a Day 29 anti-Spike IU50/ml
    b delta anti-Spike IU50/ml
    c indicator 2FR anti-Spike IU50/ml
    d indicator 4FR anti-Spike IU50/ml
4. ADCP score

a Day 29 ADCP score

b delta ADCP score

c indicator 2FR ADCP score

d indicator 4FR ADCP score

5. Functional markers subtracting out binding antibody

   a Day 29 log10 PsV nAb ID50 IU50/ml - Day 29 anti-RBD log10 BAU/ml

   b Day 29 log10 ADCP score - Day 29 anti-RBD log10 BAU/ml

For the primary analyses that only include PPBN vaccine recipients, the markers 1b, 2b, 3b, and 4b are excluded from the analysis, because for this cohort there is very little potential independent information in these markers compared to the Day 29 markers (that are not baseline subtracted).

The baseline factors without any marker data comprise a sixth set of variables to include in the superlearner modeling.

#### 12.6 Missing data

We expect a small amount of missing data from the three antibody markers (bAb Spike, RBD; pseudovirus-nAb IU50/ml), with possibly different participants missing data for different markers. We take the following approach to handle any missing data that occurs.

First, we define the two-phase sampling indicator $\epsilon$ as taking value of one if a participant has Day 1 and Day 29 bAb data for both Spike and RBD, where here we assume that the MSD platform is highly robust such that it will have nearly 100% complete data for sampled participants. Second, for the pseudovirus nAb ID50 marker, for participants with $\epsilon = 1$ but the Day 1 and/or Day 29 marker value is missing, we use single imputation to fill in any missing values, ignoring the uncertainty in the imputations in the analysis, because it should have negligible impact on results given the small amount of missing data. This process means that the two-phase data set has a simple 'all-or-nothing' missing data pattern where participants with $\epsilon = 1$ have all markers with Day 1 and Day 29 data, and are included in IPW complete-case analyses, and participants with $\epsilon = 0$ have some or all markers missing and are excluded from IPW complete-case analyses. This means that all IPW complete-case data analyses can use the same empirical frequency (IPS) sampling weights.

##### 12.6.1 Implementation of superlearner

For baseline risk score development, Superlearner is applied to the placebo arm only as mentioned in Section 10. For multivariable immune correlates of risk/estimated optimal surrogate development, Superlearner is applied to the vaccine arm only. The following details are used in the implementation of superlearner of the vaccine arm only:

- Pre-scale each quantitative and ordinal variable to have empirical mean 0 and standard deviation 1.

- For the immune correlates analysis, the final library of learners is selected accounting for the number of phase-two endpoint cases in the vaccine arm. Since for ENSEMBLE, there are large numbers of endpoint cases in the vaccine arm, 5-fold cross-validation will be used, and no more than floor($n_v/6$) input variables will be used in the model where $n_v$ is the number of evaluable vaccine endpoint cases.
- Include learning algorithms with and without screening of variables. Screens used will be: 1) glmnet (lasso) pre-screening (with default tuning parameter selection), 2) logistic regression univariate 2-sided p-value screening (at level $p < 0.10$), and 3) high-correlation variable screening (described below). For ENSEMBLE there are enough vaccine breakthrough cases to include the adaptive learners for the overall report but not for the region-specific reports. All of the selected learners are coded into the SuperLearner R package available on CRAN.
- Include high-correlation variable screening, not allowing any pair of input variables to have Spearman rank correlation $r > 0.9$. When a pair of variables has $r > 0.9$, the variable with the highest ranked signal-to-noise ratio (i.e., biological dynamic range) is selected; if these data are not available (they are not for ENSEMBLE) or there is a tie then variables are selected in the following order of priority: first pseudovirus nAb ID50, second IgG RBD, third IgG Spike. Given than the Spike and RBD variables are expected to be very highly correlated at Day 29, any model that would consider both Spike and RBD includes only RBD.
- The superlearner is conducted averaging over 10 random seeds, to make results less dependent on random number generator seed.
- All of the learners are implemented with IPS weighting, using the IPS weights $w_{start7.i} = 1/\hat{\pi}_{start7}(X_i)$ described in Section 12.3.1) to account for the two-phase sampling design. Correspondingly, all endpoints starting 7 days post D29 are included.
- Discrete-SL estimated models, derived using the learning algorithms specified in Table 9, will be used to compare the relative performance for each of the variable sets based off the estimated CV-AUC with a 95% confidence interval.
- Two levels of cross-validation are used:
  * Outer level: CV-AUC computed over 5-fold cross-validation repeated 10 times to improve stability
  * Inner level: 5-fold CV is used.
- Results for comparing classification accuracy of different models are based on point and 95% confidence interval estimates of cross-validated area under the ROC curve (CV-AUC) and difference in CV-AUC as a predictiveness metric (Hubbard et al., 2016; Williamson et al., 2022). Results are presented as forest plots of point and 95% confidence interval estimates similar to those used in Neidich et al. (2019) (Figure 3) and Magaret et al. (2019). CV-AUC is estimated using the R package *vimp* available on CRAN, including the IPS weights that are used for other data analyses.

- Multivariable CoR analysis using the Superlearner will be run only upon availability of fuller ADCP marker data.

Table 9 lists the learning algorithms that are applied to estimate the conditional probability of the outcome based on the input variable sets considered above. Most of the algorithms are non-data-adaptive type learning algorithms, such as parametric regression models (e.g., glms), which are simple, stable and advantageous for an application with limited number of endpoint events. Data-adaptive type algorithms are also included, for increasing flexibility of modeling: SL.randomForest, SL.gam, SL.polymars, and SL.xgboost. All of the selected learners are coded into the SuperLearner R package.

Before fitting the superlearner models to the vaccine arm data, a decision will be made on how to define the "baseline risk factors" input variable set, based on prediction-accuracy results of the Superlearner analysis that built the baseline behavioral risk score based on the placebo arm, as well as on external knowledge of important individual risk factors.

For ENSEMBLE the baseline factors are defined as the baseline risk score, indicator of being at heightened risk for COVID (a randomization factor) and the two indicators coding for the three geographic regions.

For the immune correlates objective the superlearner model is fit to each of the following variable sets, with immunological variables listed in Section 12.5.2:

1. Baseline risk factors
2. Baseline risk factors and the bAb anti-Spike markers
3. Baseline risk factors and the bAb anti-RBD markers
4. Baseline risk factors and the pseudovirus-nAb IU50/ml markers
5. Baseline risk factors and the ADCP score markers
6. Baseline risk factors and the functional markers subtracting out binding antibody
7. Baseline risk factors and the bAb markers and the pseudovirus-nAb IU50/ml markers
8. Baseline risk factors and the bAb markers and the ADCP score markers
9. Baseline risk factors and the bAb markers and the pseudovirus-nAb IU50/ml markers and the difference in these two markers
10. Baseline risk factors and the bAb markers and the ADCP score markers and the difference in these two markers
11. Baseline risk factors and the pseudovirus-nAb IU50/ml markers and the ADCP markers
12. Baseline risk factors and all individual marker variables
13. Baseline risk factors and the bAb markers and the combination scores across the four markers [PCA1, PCA2, FSDAM1/FSDAM2 (the first two components of nonlinear PCA), and the maximum signal diversity score He and Fong (2019)].

14. Baseline risk factors and the pseudovirus-nAb IU50/ml markers and the combination scores across the four markers
15. Baseline risk factors and the ADCP score markers and the combination scores across the four markers
16. Baseline risk factors and all individual marker variables and all combination scores (full model)

Therefore in total, 16 variable sets are studied. The reason to include the baseline risk score factors only variable set is to investigate how much incremental improvement in predicting outcome is obtained by adding antibody marker variables on top of baseline demographic/exposure factors. The other variable sets are designed to compare the four immunoassay types by their predictiveness and to investigate incremental predictive value in using multiple immunoassays. The final variable set is included as the full model that considers all variables together, which serves as another reference model.

Table 9: Learning Algorithms in the Superlearner Library of Estimators of the Conditional Probability of Outcome, for Building the Baseline Risk Score Based on the Placebo Arm[1].

| Algorithms | Screens/ Tuning Parameters |
|---|---|
| SL.mean | None |
| SL.glm | Low-collinearity and (All, Lasso, LR) |
| SL.bayesglm | Low-collinearity and (All, Lasso, LR) |
| SL.glm.interaction | Low-collinearity and (All, Lasso, LR) |
| SL.glmnet | (alpha=1; All) |
| SL.gam | Low-collinearity and (Lasso, LR) |
| SL.ksvm | Low-collinearity and (kernel="rbfdot","polydot") and (Lasso, LR) |
| SL.polymars | Low-collinearity and (Lasso, LR) |
| SL.xgboost[3] | All and (maxdepth,shrinkage,balance)= (4, 0.1, no) |
| SL.ranger[3] | All and balance = no |

[1] All continuous and ordinal covariates are pre-standardized to have empirical mean 0 and standard deviation 1.

[2] **All** = include all variables; **Lasso** = include variables with non-zero coefficients in the standard implementation of SL.glmnet that optimizes the lasso tuning parameter via cross-validation; **Low-collinearity** = do not allow any pairs of quantitative variables with Spearman rank correlation $> 0.90$; **LR** = Univariate logistic regression Wald test 2-sided p-value $< 0.10$.

In order to evaluate the relative performance of the Discrete-Superlearner estimated models for each of the variable sets, derived using the learning algorithms specified in Table 9, the CV-AUC is estimated with a 95% confidence interval (Hubbard et al., 2016; Williamson et al., 2022). The point and 95% confidence interval estimates of CV-AUC are reported in a forest plot, which provide a way to discern which Day 29 antibody assays and readouts/markers provide the most information in predicting COVID or other outcomes. The specified library of learners may be modified prior to SAP finalization (before breaking the blind of case/non-case status). As noted above CV-AUC is estimated using the R package *vimp* available on CRAN, which uses augmented inverse probability weighting to properly estimate CV-AUC accounting for the two-phase sampling design.

In addition, for selected variable sets, similar forest plots will be made comparing performance of the various estimated models (e.g., by individual learning algorithm types such as lasso), including discrete superlearner and superlearner models. The plot will be examined to determine which individual learning algorithm types are performing the best. If there is an interpretable algorithm that has performance close to the best-performing algorithm (which is most likely to be the superlearner), then it will be fit on the entire data set of vaccine recipients and the estimated model presented in a table.

Cross-validated ROC curves are plotted for the superlearner estimated models for each of the input variable sets. In addition, boxplots of cross-validated estimated probabilities of outcome by case-control status (as estimated from the superlearner models) are plotted.

###### 12.6.2 Variable set and individual variable importance

The importance of variable sets (and individual variables) will be summarized by the estimated gain in population prediction potential (also referred to as the intrinsic importance) when comparing each variable set plus baseline risk factors to baseline risk factors alone. Prediction potential (predictiveness) will be measured using CV-AUC. Inference on the intrinsic importance will be based off sample splitting; thus, both the estimated variable importance and the estimated CV-AUC of each variable set when evaluated on independent data from the data used to evaluate the CV-AUC of the baseline risk factors will be reported. The class-balancing versions of SL.xgboost will be dropped from the Super Learner library in the variable importance computation as the regression carried out to account for the two-phase sampling will be based on a continuous outcome (so there won't be any imbalance).

#### 13 Correlates of Protection: Generalities

In general, for all of the correlate of protection analyses, the same antibody markers are assessed that were analysed as correlates of risk: the Day 29 antibody markers not subtracting for the Day 1 baseline readout are used. Each of the Day 29 antibody biomarkers is separately studied as CoPs by the different analysis approaches summarized below.

#### 14 Correlates of Protection: Correlates of Vaccine Efficacy Analysis Plan

For each of the Day 29 antibody biomarkers, the method of Gilbert et al. (2020b) will be used to estimate $VE(1)$, $VE(0)$, and $(1 - VE(1))/(1 - VE(0))$, each with a 95% confidence interval and a 95% estimated uncertainty interval (EUI), where $VE(1)$ is vaccine efficacy for the subgroup of vaccine recipients with Day 29 marker if assigned vaccine above a specified cut-point value $s_{cut}$, and $VE(0)$ is vaccine efficacy for the subgroup of vaccine recipients with Day 29 marker if assigned vaccine not greater than $s_{cut}$. The analysis will be done under the **NEH** assumption ("no early harm") of Gilbert et al. (2020). The analysis is done for the cut-point defined by negative vs. positive for each marker, in particular defined by the positivity cut-off for bAb Spike and bAb RBD, and defined by the LOD for PsV nAb ID50. These analyses include p-values testing whether vaccine efficacy is different for vaccine recipients with negative vs. positive response. In addition, an exploratory analysis without p-values is done with cut-point defined by each 10th percentile above the positive/negative dividing line (starting at the 20th percentile at the lowest), up to the 80th percentile. This analysis method does not require closeout placebo vaccination (CPV) (Follmann, 2006) or a good baseline immunogenicity predictor of the Day 29 antibody marker. The method is implemented using code on the correlates_reporting2 Github repository, with applies Bryan Blette's R package "psbinary" posted at his Github repository.

A limitation of the Gilbert et al. method is that it only assesses a binary biomarker. Other analyses will be considered to estimate $VE(s)$ over biomarker values $s$ over the entire range, treating $S$ as a quantitative or categorical variable, and gaining efficiency by incorporating CPV and/or putative baseline immunogenicity predictors (BIPs). Based on earlier simulation studies (Follmann, 2006; Huang et al., 2013, e.g.,), methods that only leverage CPV data tend to have low power relative to methods that leverage BIP data alone (BIP-only methods) or both BIP and CPV data (BIP+CPV methods). Therefore, the key for improving efficiency will be the availability of a BIP. VE curve analysis for continuous $S$ will thus be conducted contingent on the availability of a BIP that satisfies the $R^2$ criterion outlined in Table 11. It is anticipated that post-crossover immune response marker data will not be available in early correlates analyses, and so BIP-only methods will be used in these initial analyses. When CPV data becomes available, new BIP+CPV analyses will be conducted that incorporate this new information. Details of the BIPs used can be found at the end of this section.

Let $Y(a)$ denote the potential binary outcome of interest if receiving intervention $a$, with $a = 1, 0$ standing for assignment to vaccine and placebo, respectively. Let $S(a)$ denote the potential biomarker value if receiving intervention $a$. The vaccine efficacy curve (Follmann, 2006; Gilbert and Hudgens, 2008) is defined as the curve of vaccine efficacy as a function of the immune response biomarker if assigned vaccination (i.e., $S(1)$): $VE(s) = 1 - P(Y(1) = 1|S(1) = s)/P(Y(0) = 1|S(1) = s)$. It characterizes the percentage reduction in clinical risk under vaccine assignment compared to under placebo assignment conditional on $S(1)$ and informs about the magnitude of potential immune response associated with certain levels of VE. Consider the existence of BIPs $X$ correlated with $S(1)$ and/or a CPV component in the trial where a subset of placebo recipients free of the outcome are vaccinated and have their immune response biomarkers measured as substitutes for $S(1)$. Under the NEE assumption and assuming the set of participants with $S(1)$ available is nested within the set of participants with BIP measures, the pseudo-score estimation method (Huang et al., 2013; Zhuang et al., 2019) based on discrete BIP measures allowing for adjustment of $X$ will be adopted for estimating the risk model $P(Y(z) = 1|S(1), X)$ and subsequently $VE(s) = 1 - \int P(Y(1) = 1|S(1), x)dF_X(x|S(1))/\int P(Y(0) = 1|S(1), x)dF_X(x|S(1))$. Hypothesis testing will be conducted for testing the null hypothesis that the VE curve is constant (Zhuang et al., 2019). Estimated parametric (Gilbert and Hudgens, 2008), semiparametric (Huang and Gilbert, 2011), or nonparametric (Li and Luedtke, 2020) likelihood estimators of VE curves will be applied to continuous BIPs. In scenarios where some BIPs are not measured from all trial participants, VE curve estimators accounting for this monotone missingness in $X$ and $S(1)$ will be adopted (Huang, 2018). If the data support positive vaccine efficacy before Day 29, sensitivity analysis approaches will be conducted for VE curve estimation under the NEH assumption. In the presence of multiple candidate biomarkers and when a CPV component is present, a multiple imputation approach as proposed in Dasgupta and Huang (2019) will be utilized to impute missing $S(1)$ data for selecting markers from multiple candidates and deriving a univariate marker score for VE curve estimation.

Finally, for scenarios with very rare events such that methods described above lack precision even with a CPV component but where the available BIP still satisfies the $R^2$ criterion outlined in Table 11, we will adopt sensitivity analysis methods that model the placebo risk conditional on the counterfactual $S(1)$ based on a sensitivity parameter that varies over some pre-specified range.

Among different strategies to identify BIPs, the following will be tried. First, we will study Day 1 bAb or nAb response to Ad26 as a BIP for the Day 29 markers of interest. Second, we will check whether Day 1 bAb or nAb to Nucleocapsid protein is a BIP for the anti-Spike/anti-RBD Day 29 markers of interest. The rationale for this latter analysis is that some studies have shown cross-reactive responses to Nucleocapsid protein and to common circulating human coronaviruses.

We will also evaluate using a multivariate BIP that corresponds to all of these aforementioned candidate univariate BIPs, which may help to achieve the target $R^2$ (see Table 11). When doing this, a separate BIP $W$ will be used for each vaccine-induced immune response marker $S(1)$. Let $Y(a)$ be the counterfactual outcome of interest — e.g., a COVID disease endpoint by a prespecified time — if randomization assignment had been set to $A = a$. The analyses conducted will provide unbiased estimates of the estimands of interest when $Y(a) \perp W|S(1)$ for $a \in \{0, 1\}$. The BIP $W$ will be a learned function of baseline covariates $L$ — that is, $W = f(L)$ for a function $f$ that will be learned based on the available data. All available baseline covariates will be considered for inclusion in $L$, including age, BMI, Day 1 bAb or nAb response to Ad26, and Day 1 bAb or nAb to Nucleocapsid protein. If the trial has more than 100 events on the vaccine arm in the subgroup of interest, then $f$ will be chosen to be an estimate of the following population-level optimization problem:

$$\begin{aligned} &\text{minimize } E[\{S - f(L)\}^2|A = 1] \\ &\text{subject to } f(L) \perp Y|A = 1, S. \end{aligned}$$

The rationale for choosing $f$ to (approximately) solve this optimization problem is that the BIP should be maximally predictive of $S$, while also satisfying the needed conditional independence assumption $Y(a) \perp W|S(1)$ when $a = 1$. Moreover, the needed conditional independence assumption $Y(a) \perp W|S(1)$ for the case that $a = 0$ is most plausible when this assumption is also satisfied for the case that $a = 1$. Also, because $W = f(L)$ for some function $f$, $Y(0) \perp W|S(1)$ is always more plausible than $Y(a) \perp L|S(1)$.

The solution to the above optimization problem is given by:

$$f(\ell) := \theta(\ell) - \frac{E[\theta(L)r(L)]}{E[r(L)^2]} r(\ell)$$

where $\theta(\ell) := E\{S|A = 1, L = \ell\}$, $r(\ell) := \frac{m(\ell)}{E[m(L)]} - \frac{1-m(\ell)}{1-E[m(L)]}$ and $m(\ell) := E[Y|A = 1, L = \ell]$. The following strategy is used to estimate this solution:

1. Obtain an estimate $\hat{\theta}$ of the function $\theta$ by running a Superlearner of $S$ against $L$ in the vaccine arm, where inverse probability of sampling weights are used to account for two-phase sampling of the marker.

2. Obtain an estimate $\hat{m}$ of $m$ by using Superlearner to regress $Y$ against $L$ in the vaccine arm.

3. Obtain an estimate $\hat{r}$ via a plug-in estimator, where $E[m(L)]$ is estimated by taking the empirical mean of $\hat{m}(L)$.

4. The final estimate $\hat{f}$ of $f$ is given by

$$\hat{f}(\ell) := \hat{\theta}(\ell) - \frac{\hat{E}[\hat{\theta}(L)\hat{r}(L)]}{\hat{E}[\hat{r}(L)^2]} \hat{r}(\ell),$$

where $\hat{E}$ denotes an empirical expectation.

Each Superlearner will be run using the same library and settings described in Table 9. If the trial has fewer than 100 events on the vaccine arm, then the function $f$ will be learned via Step 1 above only — that is, we will take $\hat{f} = \hat{\theta}$. All standard errors will be obtained via the bootstrap, with the above fitting of $\hat{f}$ redone within each bootstrap sample.

### 15 Correlates of Protection: Interventional Effects

In these analyses, we seek to understand whether, how, and to what extent Day 29 antibody markers impact vaccine efficacy in causal ways. We describe three approaches to this problem. Each involves consideration of a binary counterfactual outcome $Y(a, s)$ (e.g., indicator of the COVID disease endpoint by a pre-specified time) under a hypothetical intervention that both sets randomization assignment $A = a$ and sets the Day 29 immunologic marker $S$ to a fixed value or based upon a random draw from a analyst-specified distribution. Below, we assume that $S$ is scalar-valued, but some of the approaches below naturally extend to the case where a vector of immunologic markers are considered (currently such analyses are not planned). Given the central goal to develop a parsimonious surrogate endpoint based on a single immunoassay, the main analysis will use each of the methods to assess each of the quantitative readouts (not baseline-subtracted) separately as CoPs, adjusting for the same set of baseline covariates as used in the CoR analyses previously described in Section 12.

#### 15.1 CoP: Controlled Vaccine Efficacy

We first describe the controlled vaccine efficacy curve defined as

$$\text{CVE}(s) = 1 - \frac{P(Y(1, s) = 1)}{P(Y(0) = 1)} \; .$$

The value CVE($s$) takes represents the relative decrease in endpoint frequency achieved by administering vaccine and setting Day 29 immunologic marker level to $s$ compared to the placebo control intervention. Under our approach, the value of CVE($s$) is assumed to be monotone non-decreasing in $s$; in other words, vaccine efficacy can only potentially be improved by setting greater marker levels. The extent to which the marker plays a role in determining vaccine efficacy can be determined by the degree of flatness of the graph of CVE($s$) versus $s$.

In addition, because the primary study cohort for correlates analysis is naive to SARS-CoV-2, each of the Day 29 markers $S$ has no variability in the placebo arm [all values are 'negative,' below the assay cutoff for determining a negative or positive response]. Therefore, advantageously in this setting $CVE(s)$ has a special connection to the mediation literature (Cowling et al., 2019) where $CVE(s < cutoff)$ is the natural direct effect, and vaccine efficacy is 100% mediated through $S$ if and only if $CVE(s < cutoff) = 0$. Thus inference on $CVE(s < cutoff)$ evaluates full mediation.

Since $P(Y(0) = 1) = P(Y = 1 \mid A = 0)$ in view of vaccine versus placebo randomization, the controlled vaccine efficacy CVE($s$) at level $s$ can be identified using the fact that

$$P(Y(1, s) = 1) = E\left[P(Y = 1 \mid S = s, A = 1, X)\right]$$

whenever $Y(1, s)$ and $S$ are independent given $A = 1$ and a vector $X$ of covariates, and $P(S = s \mid A = 1, X) > 0$ almost surely. In other words, identification of the controlled vaccine efficacy requires that a rich enough set of covariates be available so that deconfounding of the relationship between endpoint $Y$ and marker $S$ is possible in the subpopulation of PPBN vaccine recipients, and that marker level $S = s$ may occur within each subpopulation defined by values of the covariates $X$ (positivity).

For each $s$, the identified parameter corresponding to CVE($s$) is an irregular parameter within nonparametric models, making its estimation at root-$n$ rate impossible; this significantly complicates estimation and inference on CVE($s$). Fortunately, the monotonicity of $s \mapsto$ CVE($s$) provides an opportunity to circumvent these difficulties. Similarly to Westling et al., 2020's approach for the causal dose-response function, we will use the general methodological template proposed in Westling and Carone, 2020 to derive (i) a nonparametric Grenander-type estimator of CVE($s$) and (ii) a plug-in confidence interval for CVE($s$) based on an asymptotic Chernoff limit. This estimator will require, as an intermediate step, estimation of several nuisance functions, including the outcome regression $P(Y = 1 \mid S = s, A = 1, X = x)$ and the propensity score $P(S = s \mid X = x, A = 1)$. These nuisance functions will be estimated using the Superlearner ensembling algorithm with a rich library including both parametric regression methods as well as flexible machine learning tools. Additionally, this method will be adapted to cases in which the marker has a mixed distribution involving a point mass at the LLOQ/2 value by constructing a nonparametric estimator of CVE(LLOQ/2) using the method of **?** and combining this with the main nonparametric estimator.

The monotonicity-based procedure we apply facilitates statistical inference for CVE($s$) for each $s$ separately, where point estimates and 95% confidence intervals for CVE($s$) will be presented.

We apply the same Cox modeling approach described in Section 12.3.2 to estimate

$$P(Y(1,s)=1) = E\left[P(Y=1 \mid S=s, A=1, X)\right],$$

with 95% confidence intervals, augmented with a sensitivity analysis, with advantages of harmonization with the CoR analysis, sensitivity analysis that is generally warranted when a no unmeasured confounders assumption is made, and efficiency gain achieved via the added modeling assumptions. The sensitivity analysis quantifies the rigor of evidence for a controlled VE CoP after accounting for potential bias from unmeasured confounding.

Gilbert et al. (2020a) details the sensitivity analysis approach, which was applied to the CYD14 and CYD15 dengue phase 3 data sets (Moodie et al., 2018); we plan to apply it in the same way to the COVID-19 data sets (as the structure of the problem is the same). We summarize here the essential details needed for application to the COVID-19 data sets.

We define $S$ to be a controlled risk CoP if $P(Y(1,s)=1)$ is monotone non-increasing in $s$ with $P(Y(1,s)=1) > P(Y(1,s')=1)$ for at least some $s < s'$, where point and 95% confidence interval estimates of $P(Y(1,s)=1)$ versus $s$, with built in robustness to unmeasured confounding, describe the strength of the CoP in terms of the amount and nature of decrease. Suppose the CoR analysis based on the Cox model is conducted as described in Section 12.3.2.

Let marginalized conditional risk

$$r_M(s) = risk_1(t_F|s)$$

and controlled risk

$$r_C(s) = P(Y(1,s)=1).$$

Given that CoR analysis is based on observational data — the biomarker value is not randomly assigned — a central concern is that unmeasured or uncontrolled confounding of the association between $S$ and $Y$ could render $r_M(s) \neq r_C(s)$, biasing estimates of the controlled risk curve $r_C(s)$ and of controlled risk ratios of interest

$$RR_C(s_1,s_2) = r_C(s_2)/r_C(s_1)\ .$$

Because we can never be certain that confounding is adequately adjusted for, sensitivity analysis is warranted, as considered in extensive literature — see, e.g., VanderWeele and Ding (2017) and references therein. Sensitivity analysis is useful to evaluate how strong unmeasured confounding would have to be to explain away an observed causal association, that is, to determine the strength of association of an unmeasured confounder between $S$ and

$Y$ needed for the observed exposure-outcome association to not be causal, $r_M(s) \neq r_C(s)$. We follow the recommendation of VanderWeele and Ding (2017) to report the E-value as a summary measure of the evidence of causality, or, in our application, evidence of whether $S$ is a controlled risk CoP based on variation in the controlled risk curve. We also include other closely related measures of sensitivity.

The E-value is the minimum strength of association, on the risk ratio scale, that an unmeasured confounder would need to have with both the exposure ($S$) and the outcome ($Y$) in order to fully explain away a specific observed exposure–outcome association, conditional on the measured covariates [VanderWeele and Ding (2017); VanderWeele and Mathur (2020)]. If, as in CoP analyses, the estimated marginalized risk ratio $\widehat{RR}_M(s_1, s_2) = \widehat{r}_M(s_2)/\widehat{r}_M(s_1)$ for $s_1 < s_2$ is less than one, then the E-value for $\widehat{RR}_M(s_1, s_2)$ is calculated as

$$e_{RR}(s_1, s_2) = \frac{1 + \sqrt{1 - \widehat{RR}_M(s_1, s_2)}}{\widehat{RR}_M(s_1, s_2)} \,. \tag{4}$$

We include the argument $(s_1, s_2)$ in the notation, with $s_1 < s_2$ by convention, to be clear that the E-value depends on specification of two specific marker-level subgroups.

To illustrate the interpretation of an E-value, suppose $S$ is binary and regression analysis yields an estimate $\widehat{RR}_M(0, 1) = \widehat{r}_M(1)/\widehat{r}_M(0) = 0.40$ with 95% confidence interval (CI) $(0.14, 0.78)$. An E-value $e(0, 1)$ of 4.4 means that a marginalized risk ratio $RR_M(0, 1)$ at the observed value 0.40 could be explained away (i.e., $RR_C(0, 1) = 1.0$) by an unmeasured confounder associated with both the exposure and the outcome by a marginalized risk ratio of 4.4-fold each, after accounting for the vector $X$ of measured confounders, but that weaker confounding could not do so.

In addition, we follow the recommendation of VanderWeele and Ding (2017) to also report the E-value $e_{UL}(s_1, s_2)$ for the upper limit $\widehat{UL}(s_1, s_2)$ of the 95% CI for the observed marginalized risk ratio $\widehat{RR}_M(s_1, s_2)$, computed as 1 if $\widehat{UL}(s_1, s_2) \geq 1$ and, otherwise, as

$$\frac{1 + \sqrt{1 - \widehat{UL}(s_1, s_2)}}{\widehat{UL}(s_1, s_2)} \,,$$

which in the example equals $e_{UL}(0, 1) = 1.88$. This E-value for the upper limit indicates, for given $s_1 < s_2$, the strength of unmeasured confounding at which statistical significance of the inference that $RR_C(s_1, s_2) < 1$ would be lost. The two E-values above are useful for judging how confident we can be that an immunologic biomarker is a controlled risk CoP, with E-values near one suggesting weak support and evidence increasing with greater E-values.

$RR_C(s_1, s_2) = (1 - CVE(s_2))/(1 - CVE(s_1))$, evidence for $RR_C(s_1, s_2) < 1$ is equivalently evidence for $CVE(s_1) < CVE(s_2)$. Thus in a placebo-controlled trial $RR_C(s_1, s_2)$ can be interpreted as the multiplicative degree of superior vaccine efficacy caused by marker level $s_2$ vs. marker level $s_1$, and E-values equivalently quantify evidence for whether $CVE(s_1)$ differs from $CVE(s_2)$.

It is also useful to provide conservative estimates of controlled risk ratios and of the controlled risk curve, accounting for unmeasured confounding. We approach these tasks based on the sensitivity analysis, or bias analysis, approach of Ding and VanderWeele (2016). We give their main result and refer readers to the paper for details. We begin by defining two (possibly context-specific) fixed sensitivity parameters. First, we set $RR_{UD}(s_1, s_2)$ to be the maximum risk ratio for the outcome $Y$ comparing any two categories of the unmeasured confounders $U$, within either exposure group $S = s_1$ or $S = s_2$, conditional on the vector $X$ of observed covariates. Second, we set $RR_{EU}(s_1, s_2)$ to be the maximum risk ratio for any specific level of the unmeasured confounder $U$ comparing individuals with $S = s_1$ to those with $S = s_2$, with adjustment already made for the measured covariate vector $X$. Thus, $RR_{UD}(s_1, s_2)$ quantifies the importance of the unmeasured confounder $U$ for the outcome, and $RR_{EU}(s_1, s_2)$ quantifies how imbalanced the exposure/marker subgroups $S = s_1$ and $S = s_2$ are in the unmeasured confounder $U$. The values $RR_{UD}(s_1, s_2)$ and $RR_{EU}(s_1, s_2)$ are always specified as greater than or equal to one. We suppose that $RR_M(s_1, s_2) < 1$ for the fixed values $s_1 < s_2$ — this is the case of interest for immune correlates.

Define the bias factor

$$B(s_1, s_2) = \frac{RR_{UD}(s_1, s_2) RR_{EU}(s_1, s_2)}{RR_{UD}(s_1, s_2) + RR_{EU}(s_1, s_2) - 1}$$

for $s_1 \leq s_2$, and define $RR_M^U(s_1, s_2)$ the same way as $RR_M(s_1, s_2)$, except marginalizing over the joint distribution of $X$ and $U$. Then, $RR_M^U(s_1, s_2) \leq RR_M(s_1, s_2) \times B(s_1, s_2)$, where $RR_M^U(s_1, s_2) = E\{r(s_2, X^*)\}/E\{r(s_1, X^*)\}$ with $X^* = (X, U)$ and $r$ conditional risk.Ding and VanderWeele (2016)

Translating this result to our problem context, under the positivity assumption, we have that $RR_M^U(s_1, s_2) = RR_C(s_1, s_2)$ and so, it follows that

$$RR_C(s_1, s_2) \leq RR_M(s_1, s_2) \times B(s_1, s_2) \ . \tag{5}$$

This inequality states that the causal risk ratio is bounded above by the marginalized risk ratio multiplied by the bias factor. It follows that a conservative (upper bound) estimate of $RR_C(s_1, s_2)$ is obtained as $\widehat{RR}_M(s_1, s_2) \times B(s_1, s_2)$, and a conservative 95% CI is obtained by multiplying each confidence limit for $RR_M(s_1, s_2)$ by $B(s_1, s_2)$. These estimates for $RR_C(s_1, s_2)$ account for the presumed-maximum plausible amount of deviation from the no unmeasured confounders assumption specified by $RR_{UD}(s_1, s_2)$ and $RR_{EU}(s_1, s_2)$. An appealing feature of this approach is that the bound (5) holds without making any assumption about the confounder vector $X$ or the unmeasured confounder $U$.

The above approach does not directly provide a conservative estimate of the controlled risk curve $r_C(s)$, because additional information is needed for absolute versus relative risk estimation. To provide conservative inference for $r_C(s)$, we next select a central value $s^{cent}$ of $S$ such that $\widehat{r}_M(s^{cent})$ matches the observed overall risk, $\widehat{P}(Y = 1|A = 1)$. This value is a 'central' marker value at which the observed marginalized risk equals the observed overall risk. Next, we 'anchor' the analysis by assuming $r_C(s^{cent}) = r_M(s^{cent})$, where picking the central value $s^{cent}$ makes this plausible to be at least approximately true. Under this assumption, the bound (5) implies the bounds

$$r_C(s) \leq r_M(s)B(s^{cent}, s) \quad \text{if} \quad s \geq s^{cent} \tag{6}$$

$$r_C(s) \geq r_M(s)\frac{1}{B(s, s^{cent})} \quad \text{if} \quad s < s^{cent}. \tag{7}$$

Therefore, after specifying $B(s^{cent}, s)$ and $B(s^{cent}, s)$ for all $s$, we conservatively estimate $r_c(s)$ by plugging $\widehat{r}_M(s)$ into the formulas (6) and (7). Because $B(s_1, s_2)$ is always greater than one for $s_1 < s_2$, formula (6) pulls the observed risk $\widehat{r}_M(s)$ upwards for subgroups with high biomarker values, and formula (7) pulls the observed risk $\widehat{r}_M(s)$ downwards for subgroups with low biomarker values. This makes the estimate of the controlled risk curve flatter, closer to the null curve, as desired for a sensitivity/robustness analysis.

To specify $B(s_1, s_2)$, we note that it should have greater magnitude for a greater distance of $s_1$ from $s_2$, as determined by specifying $RR_{UD}(s_1, s_2)$ and $RR_{EU}(s_1, s_2)$ increasing with $s_2 - s_1$ (for $s_1 \leq s_2$). We consider one specific approach, which sets $RR_{UD}(s_1, s_2) = RR_{EU}(s_1, s_2)$ to the common value $RR_U(s_1, s_2)$ that is specified log-linearly: $\log RR_U(s_1, s_2) = \gamma(s_2 - s_1)$ for $s_1 \leq s_2$. Then, for a user-selected pair of values $s_1 = s_1^{fix}$ and $s_2 = s_2^{fix}$ with $s_1^{fix} < s_2^{fix}$, we set a sensitivity parameter $RR_U(s_1^{fix}, s_2^{fix})$ to some value above one. It follows that

$$\log RR_U(s_1, s_2) = \left(\frac{s_2 - s_1}{s_2^{fix} - s_1^{fix}}\right) \log RR_U(s_1^{fix}, s_2^{fix}), \qquad s_1 \leq s_2.$$

We anchor the sieve analysis by setting $s_1 = s_1^{fix}$ at the $15^{th}$ percentile of the Day 29 antibody marker and $s_2 = s_2^{fix}$ at the $85^{th}$ percentile of the Day 29 antibody marker.

The sensitivity analysis is done for each of the two Cox model CoR analyses described in Section 12.3.2, first for tertiles of the Day 29 marker and second for the quantitative marker. For the former, E-values are reported for both the point estimate and the upper 95% confidence limit for $RR_C(0, 1)$, where category 1 is the upper tertile, category 0 is the lower tertile, and the intermediate middle tertile subgroup of vaccine recipients is excluded from the analysis. In addition, setting $RR_{UD}(0, 1) = RR_{EU}(0, 1) = 2$, such that $B(0, 1) = 4/3$, we report conservative estimation and inference on the causal risk ratio $RR_C(0, 1)$ and equivalently on the ratio of controlled vaccine efficacy curves $(1 - CVE(1))/(1 - CVE(0))$.

Next we repeat the analysis treating $S$ as a quantitative variable, where $P(T \leq t | S = s, X, A = 1)$ is again estimated by two-phase Cox partial likelihood regression and now $RR_M(s_1, s_2)$ is the marginalized risk ratio between $s_1$ and $s_2$. We will plot point and 95% confidence interval estimates of the observed marginalized risk and controlled risk curves, for the latter using the sensitivity analysis described in Section 15.1.

For validity the method requires the positivity assumption, and thus the method will only be applied if the data are reasonably supportive of the positivity assumption. To check positivity, we study the antibody marker distribution in vaccine recipients within each subgroup of the covariates $X$ that are adjusted for. For the tertiles analysis we require evidence that within each subgroup some vaccine recipients have lower tertile responses and some vaccine recipients have upper tertile responses. For the quantitative $S$ analysis, we look for evidence that $S$ varies over its full range within each level of the potential confounders that are adjusted for.

#### 15.2 CoP: Stochastic Interventional Effects on Risk and Vaccine Efficacy

Another approach to studying correlates of protection involves estimating the effect of shifting the immune response marker distribution in the vaccinated individuals (Hejazi et al., 2021). Specifically, we can consider the effect on risk of a given endpoint of a controlled intervention that shifts the distribution of an immune response by $\delta$ units, where $\delta$ is an analyst-specified real number. Considering a counterfactual scenario in which we are able to intervene so as to modify the immune response induced by the vaccine (e.g., a hypothetical change in dose or other re-formulation of the vaccine), we take this hypothetical intervention to lead to an improved (if $\delta > 0$) or lessened immune response (if $\delta < 0$) relative to the current vaccine (at $\delta = 0$). Using this framework, we can query the counterfactual risk of the endpoint under this hypothetical vaccine. Using notation established above, this quantity can be expressed as the mean of the counterfactual variable $Y(1, S(1) + \delta)$.

This approach is similar to the controlled effects approach described in Section 15.3, but with an important distinction. In the controlled effects approach, one assumes that it is possible to set $S = s$ for *all* individuals in the population. For high values of $s$, this assumption may be unrealistic if the vaccine fails to be strongly immunogenic for some subpopulations. On the other hand, with the interventional approach, it is only required that individuals' immune responses be shifted relative to their observed immune response, which may be more plausible for some vaccines.

Under assumptions (Hejazi et al., 2021), the main two of which being no unmeasured confounders and positivity (forms of both are also required for the Controlled VE CoP analyses), the counterfactual risk of interest $E[Y(1, S(1) + \delta)]$ is identified by

$$E[P(Y = 1 \mid A = 1, S = S + \delta, X = x) \mid A = 1, X] \ .$$

Examining this quantity across a range of $\delta$ provides insight into the relative contribution of a given immune response marker in preventing the endpoint of interest.

Hejazi et al. (2021) proposed nonparametric estimators that rely on estimates of the outcome regression (as described above) and the conditional density of the immune response marker in vaccinated participants. Their estimators efficiently account for two-phase sampling of immune responses and are implemented in the `txshift` package (Hejazi and Benkeser, 2020) for the `R` language and environment for statistical computing (R Core Team, 2020), available via both GitHub at
`https://github.com/nhejazi/txshift` and the Comprehensive `R` Archive Network at `https://CRAN.R-project.org/package=txshift`.

These estimators will be applied to each of the Day 29 antibody markers (without baseline adjustment) controlling for the same set of baseline risk factors that are controlled for in other analyses previously discussed. As with the mediation analysis approach described in

Section 15.3, the procedure will leverage low-dimensional risk factors alongside parametric regression strategies and flexible conditional density estimators for endpoints with fewer than 100 observed cases (pooling over the randomization arms); however, more flexible learning techniques will be employed for modeling the outcome process for endpoints with a greater number of observed cases.

In particular, conditional density estimates of immune response markers will be principally based on a nonparametric estimation strategy that reconstructs the conditional density through estimates of the conditional hazard of the discretized immune response marker values (Hejazi et al., 2021); this approach is an extension of the proposal of Díaz and van der Laan (2011). A Super Learner ensemble (van der Laan et al., 2007) of variants of this nonparametric conditional density estimator and semiparametric conditional density estimators based on Gaussinization of residuals will be constructed using the `sl3` R package (Coyle et al., 2020). In settings with limited numbers of case endpoints, the outcome process will be modeled as a Super Learner ensemble of a library of parametric regression techniques (as recommend by Gruber and van der Laan, 2010), while the library will be augmented with flexible regression techniques — including lasso and ridge regression (Tibshirani, 1996; Tikhonov and Arsenin, 1977; Hoerl and Kennard, 1970), elastic net regression (Zou and Hastie, 2003; Friedman et al., 2009), random forests (Breiman, 2001; Wright et al., 2017), extreme gradient boosting (Chen and Guestrin, 2016), multivariate adaptive polynomial and regression splines (Friedman et al., 1991; Stone et al., 1994; Kooperberg et al., 1997), and the highly adaptive lasso (van der Laan, 2017; Benkeser and van der Laan, 2016; Hejazi et al., 2020) — as the number of endpoint cases grows. These algorithm libraries will be coordinated to match those used in other CoP analyses.

Additionally, we note that $P(Y(0) = 1)$ is estimated in the same way as for the analysis of controlled vaccine efficacy, thus yielding an estimate of stochastic intervention VE defined by

$$SVE(\delta) = 1 - \frac{E[P(Y = 1 \mid A = 1, S = S + \delta, X = x) \mid A = 1, X]}{P(Y(0) = 1)}.$$

Output of the analyses will be presented as point and 95% point-wise confidence interval estimates of $E[Y(1, S(1) + \delta)]$ and of $SVE(s)$ over the values of $s$ for each of the Day 29 antibody markers, for each of a range of $\delta$ spanning -2 to 2.

Lastly, just as for the controlled VE CoP analyses, these analyses will only be performed if diagnostics support plausibility of the positivity assumption. Importantly, however, the positivity assumption for the stochastic interventional effects differs from that usually required. That is, where the positivity assumption for effects defined by static interventions requires a positive probability of treatment assignment across all strata defined by baseline factors (i.e., that a discretized immune response value be possible regardless of baseline factors), the positivity assumption of these effects is

$$s_i \in \mathcal{S} \implies s_i + \delta \in \mathcal{S} \mid A = 1, X = x$$

for all $x \in \mathcal{X}$ and $i = 1, \ldots n$. In particular, this positivity assumption does not require that the post-intervention exposure density, $q_{0,S}(S - \delta \mid A = 1, X)$, place mass across all strata defined by $X$. Instead, it requires that the post-intervention exposure mechanism be bounded, i.e.,

$$P\{q_{0,S}(S-\delta \mid A=1,X)/q_{0,S}(S \mid A=1,X)>0\}=1,$$

which may be readily satisfied by a suitable choice of $\delta$.

More importantly, the static intervention approach may require consideration of counterfactual variables that are scientifically unrealistic. Namely, it may be inconceivable to imagine a world where every participant exhibits high immune responses, given the phenotypic variability of participants' immune systems. This too may be resolved by considering an intervention $\delta(X)$, allowing the choice of $\delta$ to be a function of baseline covariates $X$ (Hejazi et al., 2021; Díaz and van der Laan, 2012; Haneuse and Rotnitzky, 2013; Díaz and van der Laan, 2018).

##### 15.3 CoP: Mediation of Vaccine Efficacy

A classic application of mediation is to decompose the overall VE into so-called *natural* direct and indirect effects. We will estimate this decomposition for each Day 29 immune marker individually, as well as when considering all immune markers together (although this SAP currently restricts to analysis of the individual markers).

For simplicity, as before, we describe this approach using a binary outcome, noting that extensions to time-to-event (with competing risks) are possible. The *total* effect of the vaccine can be represented by the risk ratio RR = (1 - VE),

$$\text{RR}=\frac{P(Y(1,S(1))=1)}{P(Y(0,S(0))=1)}\ .$$

The natural direct and indirect effects are, respectively,

$$\text{RR}_{DE}=\frac{P(Y(1,S(0))=1)}{P(Y(0,S(0))=1)} \ \text{ and } \ \text{RR}_{IDE}=\frac{P(Y(1,S(1))=1)}{P(Y(1,S(0))=1)}\ .$$

Note that $\text{RR}=\text{RR}_{DE}\text{RR}_{IDE}$, showing that the total effect decomposes into the direct times indirect effect. Another quantity of interest is the proportion mediated, which we express as

$$\text{PM}=1-\frac{\log(\text{RR}_{DE})}{\log(\text{RR})}\ .$$

We note that PM=1 if and only if $\text{RR}_{DE}=1$, i.e., no direct effect means that the marker fully mediates VE. We will estimate PM defined in this way.

As above, we must assume all confounders $X$ of $S$ and $Y$ have been measured. We also assume there is sufficient overlap of the immunologic marker distributions, and no confounders of the mediator-outcome relationship that are affected by treatment. Moreover, we require the assumption

$$P(S=s|A=0,X=x)>0 \text{ implies } P(S=s|A=1,X=x)>0 \tag{8}$$

for all subgroups $X=x$ (i.e., a.e.). Under these assumptions, $P(Y(a,S(a')=1)$ is identified by

$$E[P(Y=1 \mid A=a,S,X)|A=a',X]\ .$$

In our immune CoP application it is expected that, because the correlates analysis restricts to SARS-CoV-2 baseline seronegative individuals, the conditional density of the immune marker in the placebo arm will be a point mass at 0, that is with $S$ taking the value Negative Response. In other words, we do not expect any placebo recipients to have a positive value of the immune response marker. This implies the identification result that for $a = 0, 1$, $P(Y(a, S(0)) = 1) = E[P(Y = 1 \mid A = a, S = 0, X)]$. While $P(Y(0, S(1) = 1)$ is not identified, it is not necessary to estimate this term in order for estimation of the parameters of interest (natural direct effect, natural indirect effect, PM).

For a highly immunogenic vaccine, it may be the case that the needed overlap assumption (8) will be violated. This could happen, for example if each placebo recipient has immune marker value Negative Response (which is expected), and every vaccine recipient has immune marker value Positive Response. We will only include immune markers for mediation analysis if at least 10% of vaccine recipients have marker value equal to the value in placebo recipients.

**?** provide a multiply robust targeted minimum loss-based plug-in estimator of natural direct and indirect effects that is appropriate for case-control sampling. The estimator requires estimation of several regressions, which are used in an augmented inverse probability of treatment weighted estimator. The propensity score will be estimated by a main terms logistic regression model to account for chance imbalances across randomization arms. The sequential outcome regressions used by the approach will be based on a super learner with the 14 algorithms listed in Table 10.

Table 10: Learning Algorithms in the super learner Library for mediation methods[1].

| Algorithms | Screens[2]/ Tuning Parameters |
|---|---|
| SL.mean | All |
| SL.glm | Low-collinearity and (All, Lasso, LR) |
| SL.glm.interaction | (All, Lasso, LR) |
| SL.gam | Low-collinearity and (Lasso, LR) |
| SL.glmnet | All |
| SL.xgboost | All |
| SL.ranger | All |

[1] some nuisance parameters have binary outcomes, others quantitative. For the former, we used `family = binomial()` input to the `SuperLearner` function; for the latter, we used `family = gaussian()`.

[2] **All** = include all variables; **Lasso** = include variables with non-zero coefficients in the standard implementation of SL.glmnet that optimizes the lasso tuning parameter via 10-fold cross-validation; **Low-collinearity** = do not allow any pairs of quantitative variables with Spearman rank correlation $> 0.90$; **LR** = Univariate logistic regression Wald test 2-sided p-value $< 0.10$.

The estimator is implemented in the `natmed2` package available on GitHub (https://github.com/benkeser/natmed2). The baseline covariates $X$ adjusted for are the same as for the other analyses (i.e. of CoR and of controlled vaccine efficacy).

If there are fewer than 100 observed COVID-19 endpoint cases (pooled over the randomization arms), then we will leverage logistic and linear regression models, as appropriate, to estimate each of the above regressions and only include a low-dimensional set of pre-specified characteristics in $X$. In these cases, 95% confidence intervals for $\mathrm{RR}_{DE}$, $\mathrm{RR}_{IDE}$ and PM will be constructed using the percentile-based nonparametric bootstrap.

However, it is known that there are more than 100 COVID-19 endpoint cases (pooled over the randomization arms), a scenario for which we will instead employ super learning to estimate the above regression quantities and include a higher dimensional set of potential confounders in $X$; the same set of potential baseline potential confounders input into the superlearner modeling of the placebo arm for building a behavioral risk score. In this case, the super learner library includes a diversity of pre-specified algorithms. The nonparametric bootstrap cannot be used to construct confidence intervals, and we will instead rely on Wald-style confidence intervals with standard errors estimated based on the empirical variance of the estimators' estimated influence functions.

There may arise situations in which there is insufficient overlap of the immune response distribution. For example, if all individuals in the vaccinated arm have a Positive Response response, while all individuals in the placebo arm have a Negative Response, then the assumption (8) of the above methods will be violated. In fact, the assumption (8) implies that within every subgroup $X = x$, there needs to be some vaccinated participants with $S$ below the LOD. Therefore, for each Day 29 marker, the mediation analysis of the quantitative $S$ will only be done if diagnostics support this assumption. We will also conduct an analysis using a three-category ordinal version of $S$ with the marker threshold defining the lowest category selected to ensure that some vaccine recipients have values in the lowest category, across the subgroups; the default for this categorical $S$ will be tertiles and matched to the way that the three-category variable is analyzed for Cox model correlates of risk analysis.

See Benkeser et al. (rXiv) for additional details about the mediation method that is applied to the data.

In addition to studying all qualifying individual markers as mediators, once the complete ADCP marker data are available, the following sets of D29 markers will be assessed for their joint mediation: (1) bAb RBD, PsV ID50, ADCP; (2) bAb RBD, PsV ID50; (3) bAb RBD, ADCP; (4) PsV ID50, ADCP. Only one of the bAb markers is included given the high correlation of the bAb Spike and bAb RBD markers.

### 16 Summary of the Set of CoR and CoP Analyses and Their Requirements and Contingencies, and Synthesis of the Results, Including Reconciling Any Possible Contradictions in Results

Table 11 summarizes all of the Stage 1 / Day 29 marker correlates analyses that are done, including contingencies for whether and when each analysis is done. The quantitative version of each marker $S$, and the tertiles version of each marker $S$, is common across all of the analyses. All of the Day 29 markers are the versions that are not baseline subtracted, given that the cohort for analysis is baseline seronegative. Most of the analyses focus on univariate Day 29 markers. The primary reason to do this is the goal to identify a parsimonious correlate based on a single marker without needing to run the set of assays, and secondary reasons are: (1) the assay readouts are expected to be highly correlated, especially for the Spike bAb and RBD bAb readouts from the same MSD platform assay, and (2) there is ample precedent for univariate markers being accepted as immunological surrogate endpoints for approved vaccines (Plotkin, 2010).

Table 11: Summary of Stage 1 Day 29 Marker CoR and CoP Analyses with Requirements/Contingencies for Conduct of the Analysis

| | **Structure of** | **Requirements/Contingencies** | |
|---|---|---|---|
| **Analysis** | **Day 29 Marker(s)** | **Min No. Vaccine Endpoints** | **Other** |
| CoR Cox Model | Tertiles of $S$[1] | 25 | None |
| | Quant. $S = s$[2] | 25 | None |
| | Quant. $S \geq s$[1] | 25 | None |
| CoR Nonpar. threshold | Quant. $S \geq s$[1] | 35 | None |
| CoR GAM | Quant. $S = s$[2] | 35 | None |
| CoR Superlearner[3] | Quant. $S = s$, 2FR, 4FR | 35 | None |
| CoP: Correlates of VE | Binary $S$ | 50 | None |
| | Quant. $S = s$ | 50 | BIP with $R^2 \geq 0.25$ |
| CoP: Controlled VE[4] | Quant. $S = s$ | 50 | Feasibility of positivity[5] |
| | Tertiles of $S = s$ | 50 | Feasibility of positivity[5] |
| CoP: Stoch. Interv. VE | Quant. $S = s$ | 50 | Feasibility of positivity[5] |
| CoP: Mediators of VE | Quant. $S = s$ | 50 | Feasibility of positivity[5] |
| | Tertiles of $S$ | 50 | Feasibility of positivity[5] |

[1]These analyses are harmonized in addressing the same scientific question of how does endpoint risk vary over vaccinated subgroups defined by $S$ above a threshold.

[2]These exploratory supportive analyses are harmonized in addressing the same scientific question of how does endpoint risk vary over vaccinated subgroups defined by $S$ equal to a given marker value.

[3]Only this Superlearner analysis uses data from multiple assays and multiple readouts as input features; the other analyses consider one Day 29 biomarker at a time. Both the nonparametric monotone-constrained method and the Cox model based method are applied. [5]The positivity assumptions are as follows. Controlled VE: $P(S = s \mid A = 1, X) > 0$ almost surely. Stochastic Interventional VE: $s_i \in \mathcal{S} \implies s_i + \delta \in \mathcal{S} \mid A = 1, X = x$ for all $x \in \mathcal{X}$ and $i = 1, \ldots n$. Mediators of VE: $P(S = s \mid A = 1, X) > 0$ almost surely and $P(S = s|A = 0, X = x) >$ implies $P(S = s|A = 1, X = x) > 0$. Graphical diagnostic analyses are used to assess feasibility of each positivity assumption, where the assumption may be more feasible for $S$ as tertiles than as a quantiative variable. For quantitative $S$, the assumption is weaker for the Stochastic Interventional VE analysis, such that it is possible that only this analysis of the three will be done.

Some of the analyses include parametric assumptions for characterizing associations (Cox model and threshold analyses, Cox model versions of Controlled VE analyses) and others are nonparametric or approximately so (all other analyses). If parametric and nonparametric analyses of the same type (e.g., Cox model vs. nonparametric CoR analysis of the same association parameter; Controlled VE Cox model vs. nonparametric monotone dose-response) suggest contradictory results, then the interpretation from the nonparametric analysis will be prioritized, given it is more robust and less likely to be an incorrect result. The diagnostic testing of the parametric assumptions will aid this interpretation. As noted above, if the nonparametric analysis suggesting a contradictory result requires a positivity assumption, then its results will only be prioritized if diagnostics support feasibility of the positivity assumption.

##### 16.1 Synthesis Interpretation of Results

To structure the interpretation of the whole set of CoR and CoP results, we consider the Bradford-Hill criteria for supporting causality assessments:

1. Temporal sequence of association (vaccination causes generation of antibodies, which precede occurrence of the clinical disease outcome)
2. Strength of association (CoR magnitude)
3. Consistency of association (across studies and methods)
4. Biological gradient (may be interpreted as dose-response with greater Day 29 antibody corresponding to lower risk and greater VE)
5. Specificity (that the antibody marker is induced by vaccination not natural infection, and the antibody impacts the particular clinical endpoint being analyzed)
6. Plausibility [(supported by other COVID vaccines through study in efficacy trials and challenge (animal or human) trials, and by other potential studies such as natural history re-infection studies and monoclonal antibody prevention efficacy studies that could be challenge (animal or human) or field trials])
7. Coherence (the causality assumption does not appear to conflict with current knowledge)
8. Experimental reversibility (if VE wanes to a low level then the antibody marker also wanes coincidently; if the Day 29 marker is a strong correlate for outcome during the period of high VE, then it becomes a weaker correlate against endpoints occurring during the later period of low VE; also could be supported if vaccine breakthrough cases tend to occur early in follow-up when antibody levels are known to be relatively low)
9. Analogy (supported by other respiratory virus vaccines, and natural history studies or challenge studies of other respiratory virus vaccines)

On temporal sequence, because the analyses are done in baseline seronegative individuals, generally the Day 29 antibody responses must be generated by the vaccine, and if the outcome occurs well after Day 29, then there is clear temporal ordering of vaccination causing antibodies followed by outcome. The nuance is outcome cases with event times near 7 days post Day 29, some of which could have been infected with SARS-CoV-2 prior to Day 29 and have relatively long incubation periods, possibly perturbing temporal ordering by creating naturally-induced rather than vaccine-induced antibody. However, the knowledge about the distribution of the time period between SARS-CoV-2 acquisition and symptomatic COVID, and the time needed for an infection to create an adaptive immune response, suggests that this issue could only haves a minor impact, and overall the temporal sequence criterion readily holds. Yet, the correlates analysis that stringently only includes cases with documented antigen negativity at Day 29 may be helpful for evaluating the temporal sequence criterion.

On strength of association, this is directly quantified in all of the analyses as a core output of each method, quantified by point estimates and confidence interval estimates of covariate-adjusted association parameters or causal effect parameters.

On consistency of association, checking for similar estimates and inferences across the multiple vaccine efficacy trials will be relevant. The fact that all of the tested vaccines are designed to protect through induction of antibody to Spike protein suggest that consistency is plausible. The vaccine platform needs to be accounted for in this evaluation, where consistency may be expected for vaccines of a given type (e.g., mRNA vaccines, Spike protein vaccines, viral vector vaccines with a similar vector), whereas across types a consistent body of evidence would be very helpful, but not a requirement. FDA guidance has stipulated that a surrogate endpoint for one vaccine platform is not necessarily expected to hold for another, and that evidence for one platform would not be seen on its own as support for a surrogate endpoint for another.

In addition, we will plan to study predictiveness of the estimated optimal surrogate built on each single trial data set applied to the other trial data sets, quantified by AUC on new data sets. Moreover, consistency of association may be assessed in another sense - by studying whether the different CoR methods tend to reveal a consistent directionality and pattern of an antibody marker correlated with risk, and whether the different CoP methods tend to reveal a consistent directionality and pattern of an antibody marker connected to vaccine efficacy (as measured by the various causal effect parameters) and with different versions of vaccine efficacy. A common core element of all of the CoR and CoP methods is covariate-adjusted estimation of marker-conditional risk in vaccine recipients, e.g. of marginal conditional risk $E_X[P(T \leq t_F|S = s, A = 1, X)]$ or $E_X[P(T \leq t_F|S \geq s, A = 1, X)]$. Generally, if an estimate of this function shows strongly decreasing risk with $s$, then likely all of the CoR analyses will detect such a decrease, and the CoP analyses will detect a version of vaccine efficacy increasing in $s$. A nuance in looking for consistency of results across methods stems from the fact that different methods have different power to detect the same effect; because of this fact, consistency in magnitude (point estimate) and directionality are more important than consistency in inference/statistical significance.

The fact that all of the methods adjust for the same set of baseline covariates $X$ will aid the ability to compare the results across methods in an interpretable manner. This discussion highlights the relevance of adjusting for the same set of baseline covariates across the different efficacy trials, although our choice to do covariate-adjustment through marginalization (rather than through conditional association parameters) lends some resilience to this issue.

Our comments on consistency of association have supposed a given study endpoint, such as COVID. Another dimension of consistency evaluation could include comparing results across endpoints. On the one hand, consistency in evidence across endpoints could strengthen the case for a CoP, especially for endpoints in the same 'class' such as moderate disease and severe disease. On the other hand, the greater the difference between endpoints, the less relevant consistency may be, because the vaccine may protect through different mechanisms against each endpoint (one potential example is prevention of asymptomatic infection vs. prevention of severe disease). Thus evidence for a CoP for a given endpoint should not necessarily be down-graded based on evidence that the same marker does not appear to be a CoP for another endpoint.

On biological gradient, many of the methods are flexible and designed to detect a dose-response pattern of antibody with risk or antibody with vaccine efficacy, with tabular and graphical output of point and confidence interval estimates designed to reveal dose-response.

On specificity, as noted above antibodies generally are almost surely vaccine-induced given the analysis is done in baseline seronegative individuals, although with nuance that care is needed to evaluate whether some vaccine breakthrough cases may have had SARS-CoV-2 acquisition unusually early in follow-up (e.g., prior to second vaccination). In addition, the assays are validated for measuring specific anti-SARS-CoV-2 antigen response. Moreover, the Day 29 antibody markers can be verified to be negative in all or almost all baseline seronegative placebo recipients. Therefore, the specificity criterion should readily hold, with the proviso of the complication of the possible inclusion of unusually early infections as vaccine breakthrough cases in some analyses.

On coherence, the results will be interpreted in the light of knowledge of immune correlates of protection for the same vaccine in animal challenge studies (and human challenge studies as available), where multiple studies have demonstrated that both binding and neutralizing antibodies are a correlate of protection.

The results will also be interpreted in light of any knowledge available on passively administered SARS-CoV-2 monoclonal antibodies for prevention of SARS-CoV-2 infection or COVID disease, either in challenge studies (animals or humans) or efficacy trials. In addition, the results will be interpreted in light of results on the antibody markers as correlates of re-infection in natural history studies. Note we are cautious to not use correlates studies in already-infected individuals, because the fact of infection may readily change the nature of a correlate of protection.

On experimental reversibility, in future analyses we will evaluate whether the strength of association of the Day 29 CoRs and CoPs weakens when restricting to outcomes occurring more distal to vaccination. If the vaccine efficacy is found to wane over time, and the antibody marker wanes over time, then this decrease in the strength of association would be consistent with antibody as a correlate of protection. In contrast, if vaccine efficacy and antibody waned over time, but the strength of a Day 29 CoR and CoP was the same regardless of the timing of outcomes, it might call into question the role of the antibody marker as a CoP. The Stage 2 correlates analyses will also be helpful, where experimental reversibility could be supported simply by coincident waning of VE and waning antibody.

Experimental reversibility may also be supported by "population-level" correlates analyses, a term sometimes used in reference to meta-analysis that associates the level of VE with the population-level of a Day 29 marker across subgroups or trials; e.g. the population-level Day 29 marker response may be summarized by the geometric mean titer or geometric mean concentration. Future analyses of multiple phase 3 trial data sets will apply meta-analysis surrogate endpoint evaluation methods.

On analogy, perhaps the most relevant vaccines to consider are vaccines against other respiratory viruses, including influenza vaccine and RSV vaccines. The fact that neutralizing antibodies are a CoR and CoP for both inactivated and live virus vaccines supports that neutralizing antibodies can be a CoP for SARS-CoV-2. In addition, there is ongoing correlates of protection analysis of Novavax's Phase 3 RSV vaccine efficacy trial, that is evaluating binding antibody and neutralizing antibody CoRs and CoP correlates for severe respiratory disease in infants of vaccinated pregnant mothers (submitted). Once those results are available, they will aid in checking the analogy (and coherence) criterion.

The univariate CoR analyses assess Day 29 antibody biomarkers. The questions arise as to how do we select which biomarker seems to be the best-supported CoP, and do we need to be concerned about multiplicity adjustment issues? Given the multifactorial nature of the assessment involving biology and statistics, we for the most part avoid an approach that tries to pre-specify a quantitative ranking system; rather our approach presents the results of each marker side by side and allows human synthesis and interpretation. To guard against errors in this subjective process, we suggest that consistent results across analyses of a given trial, and consistent results (and predictive validation) across multiple trials, will provide particularly strong guidance for interpreting results. For example, if a particular Day 29 antibody marker shows remarkably consistent results in being a strong CoR and supported CoP but the other readouts do not, it may emerge as the best-supported CoP. In addition, the superlearning CoR estimated optimal surrogate objective has a special place of importance, because it includes variable importance quantification, providing some quantitative guidance on ranking the predictivneness of markers. This variable importance will be defined both internal to a given trial and based on external validation on the other efficacy trials. The metrics of CV-AUC and AUC on new trials quantifies evidence for signal in the data in a way that is protected from risk of false positive results, by virtue of having two layers of cross-validation used to estimate CV-AUC and hence avoid over-fitting. In addition, the CoR analyses use multiple hypothesis testing adjustment to help ensure clear signals and not false positive results (see Section 12.3.4). We also need a plan for minimizing the risk of false positive results for CoP analyses, which we now address.

#### 16.2 Multiple Hypothesis Testing Adjustment for CoP Analysis

For the univariable CoP analyses of the prioritized set of Day 29 antibody markers among the specified marker variables, the analysis plan seeks evidence of a CoP through four different causal effect approaches. Because of this looking for evidence through different lenses, for CoP analysis we do not focus on family-wise error rate adjustment, because FWER-adjustment aims to control the risk of making even a single false rejection. Rather, in an effort to build a body of consistent evidence and to ensure that a large fraction of that evidence is reliable, for CoP analysis we focus on false discovery rate correction. The multiplicity adjustment is performed across the Day 29 markers and across the set of CoP methods that are applied, in a single suite of hypothesis tests with calculation of FDR-adjusted p-values. As a guideline for interpreting CoP findings (but not meant to be a rigid gateway), markers with unadjusted p-value $\leq 0.05$ and FDR-adjusted p-value $\leq 0.10$ are flagged as having statistical evidence for being a CoP. These analyses will not be done until the complete marker data for the blinded phase of the study including the ADCP assay data are available.

### 17 CoP: Meta-Analysis Analysis Plan

Meta-analysis surrogate endpoint evaluation methods will be applied to the overall ENSEMBLE data set, both for assessing peak time point antibody markers (Stage 1) as surrogate endpoints for COVID and for secondary outcomes, and for assessing the antibody markers over time (Stage 2) as surrogate endpoints for COVID and for secondary outcomes. Both individual-level and trial-level meta-analysis will be applied, where the latter studies the association of vaccine effects on an antibody marker with vaccine effects on a study outcome, for example assessing how GMT nAb IU50/ml titer associates with the level of vaccine efficacy against COVID. Meta-analysis has a special role in being the only correlates approach that can potentially assess immunologic markers as CoPs that are measured using sampling types that were not stored from most trial participants (e.g., PBMC for measuring T cell responses). While the current statistical analysis plan focuses on assessing antibody markers as correlates, in the future plans may be devised to incorporate T cell response data (and potentially other data types) from phase 1-2 studies into meta-analysis evaluation.

### 18 Estimating a Threshold of Protection Based on an Established or Putative CoP (Population-Based CoP)

For each antibody marker studied as a CoP, we will apply the Chang-Kohberger Jodar et al. (2003) / Siber Siber et al. (2007) method to estimate a threshold of the antibody marker associated with the estimate of overall vaccine efficacy observed in the trial.

This method makes two simplifying assumptions: (1) that a high enough antibody marker value $s^*$ implies that individuals with $S > s^*$ have essentially zero disease risk (protection) regardless of whether they were vaccinated; and (2) $P(Y = 1|S \leq s^*, A = 1)/P(Y = 1|S \leq s^*, A = 0) = 1$ (zero vaccine efficacy if $S \leq s^*$). Based on these assumptions, $s^*$ is calculated as the value equating $1 - \widehat{P}(S \leq s^*|A = 1)/\widehat{P}(S \leq s^*|A = 0)$ to the estimate of overall vaccine efficacy. This estimate is supplemented by estimating the reverse cumulative distribution function (RCDF) of $S$ in baseline seronegative vaccine recipients and calculating a 95% confidence interval for the threshold value $s^*$ as the points of intersection of the estimated RCDF curve with the 95% confidence interval for overall vaccine efficacy (as in the figure in Andrews et al. (2014)).

This method essentially assumes that $S$ has already been established as a CoP, and under that assumption estimates a threshold that may be considered as a benchmark / study endpoint for future immunogenicity vaccine trial applications. It is acknowledged that this approach makes simplifying assumptions, namely the step-function model that unlikely holds; nonetheless it may yield a useful benchmark and complementary information on a threshold correlate of protection.

### 19 Considerations for Baseline SARS-CoV-2 Seropositive Study Participants

As stated above, if enough COVID cases in baseline seropositive vaccine and/or placebo recipients occur, then additional correlates analyses may be planned in baseline seropositive individuals. For example, the same or similar correlates of risk analysis plan that is used to analyze Day 29 marker correlates of risk in baseline seronegative vaccine recipients could be applied to assess Day 1 marker correlates of risk in baseline seropositive placebo recipients. In addition, analyses could be done to assess how vaccine efficacy in baseline seropositive participants varies with Day 1 markers. It is straightforward to make this analysis rigorous because Day 1 markers are a baseline covariate, such that regression analyses are valid based on the randomization.

### 20 Avoiding Bias with Pseudovirus Neutralization Analysis due to Use of Anti-HIV Antiretroviral Drugs

Because the lentivirus-based pseudovirus neutralization assay uses an HIV backbone, the presence of anti-retroviral drugs in serum will give a false positive neutralization signal. This can be easily screened for using an MuLV pseudotype control. Therefore, Day 1 and Day 29 samples of all study participants with data included in correlates analyses will be tested for presence of anti-retroviral drugs. Participants with any of the samples at Day 1 or Day 29 positive for antiretroviral use are excluded from analyses, for all analyses that include pseudovirus neutralization. Analyses that do not consider pseudovirus neutralization are unaffected by this issue.

### 21 Accommodating Crossover of Placebo Recipients to the Vaccine Arm

After the primary efficacy endpoint was met per the protocol-defined interim analysis, supporting the issuance on February 27, 2021 of an Emergency Use Authorization (EUA) from the FDA for the Janssen COVID-19 vaccine, Janssen COVID-19 vaccination was offered to participants who originally received placebo so that they could have the potential benefit of vaccination against COVID-19.

For crossed-over placebo recipients who have study visits and blood sample storage on the same schedule as if they had originally been assigned to the vaccine arm, follow-up data from the crossed over placebo recipients will be included in the correlates of risk analyses, which is expected to yield improved power and precision given the expanded sample size of vaccine recipients.

However, correlates of protection will only be assessed over follow-up through to the point that there is no longer a placebo cohort under blinded follow-up. Moreover, if immune marker data from crossed-over placebo recipients are available, then correlate of VE CoP analyses will be conducted that leverage the additional closeout placebo vaccination data.

The current SAP restricts to the primary blinded follow-up period.

A

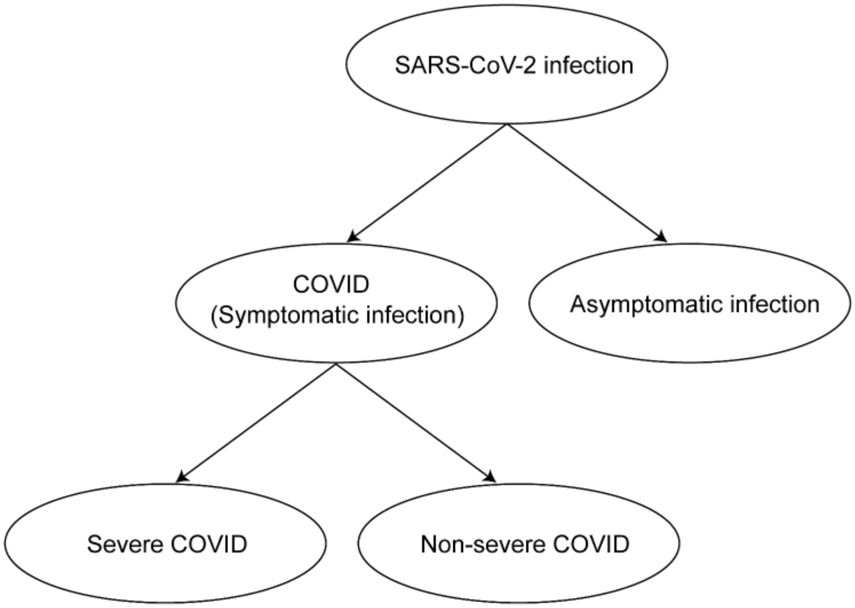

B

| **Clinical Endpoint** | **Definition** |
|---|---|
| SARS-CoV-2 infection | Positive RNA PCR test or SARS-CoV-2 seroconversion*, whichever occurs first |
| COVID (Symptomatic infection) | Meeting a protocol-specified list of COVID-19 symptoms with virological confirmation of SARS-CoV-2 infection (symptom triggered) |
| Asymptomatic infection | SARS-CoV-2 seroconversion* without prior diagnosis of the COVID endpoint¶ |
| Severe COVID | COVID endpoint with at least one protocol-specified severe disease event |
| Non-severe COVID | COVID endpoint with zero protocol-specified severe disease events |

*Seroconversion is assessed via a validated assay that distinguishes natural vs vaccine-induced SARS-CoV-2 antibodies
¶Alternatively, the asymptomatic infection endpoint can also include an RNA PCR+ test result obtained through testing regardless of symptoms (e.g., as a requirement for travel, return to school or work, or elective medical procedures) and follow-up to confirm the participant remains asymptomatic

Figure 1: A) Structural relationships among study endpoints in a COVID-19 vaccine efficacy trial. B) Study endpoint definitions.

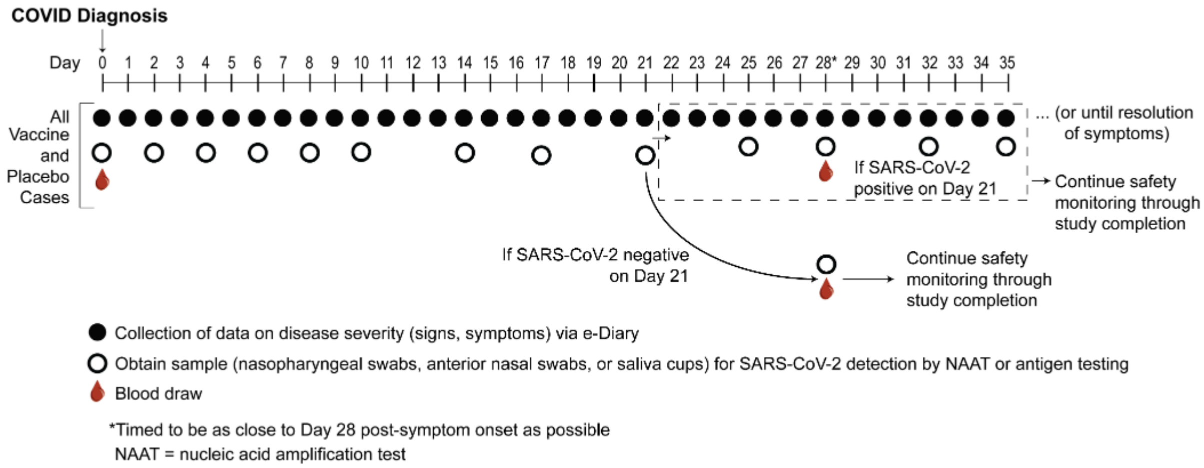

Figure 2: Example at-COVID diagnosis and post-COVID diagnosis disease severity and virologic sampling schedule, in a setting where frequent follow-up of confirmed cases can be assured. Participants diagnosed with virologically-confirmed symptomatic SARS-CoV-2 infection (COVID) enter a post-diagnosis sampling schedule to monitor viral load and COVID-related symptoms (types, severity levels, and durations).

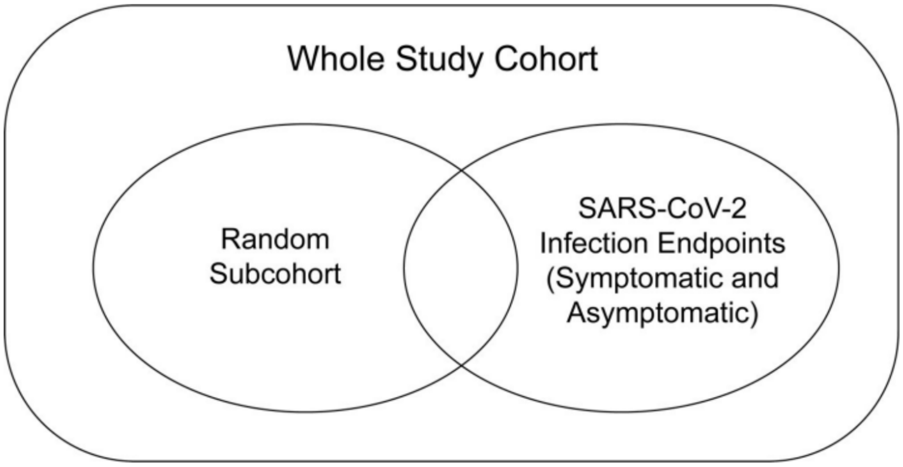

Figure 3: Case-cohort sampling design (Prentice, 1986) that measures Day 1, 29 antibody markers in all participants selected into the subcohort and in all COVID and COV-INF cases occurring outside of the subcohort.
