## Supplementary Material for "Immune Correlates Analysis of a Single Ad26.COV2.S Dose in the ENSEMBLE COVID-19 Vaccine Efficacy Clinical Trial"

**Immune Assays Team**

| Affiliation | Team Members |
|---|---|
| Biomedical Advanced Research and Development Authority (BARDA), Washington, DC | Oleg Borisov, Flora Castellino, Brett Chromy, Mark Delvecchio, Ruben O. Donis, Tremel Faison, Corey Hoffman, Christopher Houchens, Tom Hu, Pennie Hylton, Lakshmi Jayashankar, Aparna Kolhekar, James Little, Karen Martins, Jeanne Novak, Azhar Ravji, Carol Sabourin, Evan Sturtevant, Kimberly Taylor, Xiaomi Tong, John Treanor, Danielle Turley, Leah Watson, Daniel Wolfe |
| Boston Consulting Group, Boston, MA | Gian King, Andrew Li, Najaf Shah, Smruthi Suryaprakash, Jue Xiang Wang |
| Division of AIDS, NIAID, NIH, Bethesda, MD | Patricia D'Souza |
| Division of MID (Microbiology and Infectious Diseases), NIAID, NIH, Bethesda, MD | Janie Russell |
| Duke University, Durham, NC | David Beaumont, Kendall Bradley, Jiayu Chen, Xiaoju Daniell, Thomas Denny, Elizabeth Domin, Amanda Eaton, Kelsey Engel, Wenhong Feng, Juanfei Gao, Hongmei Gao, Kelli Greene, Sarah Hiles, Leihua Liu, Kristy Long, Kellen Lund, Charlene McDanal, David C. Montefiori, Marcella Sarzotti-Kelsoe, Francesca Suman, Haili Tang, Jin Tong, Olivia Widman |
| LabCorp-Monogram Biosciences, South San Francisco, CA, USA | Christos J. Petropoulos, Terri Wrin |
| The Tauri Group, an LMI company - Contract Support for U.S. Department of Defense (DOD) Joint Program Executive Office for Chemical, Biological, Radiological and Nuclear Defense (JPEO-CBRND) Joint Project Manager for Chemical, Biological, Radiological, and Nuclear Medical (JPM CBRN Medical), Fort Detrick, Maryland, USA | Christopher S. Badorrek, Gregory E. Rutkowski |
| Vaccine Research Center, NIAID, NIH, Bethesda, MD | Obrimpong Amoa-Awua, Manjula Basappa, Robin Carroll, Britta Flach, Suprabhath Gajjala, Nazaire Jean-Baptiste, Richard A. Koup, Bob C. Lin, Adrian McDermott, Christopher Moore, Mursal Naisan, Muhammed Naqvi, Sandeep Narpala, Sarah O’Connell, Clare Whittaker, Weiwei Wu, Allen Mueller, Martin Apgar, Tommy Bruington, Joe Stashick, Leo Serebryannyy, Mike Castro, Jennifer Wang |

**Janssen Team**

| Affiliation | Team Members |
|---|---|
| Janssen R&D, a division of Janssen Pharmaceutica NV, Beerse, Belgium | Sanne Roels, An Vandebosch, Carla Truyers, Frank Struyf |
| Janssen Vaccines and Prevention, Leiden, the Netherlands | Daniel J. Stieh, Mathieu Le Gars, Griet A. Van Roey, Jerald Sadoff, Jenny Hendriks, Hanneke Schuitemaker, Macaya Douoguih |

**Coronavirus Vaccine Prevention Network (CoVPN)/ENSEMBLE Team**

| Study Group Member | Affiliation | Location |
|---|---|---|
| Jerald Sadoff, M.D. | Janssen Vaccines and Prevention | Leiden, the Netherlands |
| Glenda Gray, M.B., B.Ch. | South African Research Council | Cape Town, South Africa |
| An Vandebosch, Ph.D. | Janssen Research and Development | Beerse, Belgium |
| Vicky Cárdenas, Ph.D. | Janssen Research and Development | Spring House, PA, USA |
| Georgi Shukarev, M.D. | Janssen Vaccines and Prevention | Leiden, the Netherlands |
| Beatriz Grinsztejn, M.D. | Evandro Chagas National Institute of Infectious Diseases–Fiocruz | Rio de Janeiro, Brazil |
| Paul A. Goepfert, M.D. | University of Alabama at Birmingham | Birmingham, AL, USA |
| Carla Truyers, Ph.D. | Janssen Research and Development | Beerse, Belgium |
| Hein Fennema, Ph.D. | Janssen Research and Development | Beerse, Belgium |
| Bart Spiessens, Ph.D. | Janssen Research and Development | Beerse, Belgium |
| Kim Offergeld, M.Sc. | Janssen Research and Development | Beerse, Belgium |
| Gert Scheper, Ph.D. | Janssen Vaccines and Prevention | Leiden, the Netherlands |
| Kimberly L. Taylor, Ph.D. | National Institute of Allergy and Infectious Diseases | Rockville, MD, USA |
| Merlin L. Robb, M.D. | Walter Reed Army Institute of Research | Silver Spring, MD, USA |
| John Treanor, M.D. | Biomedical Advanced Research and Development Authority | Washington, DC, USA |
| Dan H. Barouch, M.D. | Center for Virology and Vaccine Research, Beth Israel Deaconess Medical Center | Boston, MA, USA |
| Jeffrey Stoddard, M.D. | Janssen Research and Development | Raritan, NJ, USA |
| Martin F. Ryser, M.D. | Janssen Research and Development | Beerse, Belgium |
| Mary A. Marovich, M.D. | National Institute of Allergy and Infectious Diseases | Rockville, MD, USA |
| Kathleen M. Neuzil, M.D. | Center for Vaccine Development and Global Health, University of Maryland School of Medicine | Baltimore, MD, USA |
| Lawrence Corey, M.D. | Vaccine and Infectious Disease Division, Fred Hutchinson Cancer Research Center | Seattle, WA, USA |
| Nancy Cauwenberghs, Ph.D. | Janssen Research and Development | Beerse, Belgium |
| Tamzin Tanner, Ph.D. | Janssen Research and Development | Beerse, Belgium |
| Karin Hardt, Ph.D. | Janssen Research and Development | Beerse, Belgium |

| | | |
|---|---|---|
| Javier Ruiz-Guiñazú, M.D. | Janssen Research and Development | Beerse, Belgium |
| Mathieu Le Gars, Ph.D. | Janssen Vaccines and Prevention | Leiden, the Netherlands |
| Hanneke Schuitemaker, Ph.D. | Janssen Vaccines and Prevention | Leiden, the Netherlands |
| Johan Van Hoof, M.D. | Janssen Vaccines and Prevention | Leiden, the Netherlands |
| Frank Struyf, M.D. | Janssen Research and Development | Beerse, Belgium |
| Macaya Douoguih, M.D. | Janssen Vaccines and Prevention | Leiden, the Netherlands |
| Richard Gorman, MD | Biomedical Advanced Research and Development Authority (BARDA) | Washington, DC, USA |
| Carmen A. Paez, MD, MBA | Fred Hutchinson Cancer Research Center | Seattle, WA, USA |
| Edith Swann, PhD | National Institute of Allergy and Infectious Diseases (NIAID) | Rockville, MD, USA |
| James Kublin, MD, MPH | Fred Hutchinson Cancer Research Center | Seattle, WA, USA |
| Simbarashe G. Takuva, MBChB, MSc | Fred Hutchinson Cancer Research Center; University of the Witwatersrand; University of Pretoria | Seattle, WA, USA; Johannesburg, South Africa; Pretoria, South Africa |
| Alex Greninger, MD, PhD, MS, MPhil | University of Washington; Fred Hutchinson Cancer Research Center | Seattle, WA, USA |
| Pavitra Roychoudhury, PhD | University of Washington; Fred Hutchinson Cancer Research Center | Seattle, WA, USA |
| Robert W. Coombs, MD, PhD | University of Washington | Seattle, WA, USA |
| Keith R. Jerome, MD, PhD | University of Washington; Fred Hutchinson Cancer Research Center | Seattle, WA, USA |
| Flora Castellino, MD | Biomedical Advanced Research and Development Authority (BARDA) | Washington, DC, USA |
| Xiaomi Tong, PhD | Biomedical Advanced Research and Development Authority (BARDA) | Washington, DC, USA |
| Corrina Pavetto, MS, RAC | Biomedical Advanced Research and Development Authority (BARDA) | Washington, DC, USA |
| Teletha Gipson, PhD, MS | Biomedical Advanced Research and Development Authority (BARDA) | Washington, DC, USA |
| Tina Tong, DrPH(c), MS, RAC(US) | National Institute of Allergy and Infectious Diseases (NIH/NIAID) | Rockville, MD, USA |
| Marina Lee, PhD | National Institute of Allergy and Infectious Diseases (NIH/NIAID) | Rockville, MD, USA |
| James Zhou, PhD, MS | Biomedical Advanced Research and Development Authority (BARDA) | Washington, DC, USA |
| Michael Fay, PhD | National Institute of Allergy and Infectious Diseases (NIH/NIAID) | Rockville, MD, USA |

| | | |
|---|---|---|
| Kelly McQuarrie, BSN | Janssen Research & Development | Horsham, PA, USA |
| Chimeremma Nnadi, MD, PhD | Janssen Pharmaceuticals | Titusville, NJ, USA |
| Obiageli Sogbetun, MD, MPH | Janssen Infectious Disease and Vaccines | Titusville, NJ, USA |
| Nina Ahmad, MD | Janssen Pharmaceuticals | Titusville, NJ, USA |
| Ian De Proost, PhD | Johnson & Johnson | Beerse, Belgium |
| Cyrus Hoseyni, PhD | Janssen Research & Development | Spring House, PA, USA |
| Paul Coplan, ScD, MS, MBA | Johnson & Johnson Epidemiology | New Brunswick, NJ, USA |
| Najat Khan, PhD | Janssen Research & Development | New Brunswick, NJ, USA |
| Peter Ronco, BA | Janssen Research & Development | Raritan, NJ, USA |
| Dawn Furey, BA | Janssen Research & Development | Titusville, NJ, USA |
| Jodi Meck, MHA | Janssen Research & Development | Titusville, NJ, USA |
| Johan Vingerhoets, PhD | Janssen Pharmaceutica NV | Beerse, Belgium |
| Boerries Brandenburg, PhD | Janssen Vaccines & Prevention B.V. | Leiden, The Netherlands |
| Jerome Custers, PhD | Janssen Vaccines & Prevention B.V. | Leiden, The Netherlands |
| Jenny Hendriks, PhD | Janssen Vaccines & Prevention B.V. | Leiden, The Netherlands |
| Jarek Juraszek, PhD | Janssen Vaccines & Prevention B.V. | Leiden, The Netherlands |
| Anne Marit de Groot, PhD | Janssen Vaccines & Prevention B.V. | Leiden, The Netherlands |
| Griet Van Roey, PhD | Janssen Vaccines & Prevention B.V. | Leiden, The Netherlands |
| Dirk Heerwegh, PhD | Janssen Research & Development; Janssen | Beerse, Belgium |
| Ilse Van Dromme, PhD | Johnson & Johnson | Beerse, Belgium |

### CoVPN/ENSEMBLE Team, Cont'd: Participating Investigators

The following Principal Investigators participated in the ENSEMBLE study:

| Name | Institute | Location |
|---|---|---|
| Aberg, Judith | Icahn School of Medicine at Mount Sinai | New York, NY, USA |
| Adams, Mark | Central Kentucky Research Associates, Inc. | Lexington, KY, USA |
| Adams, Michael | Synexus Clinical Research US, Inc. | Murray, UT, USA |
| Aguayo, Samuel | VA Medical Center | Phoenix, AZ, USA |
| Ahsan, Habibul | The University of Chicago Medicine | Chicago, IL, USA |
| Aizenberg, Diego | Centro Médico Viamonte SRL | Buenos Aires, Argentina |
| Alonto, Augusto | Jesse Brown VAMC Department of Surgery | Chicago, IL, USA |
| Alzogaray, Maria Fernanda | Instituto Médico Platense | Buenos Aires, Argentina |
| Anderson, Evan | Emory University School of Medicine | Atlanta, GA, USA |
| Andrade Pinto, Jorge | Hospital das Clínicas da Universidade Federal de Minas Gerais | Belo Horizonte, Brazil |
| Arns da Cunha, Clóvis | Hospital Nossa Senhora das Graças | Paraná, Brazil |
| Avelino Silva, Vivian | Hospital das Clínicas da Faculdade de Medicina da USP | São Paulo, Brazil |
| Badal-Faesen, Sharlaa | University of Witwatersrand – Helen Joseph Hospital – Themba Lethu HIV Research Centre | Johannesburg, South Africa |
| Baden, Lindsey | Brigham and Women's Hospital, Inc. | Boston, MA, USA |
| Barnabas, Shaun | Family Clinical Research Unit FAM-CRU | Western Cape, South Africa |
| Bedimo, Roger | AIDS Arms Incorporated Trinity Health and Wellness Center | Dallas, TX, USA |
| Bekker, Linda-Gail | Desmond Tutu HIV Centre, University of Cape Town | Cape Town, South Africa |
| Berhe, Mezgebe | North Texas Infectious Diseases Consultants | Dallas, TX, USA |
| Bessesen, Mary | Rocky Mountain Regional VA Medical Center | Aurora, CO, USA |
| Blaser, Martin J | Rutgers Robert Wood Johnson Medical School | New Brunswick, NJ, USA |
| Bonvehi, Pablo Eduardo | CEMIC Saavedra | Buenos Aires, Argentina |
| Borger, Judith | Carolina Institute for Clinical Research | Fayetteville, NC, USA |

| | | |
|---|---|---|
| Brites Alves, Carlos Roberto | Fundação Bahiana de Infectologia | Bahia, Brazil |
| Brown, Sheldon | Bronx Veterans Affairs Medical Center | Bronx, NY, USA |
| Brumskine, William | The Aurum Institute Rustenburg Clinical Research Centre | Rustenburg, South Africa |
| Brune, Daniel | Optimal Research, LLC | Peoria, IL, USA |
| Buynak, Robert | Buynak Clinical Research | Valparaiso, IN, USA |
| Cabrera May, Carlos Antonio | Unidad de Atención Médica e Investigación en Salud (UNAMIS) | Yucatán, Mexico |
| Cadena Bonfanti, Andres Angelo | Clínica de la Costa | Barranquilla, Colombia |
| Cahn, Pedro | Fundación Huésped | Buenos Aires, Argentina |
| Carson, Jeffrey L. | Rutgers Robert Wood Johnson Medical School | New Brunswick, NJ, USA |
| Casapia Morales, Wilfredo Martin | Asociación Civil Selva Amazónica (ACSA) | Loreto, Peru |
| Cassetti, Lidia Isabel | Helios Salud S.A. | Buenos Aires, Argentina |
| Castex, Julie | Ochsner Medical Center | New Orleans, LA, USA |
| Chu, Laurence | Benchmark Research | Austin, TX, USA |
| Cotugno, Michael | Benchmark Research | Metairie, LA, USA |
| Creech, Clarence | Vanderbilt University Medical Center | Nashville, TN, USA |
| Crofoot, Gordon | CrofootMD Clinic and Research Center | Houston, TX, USA |
| Curtis, Brian | Corvallis Clinic PC | Corvallis, OR USA |
| da Silva Pilotto, José Henrique | Hospital Geral de Nova Igauçu | Rio de Janeiro, Brazil |
| Dal Ben Corradi, Mirian de Freitas | Hospital Sírio-Libanês | São Paulo, Brazil |
| Dal Pizzol, Felipe | Hospital São José | Santa Catarina, Brazil |
| Davis, Matthew | Rochester Clinical Research, Inc. | Rochester, NY, USA |
| De Carvalho Santana, Rodrigo | Hospital das Clínicas da Faculdade de Medicina de Ribeirão Preto daUSP | São Paulo, Brazil |
| de Faria Freire, Antônio Tarcísio | Santa Casa de Misericórdia de Belo Horizonte | Belo Horizonte, Brazil |
| DeJesus, Edwin | Orlando Immunology Center | Orlando, FL, USA |
| Delafontaine, Patrice | New Orleans Adolescent Trials Unit CRS | New Orleans, LA, USA |
| Delano Bronstein, Marcello | CPQuali Pesquisa Clínica LTDA ME | São Paulo, Brazil |
| Dell'Italia, Louis | University of Alabama at Birmingham | Birmingham, AL, USA |

| | | |
|---|---|---|
| Deluca, Mercedes | Clinical Trials Division – Stamboulian Servicios de Salud | Buenos Aires, Argentina |
| Denham, Douglas | Clinical Trials of Texas, Inc. | San Antonio, TX, USA |
| Diacon, Andreas | TASK Central | Western Cape, South Africa |
| Dubula, Thozama | Nelson Mandela Academic Clinical Research Unit (NeMACRU) | Mthatha, South Africa |
| Eder, Frank | Meridian Clinical Research, LLC | Endwell, NY, USA |
| Edupuganti, Srilatha | The Hope Clinic at Emory University | Decatur, GA, USA |
| Ervin, John | The Center for Pharmaceutical Research | Kansas City, MO, USA |
| Esteves Coelho, Lara | FIO CRUZ – Fundação Oswaldo Cruz – Inst de Pesquisa Clínica Evandro Chagas | Rio de Janeiro, Brazil |
| Eudes Leal, Fabio | Universidade Municipal de São Caetano do Sul | São Paulo, Brazil |
| Fairlie, Lee | Shandukani Research Centre | Gauteng, South Africa |
| Fierro, Carlos | Johnson County Clin-Trials | Lenexa, KS, USA |
| Fogarty, Charles | Spartanburg Medical Research | Spartanburg, SC, USA |
| Fragoso, Veronica | Texas Center for Drug Development, Inc. | Houston, TX, USA |
| Frank, Ian | University of Pennsylvania | Philadelphia, PA, USA |
| Frey, Sharon | Saint Louis University | St Louis, MO, USA |
| Gallardo Cartagena, Jorge | Centro de Investigaciones Tecnológicas, Biomédicas y Medioambientales (CITBM) | Lima, Peru |
| Gamarra Ayarza, Cesar Augusto | Centro de Investigaciones Médicas | Callao, Peru |
| Garcia Diaz, Julia | Ochsner Medical Center | New Orleans, LA, USA |
| Gaur, Aditya | St Jude Children's Research Hospital | Memphis, TN, USA |
| Gentile, Nina | Temple University Hospital | Philadelphia, PA, USA |
| Gill, Katherine | Masiphumelele Research Centre | Cape Town, South Africa |
| Gonzalez, Alexander | MedPlus Medicina Prepagadá S.A. | Bogotá, Colombia |
| Gottlieb, Robert | Baylor Scott & White Research Institute | Dallas, TX, USA |
| Grant, Philip | Stanford University Medical Center | Palo Alto, CA, USA |
| Greenberg, Richard | University of Kentucky | Lexington, KY, USA |
| Greiwe, Cathy | Synexus Clinical Research US, Inc. | Columbus, OH, USA |

| | | |
|---|---|---|
| Guedes Barbosa, Luiz Sergio | Oncovida – Centro de Onco-Hematologia de Mato Grosso | Mato Grosso, Brazil |
| Han-Conrad, Laurie | WR-MCCR, LCC | San Diego, CA, USA |
| Hidalgo Vidal, Jose Alfredo | Asociación Civil Vía Libre | Lima, Peru |
| Higuera Cobos, Juan Diego | Fundación Oftalmológica de Santander – FOSCAL | Santander, Colombia |
| Hong, Matthew | Wake Research Associates | Raleigh, NJ, USA |
| Innes, James Craig | The Aurum Institute Klerksdorp Clinical Research Centre | Klerksdorp, South Africa |
| Jackson, Lisa | Kaiser Permanente Washington Health Research Institute | Seattle, WA, USA |
| Jackson-Booth, Peta-Gay | Optimal Research, LLC | Rockville, MD, USA |
| Jaller Raad, Juan Jose | Centro de Reumatología y Ortopedia | Barranquilla, Colombia |
| Jayaweera, Dushyantha | University of Miami – Miller School of Medicine | Miami, FL, USA |
| Jennings, William | Synexus Clinical Research US, Inc. | San Antonio, TX, USA |
| João Filho, Esaú Custódio | Hospital Federal dos Servidores do Estado | Rio de Janeiro, Brazil |
| Kassim, Sheetal | Desmond Tutu HIV Foundation – University of Cape Town | Cape Town, South Africa |
| Kennelly, Christina | Tryon Medical Partners | Charlotte, NC, USA |
| Khetan, Shishir | Meridian Clinical Research, LLC | Rockville, MD, USA |
| Kilgore, Paul E. | Henry Ford Health System | Detroit, MI, USA |
| Kim, Kenneth | Ark Clinical Research | Long Beach, CA, USA |
| Kirby, William | Synexus Clinical Research US, Inc. | Birmingham, AL, USA |
| Kopp, James | Synexus Clinical Research US, Inc. | Anderson, SC, USA |
| Kotze, Philip | Qhakaza Mbokodo Research Clinic | KwaZulu-Natal, South Africa |
| Kotze, Sheena | Stanza Clinical Research Centre: Mamelodi | Gauteng, South Africa |
| Kriesel, John | University of Utah | Salt Lake City, UT, USA |
| Kutner, Mark | Suncoast Research Group | Miami, FL, USA |
| Lacerda Nogueira, Maurício | Fundação Faculdade Regional de Medicina de São José do Rio Preto | São Paulo, Brazil |
| Laher, Fatima | Perinatal HIV Research Unit, Chris Hani Baragwanath Hospital | Gauteng, South Africa |
| Lama Valdivia, Javier Ricardo | Asociación Civil Impacta Salud y Educación – Barranco | Lima, Peru |
| Lazarus, Erica | Perinatal HIV Research Unit (PHRU), Kliptown | Soweto, South Africa |

| | | |
|---|---|---|
| Lazcano Ponce, Eduardo Cesar | Instituto Nacional de Salud Pública | Morelos, Mexico |
| Leibman, Daniel | VA Medical Center | Columbia, SC, USA |
| Levin, Michael | Clinical Research Center of Nevada | Las Vegas, NV, USA |
| Levin, Myron | Children's Hospital Colorado | Aurora, CO, USA |
| Little, Susan | UCSD AntiViral Research Center (AVRC) | San Diego, CA, USA |
| Lombaard, Johannes | Josha Research | Bloemfontein, South Africa |
| Lopez Medina, Eduardo | Centro de Investigaciones Clínicas S.A.S. | Cali, Colombia |
| Losso, Marcelo | Hospital J.M. Ramos Mejía | Buenos Aires, Argentina |
| Luabeya, Angelique | SATVI, Brewelskloof Hospital | Western Cape, South Africa |
| Lucksinger, Gregg | Clinical Research Institute of Southern Oregon, P.C. | Medford, OR, USA |
| Lugogo, Njira | University of Michigan Neurosurgery A. Alfred Taubman Health CareCenter | Ann Arbor, MI, USA |
| Luz, Kleber Giovanni | Centro de Estudos e Pesquisas em Moléstias Infecciosas | Rio Grande do Norte, Brazil |
| Maboa, Rebone | Ndlovu Elandsdoorn Site | Limpopo, Dennilton, South Africa |
| Macareno Arroyo, Hugo Andres | Hospital Universidad del Norte | Barranquilla, Colombia |
| Makhaza, Disebo | CAPRISA Vulindlela Clinic | KwaZulu-Natal, South Africa |
| Malahleha, Mookho | Setshaba Research Centre | Soshanguve, South Africa |
| Malan, Daniel | PHOENIX Pharma (Pty) Ltd. | Eastern Cape, South Africa |
| Mamba, Musawenkosi | CRISMO Bertha Gxowa Research Centre | Gauteng, South Africa |
| Manning, Mary Beth | Rapid Medical Research | Cleveland, OH, USA |
| Martin, Judith | University of Pittsburgh | Pittsburgh, PA, USA |
| Mauricio da Silva, Cesar | Faculdade de Medicina Barretos – FACISB | Barretos, SP, Brazil |
| McGettigan, John | Quality of Life Medical & Research Center, LLC | Tucson, AZ, USA |
| Medrano Allende, Juan | Clínica y Maternidad Suizo Argentina | Buenos Aires, Argentina |
| Mena, Leandro | University of Mississippi Medical Center | Jackson, MS, USA |
| Messer, William | Oregon Health & Science University | Portland, OR, USA |
| Middleton, Randle | Optimal Research, LLC | Huntsville, AL, USA |

| | | |
|---|---|---|
| Mills, Anthony | Anthony Mills Medical, Inc. | Los Angeles, CA, USA |
| Mills, Richard | PMG Research of Charleston, LLC | Mount Pleasant, SC, USA |
| Mngadi, Kathryn | The Aurum Institute: Tembisa – Clinic 4 | Tembisa, South Africa |
| Moanna, Abeer | VA Medical Center – Atlanta | Decatur, GA, USA |
| Mofsen, Ricky | Massachusetts General Hospital | Boston, MA, USA |
| Moncada Vilela, Zandra | Hospital Nacional Arzobispo Loayza | Lima, Peru |
| Montaña, Oscar Romano | DIM Clínica Privada | Buenos Aires, Argentina |
| Moreno Hoyos Abril, Juan | Hospital Universitario de Nuevo León 'Dr. José Eleuterio González' | Nuevo León, Mexico |
| Morse, Caryn | Wake Forest Baptist Medical Center | Winston-Salem, NC, USA |
| Muñoz Reyes, Manuel | Hospital Dr. Hernán Henríquez Aravena | Araucanía, Chile |
| Murray, Linda | Synexus Clinical Research US, Inc. | Pinellas Park, FL, USA |
| Naicker, Nivashnee | Centre for the AIDS Programme of Research in South Africa | KwaZulu-Natal, South Africa |
| Naicker, Vimla | South Africa Medical Research Council | KwaZulu-Natal, South Africa |
| Naidoo, Logashvari | South African Medical Research Council Chatsworth Clinical ResearchSite | KwaZulu-Natal, South Africa |
| Nchabeleng, Maphoshane | MeCRU Clinical Research Unit | Gauteng, South Africa |
| Newman Lobato Souza, Tamara | Instituto de Infectologia Emilio Ribas | São Paulo, Brazil |
| Novak, Richard | University of Illinois at Chicago | Chicago, IL, USA |
| Nugent, Paul | Synexus Clinical Research US, Inc. | Cincinnati, OH, USA |
| O'Ryan Gallardo, Miguel Luis | Facultad de Medicina, Universidad de Chile | Santiago, Chile |
| Oyanguren Miranda, Martin | Hospital Nacional Edgardo Rebagliati Martins | Lima, Peru |
| Padala, Kalpana | Central Arkansas Veterans Healthcare System | Little Rock, AR, USA |
| Panettieri, Jr., Reynold A. | Rutgers Robert Wood Johnson Medical School | New Brunswick, NJ, USA |
| Panjwani, Sameer G. | Rush University Medical Center | Chicago, IL, USA |
| Patelli Juliani Souza Lima, Maria | Hospital e Maternidade Celso Pierro | São Paulo, Brazil |
| Pelkey, Leslie | Cherry Street Services, Inc. | Grand Rapids, MI, USA |
| Petrick, Friedrich | Mzansi Ethical Research Centre | Middelburg, South Africa |
| Pounds, Kevin | Synexus Clinical Research US, Inc. | Tucson, AZ, USA |

| | | |
|---|---|---|
| Powell, Richard | New Horizons Clinical Research | Cincinnati, OH, USA |
| Pragalos, Antoinette | CTI Clinical Trial and Consulting Services | Cincinnati, OH, USA |
| Pratley, Richard E. | AdventHealth Orlando | Orlando, FL, USA |
| Presti, Rachel | Washington University School of Medicine | St Louis, MO, USA |
| Ramalho Madruga, José Valdez | Centro de Referência e Treinamento DST/AIDS | São Paulo, Brazil |
| Ramesh, Mayur | Henry Ford Health System | Detroit, MI, USA |
| Ramirez Sanchez, Isabel Cristina | Hospital Pablo Tobón Uribe | Antioquia, Colombia |
| Ramirez, Julio | University of Louisville | Louisville, KY, USA |
| Rankin, Bruce | Avail Clinical Research, LLC | DeLand, FL, USA |
| Restrepo, Jaime | Fundación Centro de Investigación Clinícа (CIC) | Medellín, Colombia |
| Reynales Londono, Humberto | Centro de Atención e Investigación Médica S.A. – CAIMED | Bogotá, Colombia |
| Reynolds, Michele | Synexus Clinical Research US, Inc. | Dallas, TX, USA |
| Rhame, Frank | Abbott Northwestern Hospital Clinic | Minneapolis, MN, USA |
| Rhee, Margaret | Synexus Clinical Research US, Inc. | Akron, OH, USA |
| Riddle, Mark | VA Sierra Nevada Health Care System | Reno, NV, USA |
| Riegel Santos, Breno | Hospital Nossa Senhora da Conceição (HNSC) | Rio Grande do Sul, Brazil |
| Riffer, Ernie | Central Phoenix Medical Clinic | Phoenix, AZ, USA |
| Rizzardi, Barbara | Advanced Clinical Research | West Jordan, UT, USA |
| Rosso, Fernando | Fundación Valle del Lili | Valle del Cauca, Colombia |
| Saavedra, Carla | Bioclinica Santiago Bulnes | Santiago, Chile |
| Safirstein, Beth | MD Clinical | Hallandale Beach, FL, USA |
| Scapellato, Pablo | CEMEDIC | Buenos Aires, Argentina |
| Scheinberg, Phillip | Real e Benemérita Associação Portuguesa de Beneficência | São Paulo, Brazil |
| Schwartz, Howard | Research Centers of America, LLC | Hollywood, FL, USA |
| Sedillo, David J. | Rush University Medical Center | Chicago, IL, USA |
| Servilla, Karen | Raymond G. Murphy VA Medical Center | Albuquerque, NM, USA |
| Shah, Raj | Rush University Medical Center | Chicago, IL, USA |

| | | |
|---|---|---|
| Siegel, Amy | MediSync Clinical Research | Petal, MS, USA |
| Silva Orellana, Rafael | Clínica del Maule | Talca, Chile |
| Silva, Federico | Fundación Cardiovascular de Colombia – Instituto del Corazón Floridablanca | Floridablanca, Colombia |
| Smith, Steven R. | AdventHealth Orlando | Orlando, FL, USA |
| Spooner, Elizabeth | South Africa Medical Research Council | KwaZulu-Natal, South Africa |
| Sprinz, Eduardo | Hospital de Clínicas de Porto Alegre | Porto Alegre, Brazil |
| Sriram, Peruvemba | North Florida/South Georgia Veterans Health System | Gainesville, FL, USA |
| Strout, Cynthia | Coastal Carolina Research Center, Inc. | Mount Pleasant, SC, USA |
| Swiatlo, Edwin | Southeast Louisiana Veterans Health Care System | New Orleans, LA, USA |
| Taiwo, Babafemi | Northwestern University | Evanston, IL, USA |
| Talwani, Rohit | Baltimore VA Medical Center | Baltimore, MD, USA |
| Tavares Russo, Luís Augusto | Instituto Brasil de Pesquisa Clínica | Rio de Janeiro, Brazil |
| Terront Lozano, Monica Alexandra | Solano y Terront Servicios Médicos Ltda | Bogotá, Colombia |
| Tharenos, Leslie | Synexus Clinical Research US, Inc. | St Louis, MO, USA |
| Tien, Phyllis | VA Medical Center | San Francisco, CA, USA |
| Tieu, Hong Van | New York Blood Center | New York, NY, USA |
| Toney, John | James A. Haley VA Hospital GNS | Tampa, FL, USA |
| Urbach, Dorothea | Synexus Helderberg Clinical Research Centre | Western Cape, South Africa |
| Vachris, Timothy | Optimal Research, LLC | Austin, TX, USA |
| Valencia, Javier | Asociación Civil Impacta Salud y Educación – San Miguel CRS | Lima, Peru |
| van Nieuwenhuizen, Elane | Synexus Watermeyer | Gauteng, South Africa |
| Vannucci Lomonte, Andrea | CEPIC – Centro Paulista de Investigação Clínica | São Paulo, Brazil |
| Vasconcellos, Eduardo | Instituto de Pesquisas Clínicas | Distrito Federal, Brazil |
| Velez, Ivan Dario | Programa de Estudio y Control de Enfermedades Tropicales | Medellín, Colombia |
| Ward, Amy | University of Cape Town IDM/CIDRI Research Site | Cape Town, South Africa |
| Wells, Cassia | Harlem Hospital Center | New York, NY, USA |

| | | |
|---|---|---|
| White, Judith | Synexus Clinical Research US, Inc. | Orlando, FL, USA |
| Winkle, Peter | Anaheim Clinical Trials, LLC | Anaheim, CA, USA |
| Woods, Christopher | Durham VAMC | Raleigh, NC, USA |
| Zaidman, Cesar Javier | CIPREC | Buenos Aires, Argentina |

**United States Government (USG)/Coronavirus Prevention Network (CoVPN) Biostatistics Team**

| Affiliation | Team Members |
|---|---|
| Biomedical Advanced Research and Development Authority (BARDA), Washington, DC | Di Lu, James Zhou |
| Department of Biostatistics and Bioinformatics, Rollins School of Public Health, Emory University | David Benkeser, Sohail Nizam |
| Vaccine and Infectious Disease Division, Fred Hutchinson Cancer Center, Seattle, WA | Jessica Andriesen, Bhavesh Borate, Lindsay N. Carpp, Andrew Fiore-Gartland, Youyi Fong*, Peter B. Gilbert*, Ying Huang*, Yunda Huang*, Ellis Hughes, Ollivier Hyrien, Holly E. Janes*, Michal Juraska, Yiwen Lu, April K. Randhawa, Brian Simpkins, Brian D. Williamson*, Lars W.P. van der Laan, Chenchen Yu |
| Biostatistics Research Branch, NIAID, NIH, Bethesda, MD | Michael P. Fay, Dean Follmann, Martha Nason |
| Department of Biostatistics, University of Washington, Seattle, WA | Marco Carone, Avi Kenny, Kendrick Li, Wenbo Zhang |
| Department of Statistics, University of Washington, Seattle, WA | Alex Luedtke |
| Division of Biostatistics, School of Public Health, Department of Population Health Sciences, Weill Cornell Medicine, New York, New York | Nima S. Hejazi |
| Department of Population Health Sciences, Weill Cornell Medical College, New York, New York | Iván Díaz |

*YF, PBG, YiH, and HEJ are also affiliated with the Department of Biostatistics, University of Washington, Seattle, WA. PBG and YuH are also affiliated with the Public Health Sciences Division, Fred Hutchinson Cancer Research Center, Seattle, WA. YuH is also affiliated with the Department of Global Health, University of Washington, Seattle, WA. Brian D. Williamson is also affiliated with Kaiser Permanente Washington Health Research Institute, Seattle, Washington, USA.

**Extended Data Figure 1. Case-cohort set and trial timeline. A)** Case-cohort set. **B-D)** Distribution of participants in the case-cohort set by geographic region: **B)** Latin America, **C)** South Africa, **D)** US. **E)** Phases of the ENSEMBLE trial, timing of Ad26.COV2.S dose and serum sampling, and time period for diagnosis of the COVID-19 endpoint.

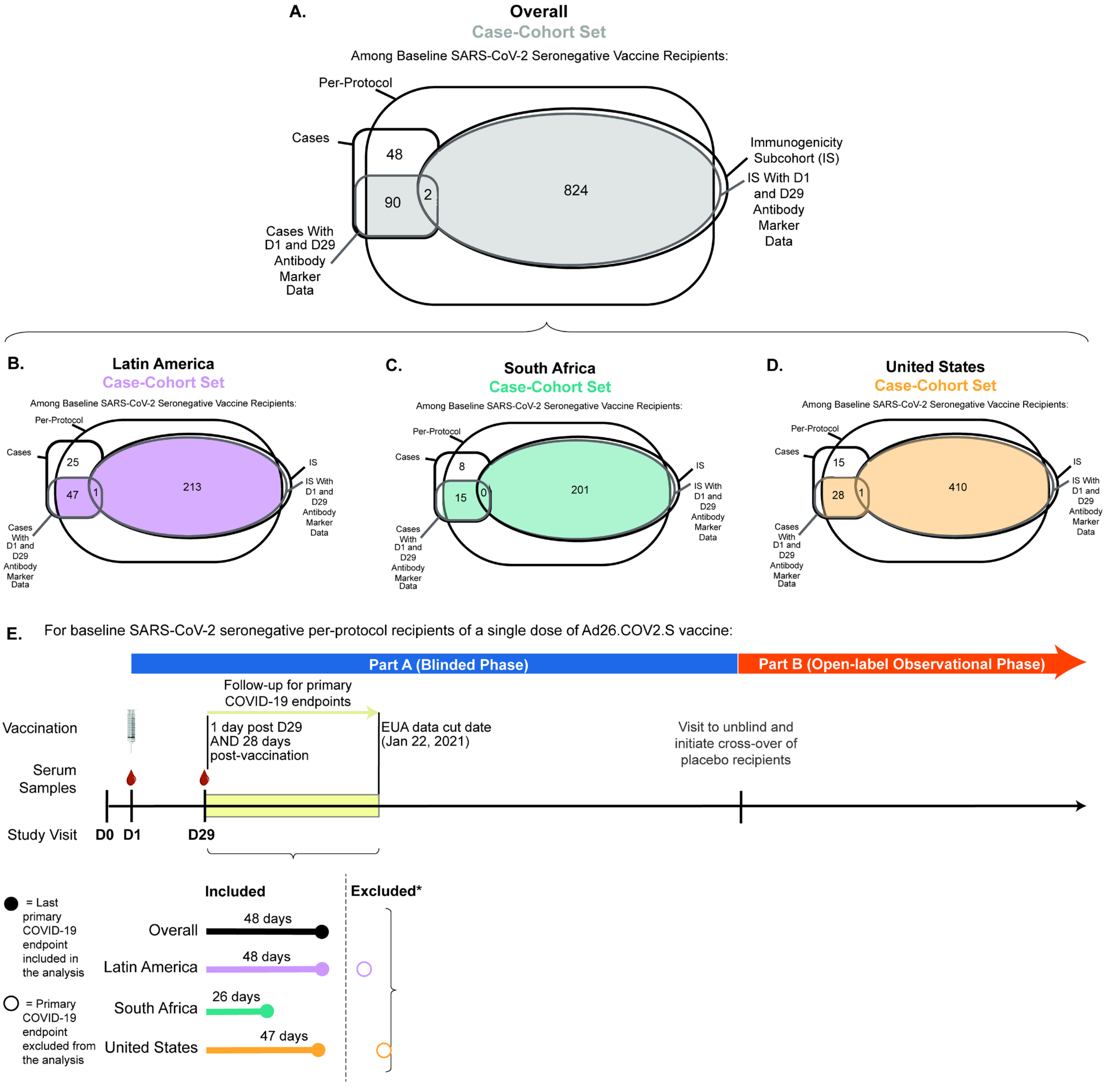


*Two primary COVID-19 endpoints beyond 54 days post D29 study visit (one in Latin America at 58 days post D29 visit and one in the United States at 66 days post D29 visit) were excluded due to small numbers of subcohort participants at-risk in this right tail of follow-up time

**Extended Data Figure 2. Flowchart of study participants from enrollment to the case-cohort set of baseline SARS-CoV-2 seronegative per-protocol participants.**

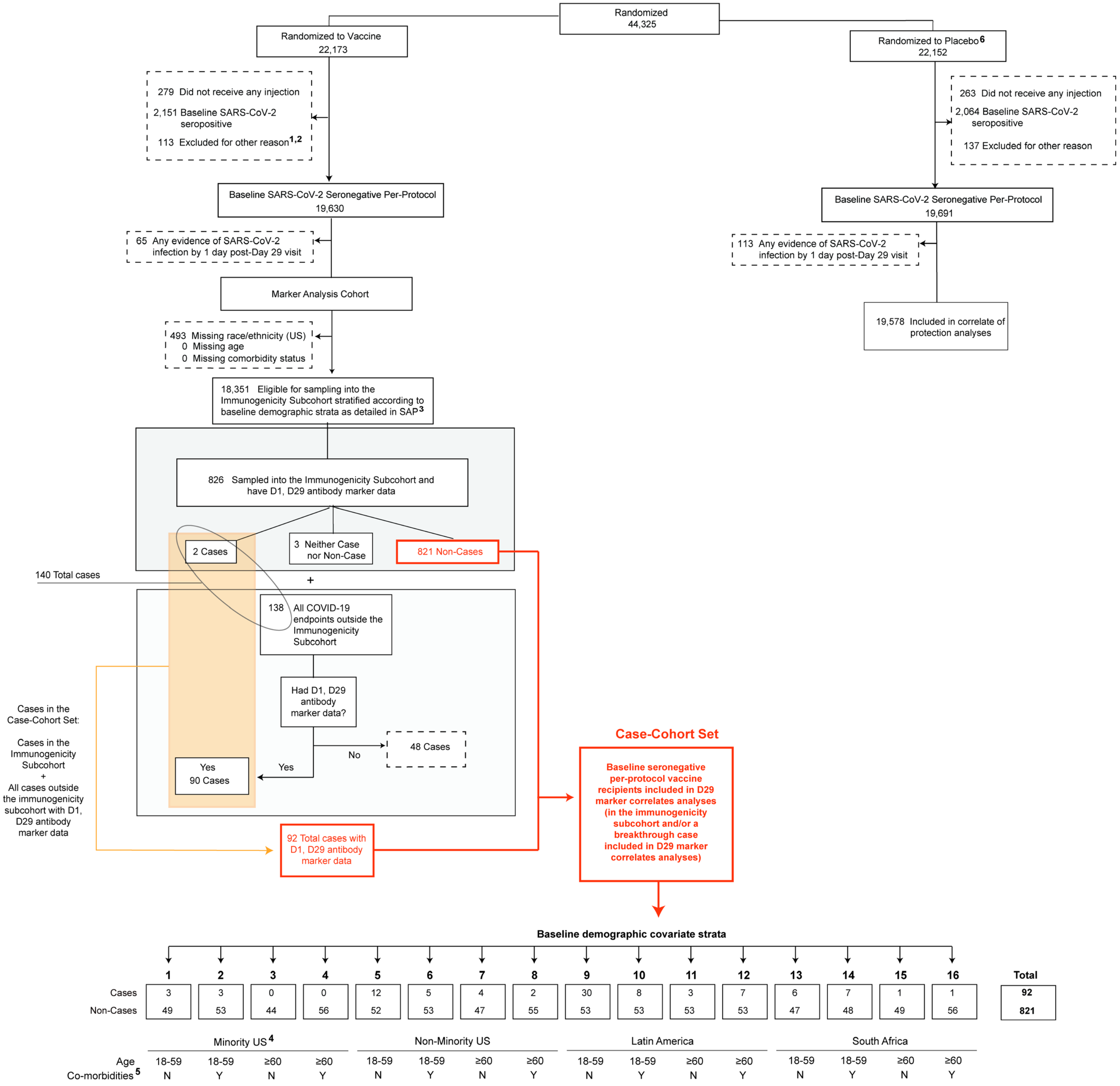


1 Participants could have been excluded for more than one reason.
2 Other reasons for exclusion included RT-PCR positive at baseline, violated inclusion or exclusion criteria, received wrong vaccine or incorrect dose, received disallowed concomitant medication, or other.
3 See the SAP for details.
4 Minority is defined as the complement of being known to be White Non-Hispanic. White Non-Hispanic is defined as Race=White and Ethnicity=Not Hispanic or Latino. All other Race subgroups are defined as Black, Asian, American Indian or Alaska Native, Native Hawaiian or Other Pacific Islander, Multiracial, Other, Not reported, or Unknown. (In Latin America, the American Indian or Alaska Native category was labeled as "Indigenous South American".) Minority status was only reported in the U.S.
5 Co-morbidities are listed in Table S4 of Sadoff et al. 2021, NEJM and consisted of conditions that have been associated with increased risk of severe COVID-19.
6 Placebo recipient ptids are used in the correlate of protection analyses. However, antibody data from the placebo arm are not used in correlates analyses, given no variability in values; they were only used to verify low false positive rates of the immunoassays.

**Extended Data Figure 3. D29 antibody marker level in participants in Latin America by COVID-19 outcome status.** (A) Anti-spike IgG concentration, (B) anti-receptor binding domain (RBD) IgG concentration, and (C) pseudovirus (PsV) neutralization ID50 titer. Data points are from the Latin America subgroup of baseline SARS-CoV-2 seronegative per-protocol vaccine recipients in the set. The violin plots contain interior box plots with upper and lower horizontal edges the 25th and 75th percentiles of antibody level and middle line the 50th percentile, and vertical bars the distance from the 25th (or 75th) percentile of antibody level and the minimum (or maximum) antibody level within the 25th (or 75th) percentile of antibody level minus (or plus) 1.5 times the interquartile range. Each side shows a rotated probability density (estimated by a kernel density estimator with a default Gaussian kernel) of the data. Positive response rates were computed with inverse probability of sampling weighting. Pos.Cut, Positivity cut-off. Positive response for spike IgG was defined by IgG > 10.8424 BAU/ml and for RBD IgG was defined by IgG > 14.0858 BAU/ml. ULoQ, upper limit of quantitation. ULoQ = 238.1165 BAU/ml for spike IgG and 172.5755 BAU/ml for RBD IgG. LLoQ, lower limit of quantitation. Positive response for ID50 was defined by value > LLoQ (2.7426 IU50/ml). ULoQ = 619.3052 IU50/ml for ID50. Cases are baseline SARS-CoV-2 seronegative per-protocol vaccine recipients with the primary COVID-19 endpoint (moderate to severe-critical COVID-19 with onset both ≥ 1 day post D29 and ≥ 28 days post-vaccination) up to 54 days post D29 but no later than January 22, 2021.

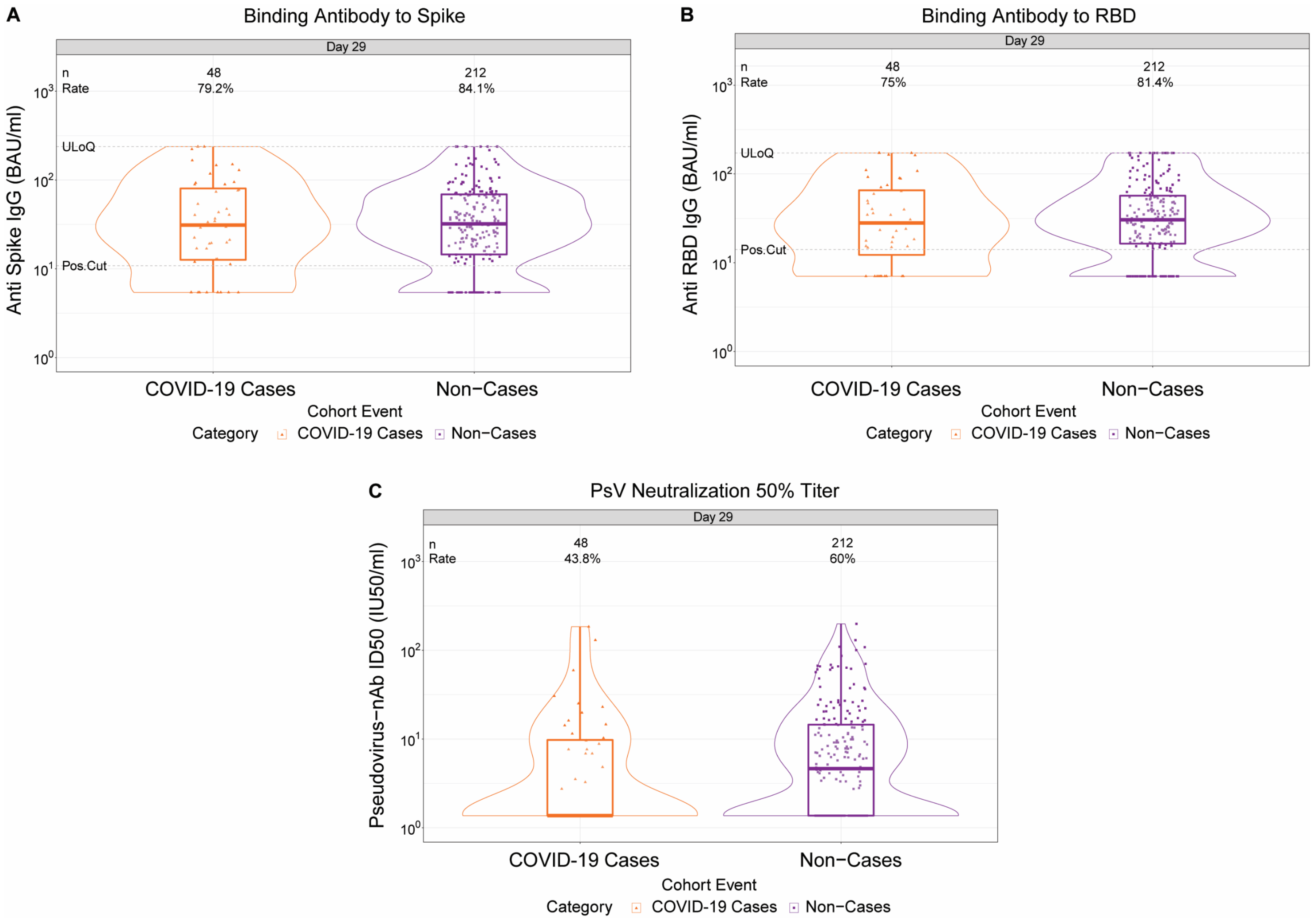

**Extended Data Figure 4. D29 antibody marker level in participants in South Africa by COVID-19 outcome status.** (A) Anti-spike IgG concentration, (B) anti-receptor binding domain (RBD) IgG concentration, and (C) pseudovirus (PsV) neutralization ID50 titer. Data points are from the South Africa subgroup of baseline SARS-CoV-2 seronegative per-protocol vaccine recipients in the set. The violin plots contain interior box plots with upper and lower horizontal edges the 25th and 75th percentiles of antibody level and middle line the 50th percentile, and vertical bars the distance from the 25th (or 75th) percentile of antibody level and the minimum (or maximum) antibody level within the 25th (or 75th) percentile of antibody level minus (or plus) 1.5 times the interquartile range. Each side shows a rotated probability density (estimated by a kernel density estimator with a default Gaussian kernel) of the data. Positive response rates were computed with inverse probability of sampling weighting. Pos.Cut, Positivity cut-off. Positive response for spike IgG was defined by IgG > 10.8424 BAU/ml and for RBD IgG was defined by IgG > 14.0858 BAU/ml. ULoQ, upper limit of quantitation. ULoQ = 238.1165 BAU/ml for spike IgG and 172.5755 BAU/ml for RBD IgG. LLoQ, lower limit of quantitation. Positive response for ID50 was defined by value > LLoQ (2.7426 IU50/ml). ULoQ = 619.3052 IU50/ml for ID50. Cases are baseline SARS-CoV-2 seronegative per-protocol vaccine recipients with the primary COVID-19 endpoint (moderate to severe-critical COVID-19 with onset both ≥ 1 day post D29 and ≥ 28 days post-vaccination) up to 54 days post D29 but no later than January 22, 2021.

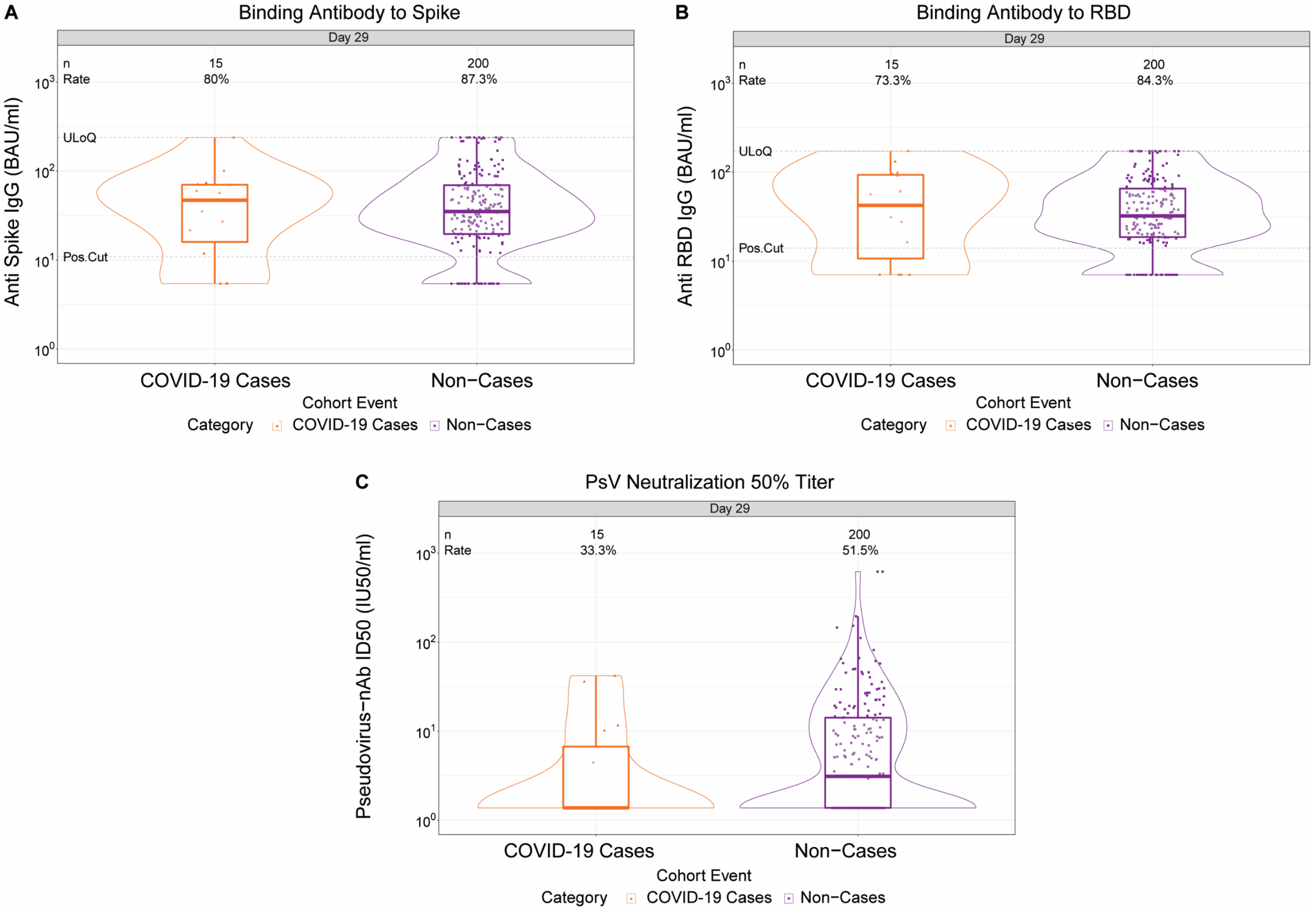

**Extended Data Figure 5. D29 antibody marker level in participants in the United States by COVID-19 outcome status.** (A) Anti-spike IgG concentration, (B) anti-receptor binding domain (RBD) IgG concentration, and (C) pseudovirus neutralization ID50 titer. Data points are from the United States subgroup of baseline SARS-CoV-2 seronegative per-protocol vaccine recipients in the set. The violin plots contain interior box plots with upper and lower horizontal edges the 25th and 75th percentiles of antibody level and middle line the 50th percentile, and vertical bars the distance from the 25th (or 75th) percentile of antibody level and the minimum (or maximum) antibody level within the 25th (or 75th) percentile of antibody level minus (or plus) 1.5 times the interquartile range. Each side shows a rotated probability density (estimated by a kernel density estimator with a default Gaussian kernel) of the data. Positive response rates were computed with inverse probability of sampling weighting. Pos.Cut, Positivity cut-off. Positive response for spike IgG was defined by IgG > 10.8424 BAU/ml and for RBD IgG was defined by IgG > 14.0858 BAU/ml. ULoQ, upper limit of quantitation. ULoQ = 238.1165 BAU/ml for spike IgG and 172.5755 BAU/ml for RBD IgG. LLoQ, lower limit of quantitation. Positive response for ID50 was defined by value > LLoQ (2.7426 IU50/ml). ULoQ = 619.3052 IU50/ml for ID50. Cases are baseline SARS-CoV-2 seronegative per-protocol vaccine recipients with the primary COVID-19 endpoint (moderate to severe-critical COVID-19 with onset both ≥ 1 day post D29 and ≥ 28 days post-vaccination) up to 54 days post D29 but no later than January 22, 2021.

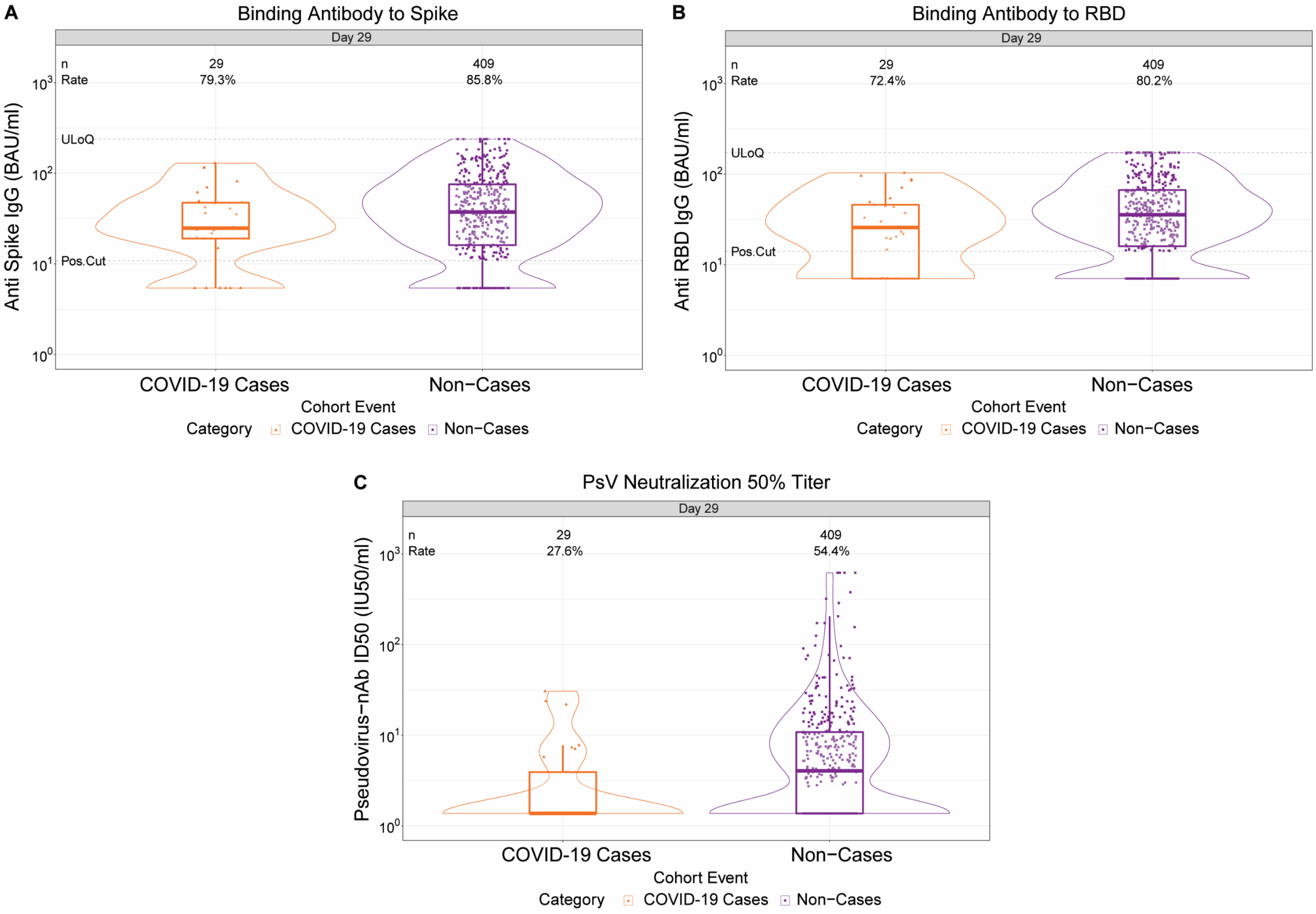

**Extended Data Figure 6. Correlations of D29 antibody markers in baseline SARS-CoV-2 seronegative per-protocol vaccine recipients in the immunogenicity subcohort. A)** Scatterplot of receptor binding domain (RBD) IgG and spike IgG; **B)** Scatterplot of RBD IgG and PsV-nAb ID50; **C)** Scatterplot of spike IgG and PsV-nAb ID50. Cor = Spearman rank correlation.

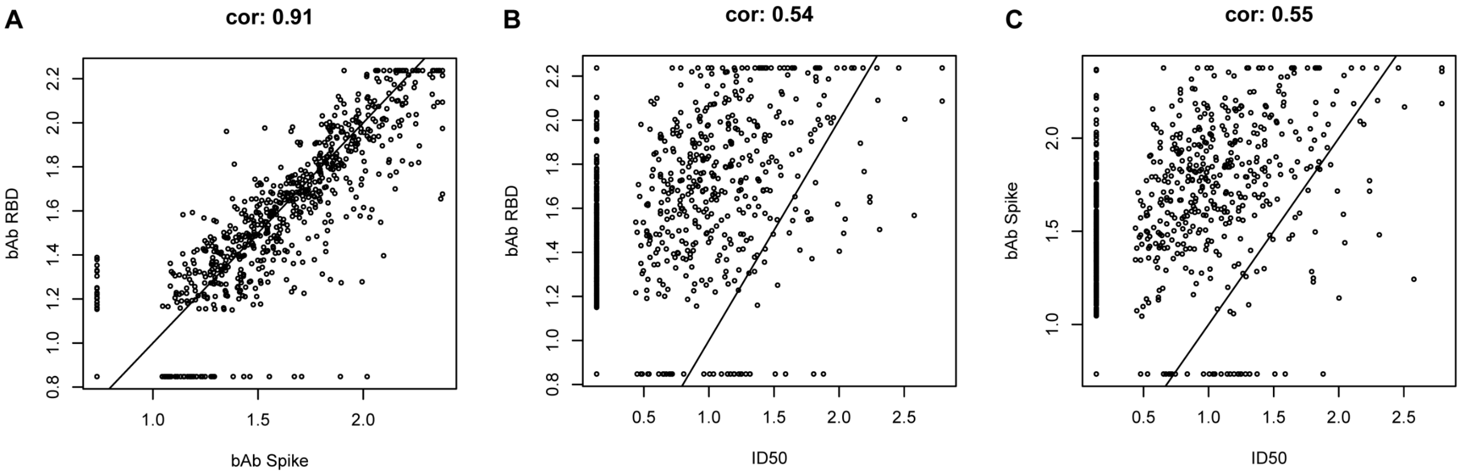

**Extended Data Figure 7. Antibody marker values [spike IgG, receptor binding domain (RBD) IgG, ID50] by COVID-19 outcome status in placebo recipients.** Data points are from baseline SARS-CoV-2 seronegative per-protocol placebo recipients in the case-cohort set. The violin plots contain interior box plots with upper and lower horizontal edges the 25th and 75th percentiles of antibody level and middle line the 50th percentile, and vertical bars the distance from the 25th (or 75th) percentile of antibody level and the minimum (or maximum) antibody level within the 25th (or 75th) percentile of antibody level minus (or plus) 1.5 times the interquartile range. Each side shows a rotated probability density (estimated by a kernel density estimator with a default Gaussian kernel) of the data. Positive response rates were computed with inverse probability of sampling weighting. Pos.Cut, Positivity cut-off. Positive response for spike IgG was defined by IgG > 10.8424 BAU/ml and for RBD IgG was defined by IgG > 14.0858 BAU/ml. ULoQ, upper limit of quantitation. ULoQ = 238.1165 BAU/ml for spike IgG and 172.5755 for RBD IgG. LLoQ, lower limit of quantification. Positive response for ID50 was defined by value > LLoQ (2.7426 IU50/ml). ULoQ = 619.3052 IU50/ml for ID50. Cases are baseline SARS-CoV-2 seronegative per-protocol vaccine recipients with the primary COVID-19 endpoint (moderate to severe-critical COVID-19 with onset both ≥ 1 day post D29 and ≥ 28 days post-vaccination) up to 54 days post D29 but no later than January 22, 2021.

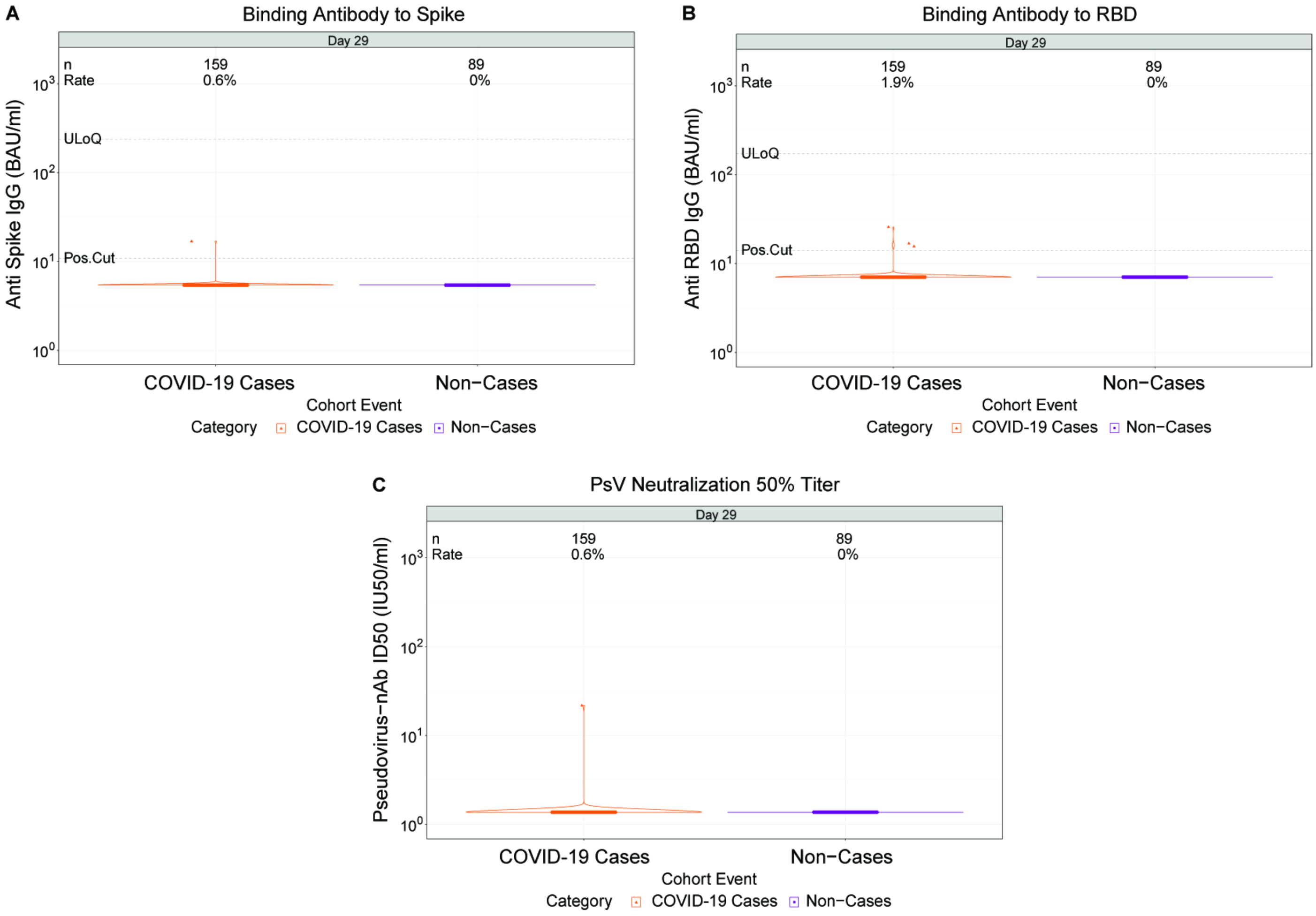

**Extended Data Figure 8. Analyses of spike IgG and receptor binding domain (RBD) IgG as correlates of risk and as correlates of protection.** Analyses were peformed in baseline SARS-CoV-2 seronegative per-protocol vaccine recipients. **A, B)** Covariate-adjusted cumulative incidence of COVID-19 by 54 days post D29 by vaccinated baseline SARS-CoV-2 seronegative per-protocol subgroups defined by D29 **(A)** anti-spike IgG or **(B)** anti-RBD IgG concentration above a threshold, with reverse cumulative distribution function (CDF) of D29 marker level overlaid in green. The blue dots are point estimates at each COVID-19 primary endpoint linearly interpolated by solid black lines; the gray shaded area is pointwise 95% confidence intervals (CIs). The estimates and CIs were adjusted using the assumption that the true threshold-response is nonincreasing. The upper boundary of the green shaded area is the estimate of the reverse cumulative distribution function (CDF) of D29 marker level in baseline SARS-CoV-2 seronegative per-protocol vaccine recipients. The vertical red dashed line is the D29 marker threshold above which no COVID-19 endpoints were observed (in the time frame of 1 through 54 days post D29). **C, D)** Covariate-adjusted cumulative incidence of COVID-19 by 54 days post D29 by D29 **(C)** anti-spike IgG or **(D)** anti-RBD IgG concentration, estimated using (solid purple line) a Cox model or (solid blue line) a nonparametric method. The dotted black lines indicate bootstrap point-wise 95% CIs. The upper and lower horizontal gray lines are the overall cumulative incidence of COVID-19 from 1 to 54 days post D29 in placebo and vaccine recipients, respectively. **E, F)** Vaccine efficacy (solid purple line) by D29 **(E)** anti-spike IgG or **(F)** anti-RBD IgG concentration, estimated using a Cox proportional hazards implementation of [1]. The dashed black lines indicate bootstrap point-wise 95% CIs. Vaccine efficacy (solid blue line) by D29 **(E)** anti-spike IgG concentration or **(F)** anti-RBD IgG concentration, estimated using a nonparametric implementation of Gilbert et al.[1] (see SAP). The blue shaded area represents the 95% CIs. In **C-F**, curves are plotted over the range from Positivity Cut-off/2 to the 97.5th percentile = (**C**, **E**) 238 BAU/ml for Spike IgG or (**D**, **F**) 173 BAU/ml for RBD IgG; In **C-F**, the green histogram is an estimate of the density of D29 marker and the horizontal gray line is the overall vaccine efficacy from 1 to 54 days post D29, with the dotted gray lines indicating the 95% CIs. Baseline covariates adjusted for were baseline risk score and geographic region.

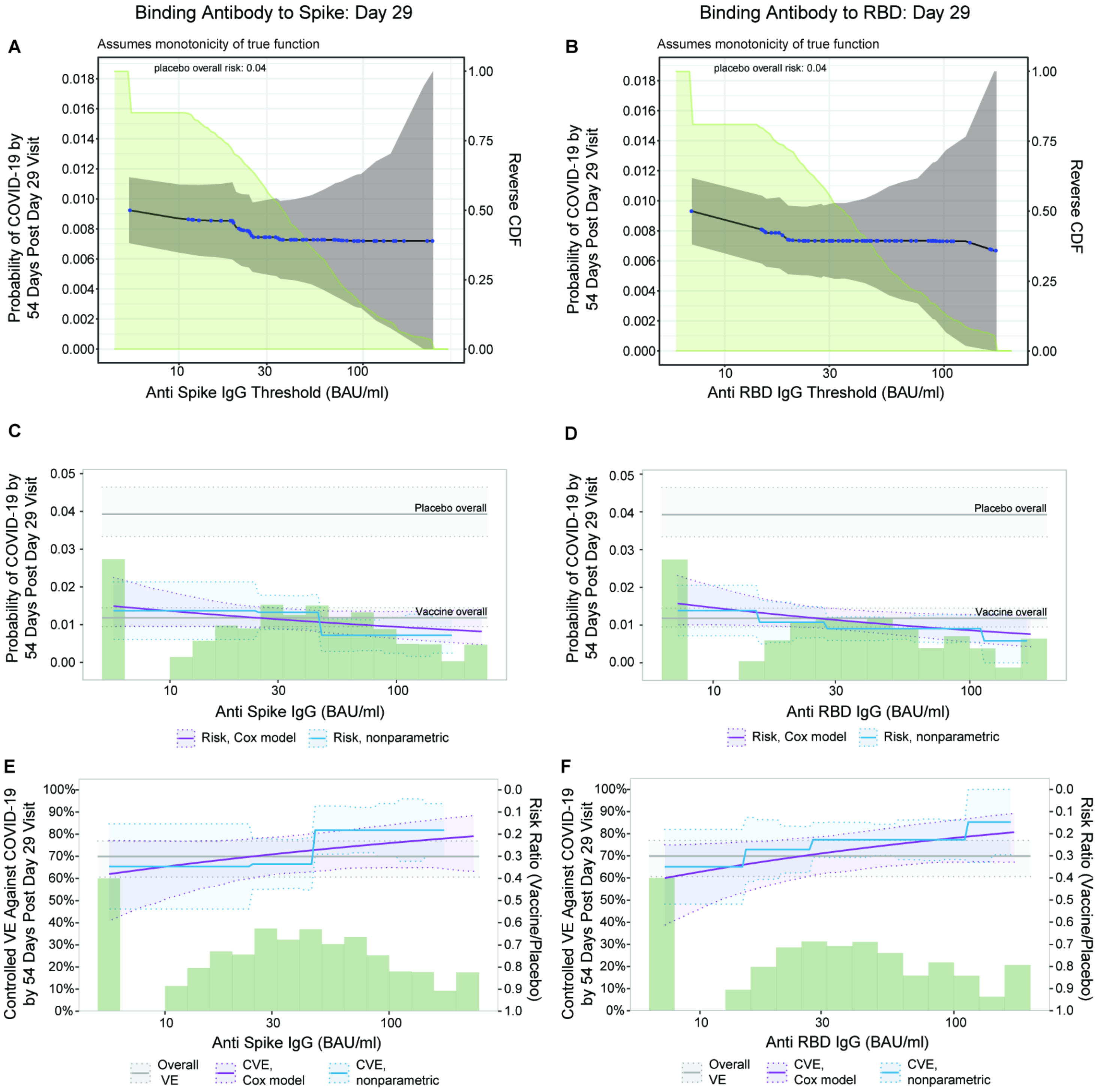

Binding Antibody to Spike: Day 29
Binding Antibody to RBD: Day 29
A
Assumes monotonicity of true function
placebo overall risk: 0.04
Probability of COVID-19 by 54 Days Post Day 29 Visit
Reverse CDF
Anti Spike IgG Threshold (BAU/ml)
B
Assumes monotonicity of true function
placebo overall risk: 0.04
Probability of COVID-19 by 54 Days Post Day 29 Visit
Reverse CDF
Anti RBD IgG Threshold (BAU/ml)
C
Probability of COVID-19 by 54 Days Post Day 29 Visit
Placebo overall
Vaccine overall
Anti Spike IgG (BAU/ml)
Risk, Cox model
Risk, nonparametric
D
Probability of COVID-19 by 54 Days Post Day 29 Visit
Placebo overall
Vaccine overall
Anti RBD IgG (BAU/ml)
Risk, Cox model
Risk, nonparametric
E
Controlled VE Against COVID-19 by 54 Days Post Day 29 Visit
Risk Ratio (Vaccine/Placebo)
Anti Spike IgG (BAU/ml)
Overall VE
CVE, Cox model
CVE, nonparametric
F
Controlled VE Against COVID-19 by 54 Days Post Day 29 Visit
Risk Ratio (Vaccine/Placebo)
Anti RBD IgG (BAU/ml)
Overall VE
CVE, Cox model
CVE, nonparametric

**Extended Data Figure 9. Vaccine efficacy with sensitivity analysis by D29 (A) anti-spike IgG concentration, (B) anti-receptor binding domain (RBD) IgG concentration, or (C) pseudovirus (PsV) neutralization ID50 titer.** Vaccine efficacy estimates were obtained a Cox proportional hazards implementation of ref.[1]. The upper boundary of the green shaded area is the estimate of the reverse cumulative distribution function of the marker in baseline SARS-CoV-2 seronegative per-protocol vaccine recipients. The pink solid line is point estimates assuming no unmeasured confounding; the dashed lines are bootstrap point-wise 95% CIs. The red solid line is point estimates assuming unmeasured confounding in a sensitivity analysis (dashed lines are bootstrap point-wise 95% CIs); see the SAP section 15.1 for details of the sensitivity analysis. The horizontal gray line is the overall vaccine efficacy from 1 to 54 days post D29, with the dotted gray lines indicating the 95% CIs. All curves are plotted over the range from (A, B) Positivity Cut-off/2 to the 97.5th percentile = 238 BAU/ml for Spike IgG or 173 BAU/ml for RBD IgG; (C) LLOQ/2 to the 97.5th percentile = 96.3 IU50/ml for PsV nAb ID50.

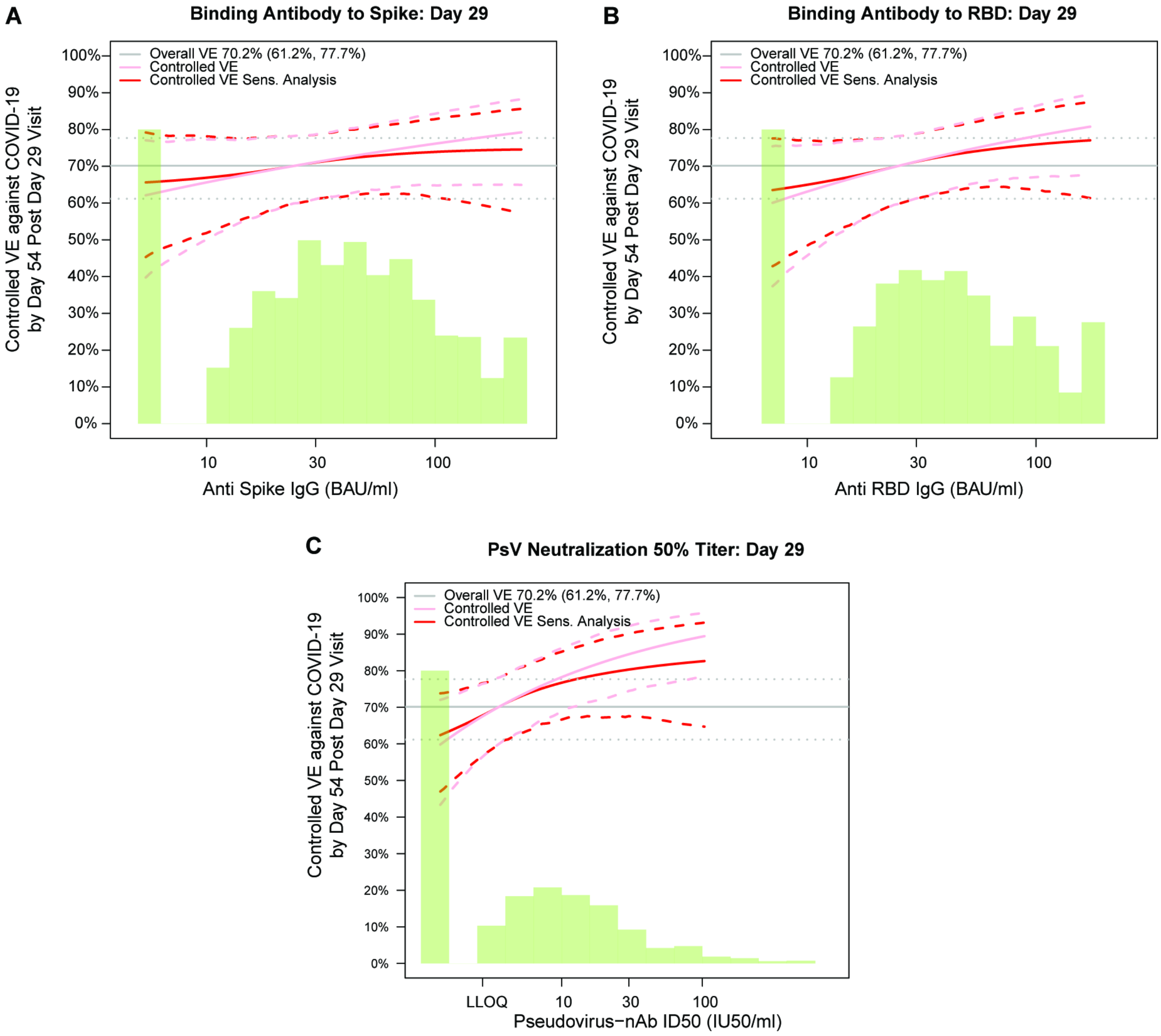

**Extended Data Figure 10.** Vaccine efficacy (solid lines) in baseline SARS-CoV-2 seronegative per-protocol vaccine recipients by A) D29 spike IgG or B) D29 receptor binding domain (RBD) IgG in ENSEMBLE by geographic region (US, United States; Lat Am, Latin America; S Afr, South Africa), estimated using the Cox proportional hazards implementation of Gilbert et al.[1] The dashed lines indicate bootstrap point-wise 95% CIs. The follow-up periods for the VE assessment were: A) ENSEMBLE-US, 1 to 53 days post D29; ENSEMBLE-Lat Am, 1 to 48 days post D29; ENSEMBLE-S Afr, 1 to 40 days post D29. The green histograms are an estimate of the density of D29 marker level by geographic region. Baseline covariates adjusted for were baseline risk score and geographic region. Curves are plotted over the range from Positivity Cut-off/2 to the 97.5th percentile = (**A**) 238 BAU/ml for Spike IgG; (**B**) 173 BAU/ml for RBD IgG.

**A**

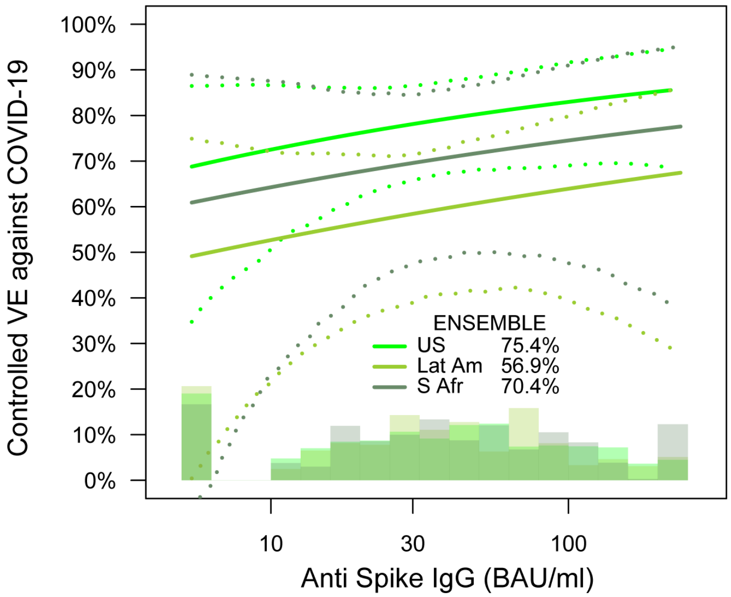


**B**

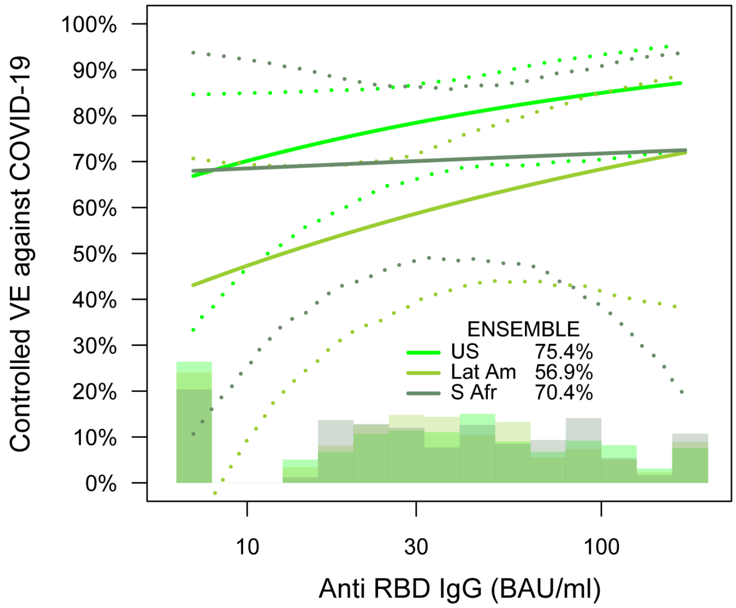

**Supplementary Table 1. Sample sizes of baseline SARS-CoV-2 seronegative per-protocol vaccine recipients included in immune correlates analyses, by baseline sampling strata and case/non-case strata.**

Case-cohort set = Baseline SARS-CoV-2 seronegative per-protocol vaccine recipients included in D29 marker correlates analysis [in the immunogenicity subcohort (IS) and/or a breakthrough COVID-19 case)]*

| | Baseline Sampling Strata of Baseline SARS-CoV-2 Seronegative Per-Protocol Vaccine Participants Included in Correlates Analyses | | | | | | | | | | | | | | | | | | | |
|---|---|---|---|---|---|---|---|---|---|---|---|---|---|---|---|---|---|---|---|---|
| | | | | | | | | | | | | | | | | | | Subtotals | | |
| | 1 | 2 | 3 | 4 | 5 | 6 | 7 | 8 | 9 | 10 | 11 | 12 | 13 | 14 | 15 | 16 | Total | US | Lat Am | S Afr |
| Breakthrough COVID-19 cases (both within and outwith the IS) with Ab marker data | 3 | 3 | 0 | 0 | 12 | 5 | 4 | 2 | 30 | 8 | 3 | 7 | 6 | 7 | 1 | 1 | 92 | 29 | 48 | 15 |
| Non-cases in the IS with Ab marker data | 49 | 53 | 44 | 56 | 52 | 53 | 47 | 55 | 53 | 53 | 53 | 53 | 47 | 48 | 49 | 56 | 821 | 409 | 212 | 200 |

Ab, antibody; IS, immunogenicity subcohort; Lat Am, Latin America; S Afr, South Africa; US, United States.

Demographic covariate strata:

1. Minority U.S., age 18-59, comorbidities N
2. Minority U.S., age 18-59, comorbidities Y
3. Minority U.S., age ≥ 60, comorbidities N
4. Minority U.S., age ≥ 60, comorbidities Y
5. non-Minority U.S., age 18-59, comorbidities N
6. non-Minority U.S., age 18-59, comorbidities Y
7. non-Minority U.S., age ≥ 60, comorbidities N
8. non-Minority U.S., age ≥ 60, comorbidities Y
9. Latin America, age 18-59, comorbidities N
10. Latin America, age 18-59, comorbidities Y
11. Latin America, age ≥ 60, comorbidities N
12. Latin America, age ≥ 60, comorbidities Y
13. South Africa, age 18-59, comorbidities N
14. South Africa, age 18-59, comorbidities Y
15. South Africa, age ≥ 60, comorbidities N
16. South Africa, age ≥ 60, comorbidities Y

Minority is defined as the complement of being known to be White Non-Hispanic. Minority status was only reported in the U.S.
White Non-Hispanic is defined as Race=White and Ethnicity=Not Hispanic or Latino. All other Race subgroups are defined as Black, Asian, American Indian or Alaska Native, Native Hawaiian or Other Pacific Islander, Multiracial, Other, Not reported, or Unknown. (In Latin America, the American Indian or Alaska Native category was labeled as "Indigenous South American".)

Cases are baseline SARS-CoV-2 seronegative per-protocol vaccine recipients with the primary COVID-19 endpoint (moderate to severe-critical COVID-19 with onset that was both ≥ 28 days post-vaccination and ≥ 1 day post-D29) through to the data cut (January 22, 2021).
Non-cases/Controls are baseline seronegative per-protocol vaccine recipients sampled into the immunogenicity subcohort with no evidence of SARS-CoV-2 infection up to the end of the correlates study period, which is up to 54 days post D29 but no later than the data cut (January 22, 2021).

**Supplementary Table 2. Demographics and clinical characteristics of baseline SARS-CoV-2 seronegative per-protocol trial participants in the immunogenicity subcohort and thus have D1, D29 antibody data.**

| Characteristics | Vaccine (N = 826) | Placebo (N = 90) | Total (N = 916) |
|---|---|---|---|
| **Age** | | | |
| Age 18-59 | 412 (49.9%) | 42 (46.7%) | 454 (49.6%) |
| Age ≥ 60 | 414 (50.1%) | 48 (53.3%) | 462 (50.4%) |
| Mean (Range) | 49.4 (18.0, 80.0) | 51.8 (18.0, 80.0) | 49.7 (18.0, 80.0) |
| **BMI** | | | |
| Underweight BMI < 18.5 | 18 (2.2%) | 2 (2.2%) | 20 (2.2%) |
| Normal 18.5 ≤ BMI < 25 | 230 (27.8%) | 20 (22.2%) | 250 (27.3%) |
| Overweight 25 ≤ BMI < 30 | 294 (35.6%) | 34 (37.8%) | 328 (35.8%) |
| Obese BMI ≥ 30 | 284 (34.4%) | 34 (37.8%) | 318 (34.7%) |
| **Risk for Severe COVID-19** | | | |
| At-risk | 429 (51.9%) | 45 (50.0%) | 474 (51.7%) |
| Not at-risk | 397 (48.1%) | 46 (50.0%) | 442 (48.3%) |
| **Age, Risk for Severe COVID-19** | | | |
| Age 18-59 At-risk | 208 (25.2%) | 21 (23.3%) | 229 (25.0%) |
| Age 18-59 Not at-risk | 204 (24.7%) | 21 (23.3%) | 225 (24.6%) |
| Age ≥ 60 At-risk | 221 (26.8%) | 24 (26.7%) | 245 (26.7%) |
| Age ≥ 60 Not at-risk | 193 (23.4%) | 24 (26.7%) | 217 (23.7%) |
| **Sex Assigned at Birth** | | | |
| Female | 373 (45.2%) | 37 (41.1%) | 410 (44.8%) |
| Male | 453 (54.8%) | 53 (58.9%) | 506 (55.2%) |
| **Hispanic or Latino Ethnicity** | | | |
| Hispanic or Latino | 307 (37.2%) | 32 (35.6%) | 339 (37.0%) |
| Not Hispanic or Latino | 492 (59.6%) | 54 (60.0%) | 546 (59.6%) |
| Not reported and unknown | 27 (3.3%) | 4 (4.4%) | 31 (3.4%) |
| **Race** | | | |
| White | 434 (52.5%) | 48 (53.3%) | 482 (52.6%) |
| Black or African American | 245 (29.7%) | 26 (28.9%) | 271 (29.6%) |
| Asian | 15 (1.8%) | | 15 (1.6%) |
| American Indian or Alaska Native | 58 (7.0%) | 6 (6.7%) | 64 (7.0%) |
| Native Hawaiian or Other Pacific Islander | 3 (0.4%) | | 3 (0.3%) |
| Multiracial | 40 (4.8%) | 7 (7.8%) | 47 (5.1%) |
| Not reported and unknown | 31 (3.8%) | 3 (3.3%) | 34 (3.7%) |
| **Underrepresented Minority Status in the U.S.*** | | | |
| White Non-Hispanic | 208 (25.2%) | 24 (26.7%) | 232 (25.3%) |
| Communities of Color | 203 (24.6%) | 23 (25.6%) | 226 (24.7%) |
| **Country** | | | |
| United States | 411 (49.8%) | 47 (52.2%) | 458 (50.0%) |
| Argentina | 33 (4.0%) | 2 (2.2%) | 35 (3.8%) |

| | | | |
|---|---|---|---|
| Brazil | 89 (10.8%) | 12 (13.3%) | 101 (11.0%) |
| Chile | 11 (1.3%) | 3 (3.3%) | 14 (1.5%) |
| Colombia | 53 (6.4%) | 5 (5.6%) | 58 (6.3%) |
| Mexico | 7 (0.8%) | 2 (2.2%) | 9 (1.0%) |
| Peru | 21 (2.5%) | 1 (1.1%) | 22 (2.4%) |
| South Africa | 201 (24.3%) | 18 (20.0%) | 219 (23.9%) |
| **HIV Status** | | | |
| Negative | 798 (96.6%) | 88 (97.8%) | 886 (96.7%) |
| Living with HIV | 28 (3.4%) | 2 (2.2%) | 30 (3.3%) |

*Data on minority status was only gathered in the U.S.

This table summarizes characteristics of per-protocol participants in the immunogenicity subcohort, which was randomly sampled from the study cohort. The sampling was stratified by strata defined by enrollment characteristics: Assigned randomization arm × Baseline SARS-CoV-2 seronegative vs. seropositive × Randomization strata. The U.S. subcohort includes 8 baseline demographic strata; the Latin America and South Africa subcohorts each include 4 baseline demographic strata.

**Supplementary Table 3. Demographics and clinical characteristics of baseline SARS-CoV-2 seronegative per-protocol trial participants in the United States subset of the immunogenicity subcohort (strata 1-8) and thus have D1, D29 antibody data.**

| Characteristics | Vaccine (N = 411) | Placebo (N = 47) | Total (N = 458) |
|---|---|---|---|
| **Age** | | | |
| Age 18-59 | 209 (50.9%) | 24 (51.1%) | 233 (50.9%) |
| Age ≥ 60 | 202 (49.1%) | 23 (48.9%) | 225 (49.1%) |
| Mean (Range) | 48.9 (18.0, 80.0) | 52.0 (18.0, 80.0) | 49.3 (18.0, 80.0) |
| **BMI** | | | |
| Underweight BMI < 18.5 | 1 (0.2%) | 1 (2.1%) | 2 (0.4%) |
| Normal 18.5 ≤ BMI < 25 | 106 (25.8%) | 11 (23.4%) | 117 (25.5%) |
| Overweight 25 ≤ BMI < 30 | 153 (37.2%) | 14 (29.8%) | 167 (36.5%) |
| Obese BMI ≥ 30 | 151 (36.7%) | 21 (44.7%) | 172 (37.6%) |
| **Risk for Severe COVID-19** | | | |
| At-risk | 217 (52.8%) | 26 (55.3%) | 243 (53.1%) |
| Not at-risk | 194 (47.2%) | 21 (44.7%) | 215 (46.9%) |
| **Age, Risk for Severe COVID-19** | | | |
| Age 18-59 At-risk | 106 (25.8%) | 13 (27.7%) | 119 (26.0%) |
| Age 18-59 Not at-risk | 103 (25.1%) | 11 (23.4%) | 114 (24.9%) |
| Age ≥ 60 At-risk | 111 (27.0%) | 13 (27.7%) | 124 (27.1%) |
| Age ≥ 60 Not at-risk | 91 (22.1%) | 10 (21.3%) | 101 (22.1%) |
| **Sex Assigned at Birth** | | | |
| Female | 191 (46.5%) | 21 (44.7%) | 212 (46.3%) |
| Male | 220 (53.5%) | 26 (55.3%) | 246 (53.7%) |
| **Hispanic or Latino Ethnicity** | | | |
| Hispanic or Latino | 104 (25.3%) | 9 (19.1%) | 113 (24.7%) |
| Not Hispanic or Latino | 292 (71.0%) | 36 (76.6%) | 328 (71.6%) |
| Not reported and unknown | 15 (3.6%) | 2 (4.3%) | 17 (3.7%) |
| **Race** | | | |
| White | 268 (65.2%) | 30 (63.8%) | 298 (65.1%) |
| Black or African American | 93 (22.6%) | 13 (27.7%) | 106 (23.1%) |
| Asian | 13 (3.2%) | | 13 (2.8%) |
| American Indian or Alaska Native | 11 (2.7%) | 1 (2.1%) | 12 (2.6%) |
| Native Hawaiian or Other Pacific Islander | 2 (0.5%) | | 2 (0.4%) |
| Multiracial | 7 (1.7%) | 2 (4.3%) | 9 (2.0%) |
| Not reported and unknown | 17 (4.1%) | 1 (2.1%) | 18 (3.9%) |
| **Underrepresented Minority Status in the U.S.** | | | |
| White Non-Hispanic | 208 (50.6%) | 24 (51.1%) | 232 (50.7%) |
| Communities of Color | 203 (49.4%) | 23 (48.9%) | 226 (49.3%) |
| **Country** | | | |
| United States | 411 (100.0%) | 47 (100.0%) | 458 (100.0%) |
| **HIV Status** | | | |

| | | | |
|---|---|---|---|
| Negative | 406 (98.8%) | 46 (97.9%) | 452 (98.7%) |
| Living with HIV | 5 (1.2%) | 1 (2.1%) | 6 (1.3%) |

This table summarizes characteristics of per-protocol participants in the United States subset of the immunogenicity subcohort (strata 1-8), which was randomly sampled from the study cohort. The sampling was stratified by strata defined by enrollment characteristics: Assigned randomization arm × Baseline SARS-CoV-2 seronegative vs. seropositive × Randomization strata.

**Supplementary Table 4. Demographics and clinical characteristics of baseline SARS-CoV-2 seronegative per-protocol trial participants in the Latin America subset of the immunogenicity subcohort (strata 9-12) and thus have D1, D29 antibody data.**

| Characteristics | Vaccine (N = 214) | Placebo (N = 25) | Total (N = 239) |
|---|---|---|---|
| **Age** | | | |
| Age 18-59 | 107 (50.0%) | 12 (48.0%) | 119 (49.8%) |
| Age ≥ 60 | 107 (50.0%) | 13 (52.0%) | 120 (50.2%) |
| Mean (Range) | 49.2 (18.0, 80.0) | 50.4 (18.0, 80.0) | 49.3 (18.0, 80.0) |
| **BMI** | | | |
| Underweight BMI < 18.5 | 1 (0.5%) | | 1 (0.4%) |
| Normal 18.5 ≤ BMI < 25 | 53 (24.8%) | 6 (24.0%) | 59 (24.7%) |
| Overweight 25 ≤ BMI < 30 | 92 (43.0%) | 12 (48.0%) | 104 (43.5%) |
| Obese BMI ≥ 30 | 68 (31.8%) | 7 (28.0%) | 75 (31.4%) |
| **Risk for Severe COVID-19** | | | |
| At-risk | 108 (50.5%) | 11 (44.0%) | 119 (49.8%) |
| Not at-risk | 106 (49.5%) | 14 (56.0%) | 120 (50.2%) |
| **Age, Risk for Severe COVID-19** | | | |
| Age 18-59 At-risk | 54 (25.2%) | 5 (20.0%) | 59 (24.7%) |
| Age 18-59 Not at-risk | 53 (24.8%) | 7 (28.0%) | 60 (25.1%) |
| Age ≥ 60 At-risk | 54 (25.2%) | 6 (24.0%) | 60 (25.1%) |
| Age ≥ 60 Not at-risk | 53 (24.8%) | 7 (28.0%) | 60 (25.1%) |
| **Sex Assigned at Birth** | | | |
| Female | 82 (38.3%) | 8 (32.0%) | 90 (37.7%) |
| Male | 132 (61.7%) | 17 (68.0%) | 149 (62.3%) |
| **Hispanic or Latino Ethnicity** | | | |
| Hispanic or Latino | 203 (94.9%) | 23 (92.0%) | 226 (94.6%) |
| Not Hispanic or Latino | 7 (3.3%) | | 7 (2.9%) |
| Not reported and unknown | 4 (1.9%) | 2 (8.0%) | 6 (2.5%) |
| **Race** | | | |
| White | 128 (59.8%) | 14 (56.0%) | 142 (59.4%) |
| Black or African American | 8 (3.7%) | 1 (4.0%) | 9 (3.8%) |
| Asian | 1 (0.5%) | | 1 (0.4%) |
| Multiracial | 21 (9.8%) | 3 (12.0%) | 24 (10.0%) |
| Not reported and unknown | 9 (4.2%) | 2 (8.0%) | 11 (4.6%) |
| **Country** | | | |
| Argentina | 33 (15.4%) | 2 (8.0%) | 35 (14.6%) |
| Brazil | 89 (41.6%) | 12 (48.0%) | 101 (42.3%) |
| Chile | 11 (5.1%) | 3 (12.0%) | 14 (5.9%) |
| Colombia | 53 (24.8%) | 5 (20.0%) | 58 (24.3%) |
| Mexico | 7 (3.3%) | 2 (8.0%) | 9 (3.8%) |
| Peru | 21 (9.8%) | 1 (4.0%) | 22 (9.2%) |
| **HIV Status** | | | |
| Negative | 207 (96.7%) | 25 (100.0%) | 232 (97.1%) |

| | | |
|---|---|---|
| Living with HIV | 7 (3.3%) | 7 (2.9%) |

This table summarizes characteristics of per-protocol participants in the Latin America subset of the immunogenicity subcohort (strata 9-12), which was randomly sampled from the study cohort. The sampling was stratified by strata defined by enrollment characteristics: Assigned randomization arm × Baseline SARS-CoV-2 seronegative vs. seropositive × Randomization strata.

**Supplementary Table 5. Demographics and clinical characteristics of baseline SARS-CoV-2 seronegative per-protocol trial participants in the South Africa subset of the immunogenicity subcohort (strata 13-16) and thus have D1, D29 antibody data.**

| Characteristics | Vaccine (N = 201) | Placebo (N = 18) | Total (N = 219) |
|---|---|---|---|
| **Age** | | | |
| Age 18-59 | 96 (47.8%) | 6 (33.3%) | 102 (46.6%) |
| Age ≥ 60 | 105 (52.2%) | 12 (66.7%) | 117 (53.4%) |
| Mean (Range) | 50.6 (18.0, 80.0) | 53.3 (18.0, 80.0) | 50.8 (18.0, 80.0) |
| **BMI** | | | |
| Underweight BMI < 18.5 | 16 (8.0%) | 1 (5.6%) | 17 (7.8%) |
| Normal 18.5 ≤ BMI < 25 | 71 (35.3%) | 3 (16.7%) | 74 (33.8%) |
| Overweight 25 ≤ BMI < 30 | 49 (24.4%) | 8 (44.4%) | 57 (26.0%) |
| Obese BMI ≥ 30 | 65 (32.3%) | 6 (33.3%) | 71 (32.4%) |
| **Risk for Severe COVID-19** | | | |
| At-risk | 104 (51.7%) | 8 (44.4%) | 112 (51.1%) |
| Not at-risk | 97 (48.3%) | 10 (55.6%) | 107 (48.9%) |
| **Age, Risk for Severe COVID-19** | | | |
| Age 18-59 At-risk | 48 (23.9%) | 3 (16.7%) | 51 (23.3%) |
| Age 18-59 Not at-risk | 48 (23.9%) | 3 (16.7%) | 51 (23.3%) |
| Age ≥ 60 At-risk | 56 (27.9%) | 5 (27.8%) | 61 (27.9%) |
| Age ≥ 60 Not at-risk | 49 (24.4%) | 7 (38.9%) | 56 (25.6%) |
| **Sex Assigned at Birth** | | | |
| Female | 100 (49.8%) | 8 (44.4%) | 108 (49.3%) |
| Male | 101 (50.2%) | 10 (55.6%) | 111 (50.7%) |
| **Hispanic or Latino Ethnicity** | | | |
| Not Hispanic or Latino | 193 (96.0%) | 18 (100.0%) | 211 (96.3%) |
| Not reported and unknown | 8 (4.0%) | | 8 (3.7%) |
| **Race** | | | |
| White | 38 (18.9%) | 4 (22.2%) | 42 (19.2%) |
| Black or African American | 144 (71.6%) | 12 (66.7%) | 156 (71.2%) |
| Asian | 1 (0.5%) | | 1 (0.5%) |
| Native Hawaiian or Other Pacific Islander | 1 (0.5%) | | 1 (0.5%) |
| Multiracial | 12 (6.0%) | 2 (11.1%) | 14 (6.4%) |
| Not reported and unknown | 5 (2.5%) | | 5 (2.3%) |
| **Country** | | | |
| South Africa | 201 (100.0%) | 18 (100.0%) | 219 (100.0%) |
| **HIV Status** | | | |
| Negative | 185 (92.0%) | 17 (94.4%) | 202 (92.2%) |
| Living with HIV | 16 (8.0%) | 1 (5.6%) | 17 (7.8%) |

This table summarizes characteristics of per-protocol participants in the South Africa subset of the immunogenicity subcohort (strata 13-16), which was randomly sampled from the study cohort. The sampling was stratified by strata defined by enrollment characteristics: Assigned randomization arm × Baseline SARS-CoV-2 seronegative vs. seropositive × Randomization strata.

**Supplementary Table 6. D29 antibody marker response rates and geometric means (GMs) by COVID-19 outcome status, presented by ENSEMBLE geographic region.** Analysis based on baseline SARS-CoV-2 seronegative per-protocol vaccine recipients in the case-cohort set. Median (interquartile range) days from vaccination to D29 was 29 (3) for Latin America, 29 (2) for South Africa, and 29 (2) for the United States. RBD, receptor-binding domain.

| | COVID-19 Cases[1] | | | Non-Cases in Immunogenicity Subcohort[2] | | | Comparison | |
|---|---|---|---|---|---|---|---|---|
| D29 Marker | N | Positive Response Rate (95% CI) | GM (95% CI) | N | Positive Response Rate (95% CI) | GM (95 % CI) | Response Rate Difference (Cases – Non-Cases) | Ratio of GM (Cases/Non-Cases) |
| **Latin America** | | | | | | | | |
| Anti Spike IgG (BAU/ml) | 48 | 79.2% (65.0%, 88.6%) | 29.90 (21.37, 41.83) | 212 | 84.1% (78.1%, 88.7%) | 32.62 (27.88, 38.16) | -4.9% (-19.9, 6.3%) | 0.92 (0.69, 1.22) |
| Anti RBD IgG (BAU/ml) | 48 | 75.0% (60.5%, 85.5%) | 27.98 (20.74, 37.75) | 212 | 81.4% (75.0%, 86.5%) | 31.73 (27.54, 36.57) | -6.4% (-21.8, 5.9%) | 0.88 (0.68, 1.14) |
| Pseudovirus-nAb ID50 (IU50/ml) | 48 | 43.8% (30.2%, 58.3%) | 3.91 (2.66, 5.73) | 212 | 60.0% (52.3%, 67.3%) | 5.62 (4.56, 6.93) | -16.3% (-31.7%, 0.2%) | 0.70 (0.49, 0.98) |
| **South Africa** | | | | | | | | |
| Anti Spike IgG (BAU/ml) | 15 | 80.0% (49.8%, 94.2%) | 32.70 (18.51, 57.78) | 200 | 87.3% (81.2%, 91.7%) | 37.90 (31.72, 45.28) | -7.3% (-37.8, 8.1%) | 0.86 (0.59, 1.26) |
| Anti RBD IgG (BAU/ml) | 15 | 73.3% (43.8%, 90.6%) | 34.12 (19.30, 60.33) | 200 | 84.3% (77.6%, 89.3%) | 35.30 (30.12, 41.36) | -11% (-40.9, 7.6%) | 0.97 (0.67, 1.40) |
| Pseudovirus-nAb ID50 (IU50/ml) | 15 | 33.3% (13.3%, 62.0%) | 3.05 (1.63, 5.70) | 200 | 51.5% (43.3%, 59.7%) | 4.99 (3.91, 6.37) | -18.2% (-39.9, 11.6%) | 0.61 (0.37, 1.00) |
| **United States** | | | | | | | | |
| Anti Spike IgG (BAU/ml) | 29 | 79.3% (59.8%, 90.8%) | 25.86 (18.15, 36.84) | 409 | 85.8% (81.3%, 89.4%) | 34.24 (30.35, 38.63) | -6.5% (-26.3, 5.8%) | 0.76 (0.58, 0.99) |
| Anti RBD IgG (BAU/ml) | 29 | 72.4% (52.7%, 86.1%) | 24.00 (17.25, 33.38) | 409 | 80.2% (75.2%, 84.4%) | 32.49 (29.08, 36.30) | -7.7% (-27.9, 6.8%) | 0.74 (0.57, 0.96) |
| Pseudovirus-nAb ID50 (IU50/ml) | 29 | 27.6% (13.9%, 47.3%) | 2.40 (1.68, 3.44) | 409 | 54.4% (48.6%, 60.1%) | 4.40 (3.83, 5.06) | -26.8% (-41.6, -6.3%) | 0.55 (0.41, 0.72) |

[1]Cases are baseline SARS-CoV-2 seronegative per-protocol vaccine recipients with the primary COVID-19 endpoint (moderate to severe-critical COVID-19 with onset that was both ≥ 28 days post-vaccination and ≥ 1 day post-D29) and before the end of the correlates study period, which is up to 54 days post D29 but no later than the data cut (January 22, 2021).
[2]Non-cases/Controls are baseline seronegative per-protocol vaccine recipients sampled into the immunogenicity subcohort with no evidence of SARS-CoV-2 infection up to the end of the correlates study period, which is up to 54 days post D29 but no later than the data cut (January 22, 2021). See Extended Data Figure 2. CI, confidence interval; GM, geometric mean.

**Supplementary Table 7. Covariate-adjusted hazard ratios of COVID-19 per standard deviation (SD) increment in each D29 antibody marker in baseline SARS-CoV-2 seronegative per-protocol vaccine recipients.**

| D29 Immunologic Marker | No. cases/ No. at-risk* | HR per SD increment Pt. Est. | 95% CI | P-value (2-sided) | FDR-adjusted p-value** | FWER-adjusted p-value |
|---|---|---|---|---|---|---|
| Anti Spike IgG (BAU/ml) | 140/18,395 | 0.84 | (0.66, 1.07) | 0.162 | 0.189 | 0.255 |
| Anti RBD IgG (BAU/ml) | 140/18,395 | 0.80 | (0.62, 1.03) | 0.079 | 0.150 | 0.144 |
| Pseudovirus-nAb ID50 (IU50/ml) | 140/18,395 | 0.65 | (0.48, 0.88) | 0.006 | 0.015 | 0.016 |

Baseline covariates adjusted for: baseline risk score, geographic region. RBD, receptor-binding domain.

*No. at-risk = estimated number in the population for analysis: baseline negative per-protocol vaccine recipients not experiencing the COVID-19 endpoint and with no evidence of SARS-CoV-2 infection through D29; no. cases = estimated number of this cohort with an observed COVID-19 endpoint starting ≥ 1 day post D29 and ≥ 28 days post-vaccination. The count 140 differs from the number 92 (Figure 1, Table 1), because the 140 includes all vaccine breakthrough cases including those without D1, 29 antibody marker data.

**FDR (false discovery rate)-adjusted p-values and FWER (family-wise error rate)-adjusted p-values are computed over the set of p-values both for quantitative markers and categorical markers (Low, Medium, High) using the Westfall and Young permutation method (10000 replicates).

**Supplementary Table 8. Sensitivity analysis to assess D29 antibody markers categorized as upper vs. lower tertiles as controlled vaccine efficacy correlates of protection against COVID-19.**

| | Marginalized Risk Ratio $RR_M(0,1)$[1] | | Controlled Risk Ratio = (1-CVE(1))/(1-CVE(0))[2] | | E-values[3] | |
|---|---|---|---|---|---|---|
| D29 Antibody Marker | Point Estimate | 95% CI | Point Estimate | 95% CI | For Point Estimate | For 95% CI Upper Limit |
| Anti-spike IgG (BAU/ml) | 0.75 | 0.40, 1.31 | 1.00 | 0.53, 1.75 | 2.0 | 1.0 |
| Anti-RBD IgG (BAU/ml) | 0.61 | 0.33, 1.08 | 0.82 | 0.43, 1.43 | 2.6 | 1.0 |
| ID50 (IU50/ml) | 0.41 | 0.21, 0.69 | 0.55 | 0.28, 0.92 | 4.3 | 2.2 |

[1] This analysis estimates the Controlled Risk Ratio under the no-unmeasured confounding and positivity assumptions.

[2] Conservative (upper bound) estimate assuming unmeasured confounding at level $RR_{UD}(0, 1) = RR_{EU}(0, 1) = 2$ and thus $B(0, 1) = 4/3$ (notation as in Ding and vanderWeele (2016)).

[3] E-values are computed for upper tertile ($s = 1$) vs. lower tertile ($s = 0$) biomarker subgroups after controlling for baseline risk score and geographic region; UL = upper limit.

**Supplementary Table 9. Assay limits of the three antibody markers evaluated as immune correlates.** BAU = binding antibody units; IU = International Units; LLOQ, lower limit of quantification; RBD, receptor binding domain; ULOQ, upper limit of quantification.

| MSD Binding Assay (VRC) | | |
|---|---|---|
| Reported units | BAU/ml | |
| | Spike | RBD |
| Positivity Cutoff | 10.8424 | 14.0858 |
| LOD | 0.3076 | 1.5936 |
| LLOQ | 1.8429 | 5.0243 |
| ULOQ | 238.1165 | 172.5755 |
| All values < Positivity Cutoff were set to Positivity Cutoff/2 | | |
| All values > ULOQ were set to ULOQ (for immune correlates analyses) | | |
| **Pseudovirus neutralization ID50 titer (Monogram)** | | |
| Reported units | IU50/ml | |
| LOD | 2.612 | |
| LLOQ | 2.7426 | |
| ULOQ | 619.3052 | |
| All values < LLOQ were set to LLOQ/2 | | |
| All values > ULOQ were set to ULOQ (for immune correlates analyses) | | |

**Supplementary Table 10. Individual baseline variables input into the Superlearner model for predicting occurrence of the COVID-19 primary endpoint in baseline SARS-CoV-2 seronegative per-protocol placebo recipients.**

| Variable Name | Definition | Total missing values |
|---|---|---|
| EthnicityHispanic | Indicator ethnicity = Hispanic (1 = Hispanic, 0 =complement) | 0/18116 (0.0%) |
| EthnicityNotreported | Indicator ethnicity = Not reported (1 = Not reported, 0 =complement) | 0/18116 (0.0%) |
| Black | Indicator race = Black (1=Black, 0=complement) | 0/18116 (0.0%) |
| Asian | Indicator race = Asian (1=Asian, 0=complement) | 0/18116 (0.0%) |
| NatAmer | Indicator race = American Indian or Alaska Native (1=NatAmer, 0=complement) | 0/18116 (0.0%) |
| Multiracial | Indicator race = Multiracial (1=Multiracial, 0=complement) | 0/18116 (0.0%) |
| Notreported | Indicator race = Not reported (1=Notreported, 0=complement) | 0/18116 (0.0%) |
| Unknown | Indicator race = unknown (1=Unknown, 0=complement) | 0/18116 (0.0%) |
| URMforsubcohortsampling | Indicator of under-represented minority (1=Yes, 0=No) | 0/18116 (0.0%) |
| HighRiskInd | Baseline covariate indicating >= 1 Co-existing conditions | 0/18116 (0.0%) |
| HIVinfection | (1=yes, 0=no, NA=missing)<br>Indicator living with HIV at enrollment (1=living with HIV, 0=not living with HIV) | 0/18116 (0.0%) |
| Sex | Sex assigned at birth (1=female, 0=male/undifferentiated/unknown) | 0/18116 (0.0%) |
| Age | Age at enrollment in years (integer >= 18, NA=missing). Note that the randomization strata included Age 18-59 vs. Age >=60. | 0/18116 (0.0%) |
| BMI | BMI at enrollment (Ordered categorical 1,2, 3, 4, NA=missing); 1 = Underweight BMI < 18.5; 2 = Normal BMI 18.5 to < 25; 3 = Overweight BMI 25 to < 30; 4 = Obese BMI >= 30 | 15/18116 (0.1%) |
| Country.X1 | Indicator country = Argentina (1 = Argentina, 0 = complement) | 0/18116 (0.0%) |
| Country.X2 | Indicator country = Brazil (1 = Brazil, 0 = complement) | 0/18116 (0.0%) |
| Country.X4 | Indicator country = Colombia (1 = Colombia, 0 = complement) | 0/18116 (0.0%) |
| Country.X6 | Indicator country = Peru (1 = Peru, 0 = complement) | 0/18116 (0.0%) |
| Country.X7 | Indicator country = South Africa (1 = South Africa, 0 = complement) | 0/18116 (0.0%) |
| Region.X1 | Indicator region = Latin America (1 = Latin America, 0 = complement) | 0/18116 (0.0%) |

| | | |
|---|---|---|
| Region.X2 | Indicator country = Southern Africa (1 = Southern Africa, 0 = complement) | 0/18116 (0.0%) |
| CalDtEnrollIND.X1 | Indicator variable representing enrollment occurring between 4-8 weeks periods of first subject enrolled (1 = Enrollment between 4-8 weeks, 0 = complement). | 0/18116 (0.0%) |
| CalDtEnrollIND.X2 | Indicator variable representing enrollment occurring between 8-12 weeks periods of first subject enrolled (1 = Enrollment between 8-12 weeks, 0 = complement). | 0/18116 (0.0%) |
| CalDtEnrollIND.X3 | Indicator variable representing enrollment occurring between 12-16 weeks periods of first subject enrolled (1 = Enrollment between 12-16 weeks, 0 = complement). | 0/18116 (0.0%) |

*Note:*
1. Binary input variable/s EthnicityUnknown, PacIsl, Country.X3, Country.X5 had <= 3 cases in the variable = 1 or 0 subgroup and were dropped from analysis.
2. No input variable had more than 5% missing values.
3. BMI had less than 5% missing values. The missing values were imputed using the mice package in R.

Supplementary Text 1.

In addition to adjusting for geographic region, all correlates analyses adjust for a baseline COVID-19 risk score that was developed through machine learning of the baseline SARS-CoV-2 seronegative per-protocol placebo arm.
Supplementary Table 10 lists the input variables that were included in the machine learning to build a model predicting the COVID-19 endpoint, where cases are COVID-19 endpoints occurring at least 1 day post D29 and non-cases are participants with follow-up beyond 7 days post D29 visit and that never registered a COVID-19 endpoint. The risk score is defined as the logit of the predicted COVID-19 outcome probability from the predictive regression model, estimated using the ensemble algorithm superlearner (i.e. stacking), where this logit predicted outcome is scaled to have empirical mean zero and empirical standard deviation one.

The receiver operating characteristic curve for the Superlearner model built from baseline SARS-CoV-2 seronegative per-protocol placebo recipients was applied to baseline SARS-CoV-2 seronegative per-protocol vaccine recipients, yielding an AUC of 0.670 for classifying COVID-19 outcome status.

**Supplementary Figure 1. Inverse probability sampling-weighted empirical reverse cumulative distribution function curves for each D29 marker [(A) spike IgG, (B) receptor-binding domain (RBD) IgG, (C) ID50] in the vaccine arm and application of the Siber 2007 method for estimating a threshold of perfect vs. no protection.**

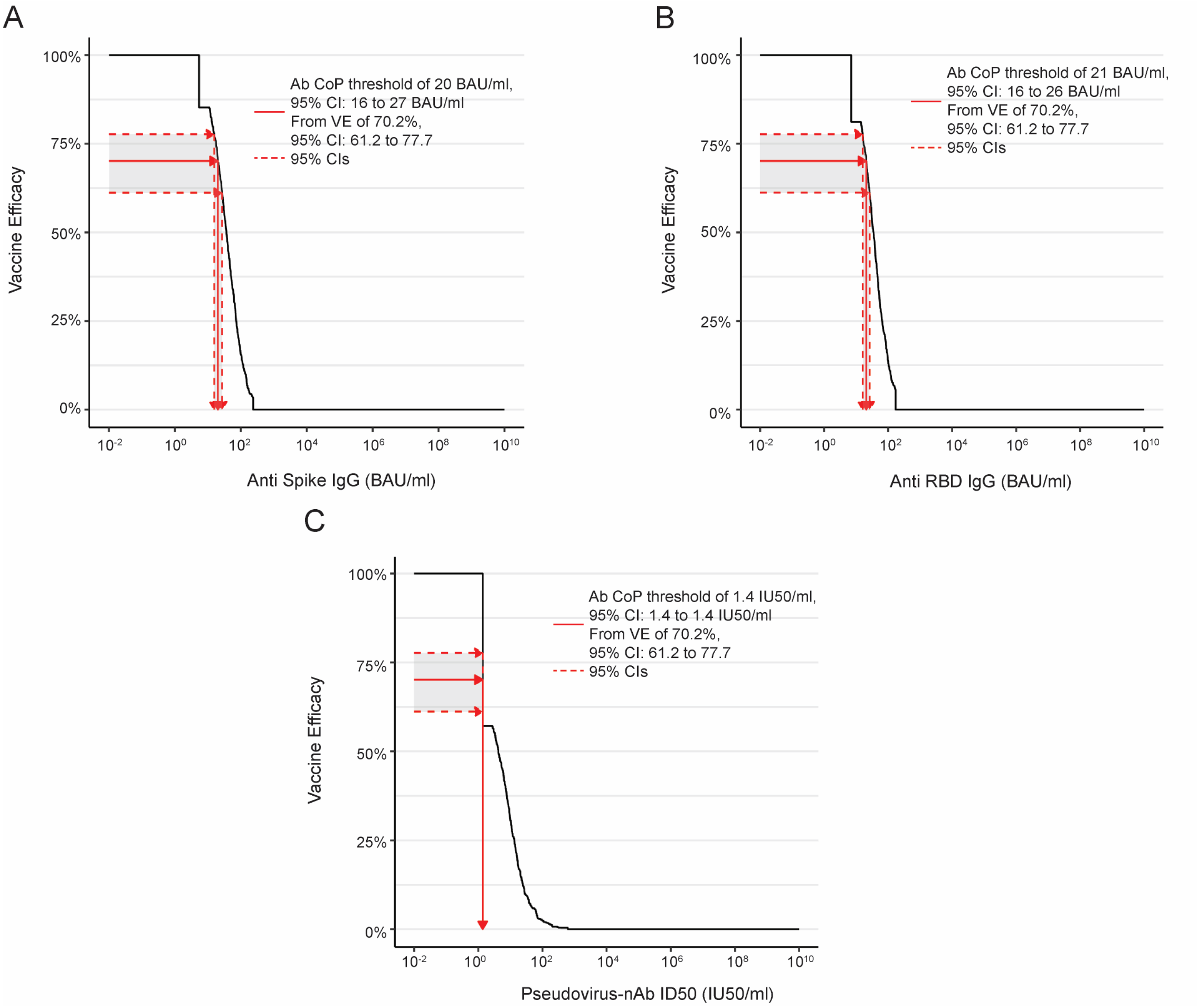


Siber GR, Chang I, Baker S, et al. Estimating the protective concentration of anti-pneumococcal capsular polysaccharide antibodies. *Vaccine* 2007; **25**(19): 3816-26.
